# A Clinical Trial to Determine the Impact of Tumor Size, Histological Subtype, and Vitamin D Status on the Therapeutic Response of Basal Cell Carcinoma to Photodynamic Therapy

**DOI:** 10.1101/2025.01.30.25321144

**Authors:** Edward V. Maytin, Nathalie C. Zeitouni, Abigail Updyke, Jeffrey Negrey, Alan S. Shen, Lauren E. Heusinkveld, Judith A. Mack, Bo Hu, Sanjay Anand, Terence A. Maytin, Laura Giostra, Christine B. Warren, Tayyaba Hasan

## Abstract

Photodynamic therapy (PDT) with topical aminolevulinic acid (ALA) can be effective for select basal cell carcinoma (BCC) lesions. However, the histological depth and subtype of tumors that respond to PDT remain uncertain. Here, we report a clinical trial of high-dose oral Vitamin D (VD), used as a PDT neoadjuvant for BCC. In this multi-institutional, intra-patient, randomized trial, 35 patients (9 with Gorlin Syndrome) received three PDT sessions (20% ALA; 417 nm blue light) preceded by oral VD, placebo, or no pretreatment. Tumors (122 BC) were monitored using 3D photography and computer-assisted volumetric analysis. Values for absolute volume (3DAbsVol) and average height (3DAvHt) were calculated and used to quantify tumor response kinetics. From histological sections, 3DAvHt was found to correlate with actual tumor depth, although 3DAvHt is only ∼10-20% of the latter. Importantly, 3DAvHt measurements revealed a distinct depth threshold that predicts PDT responsiveness. Of 122 tumors analyzed, 70% cleared after PDT; remaining tumors were micronodular or other aggressive histologic subtypes. To evaluate VD’s effects upon treatment response kinetics after PDT, only 40% of original lesions were available for analysis. By stratifying remaining tumors by 3DAvHt, we found 65% of thin tumors to be VD-responsive, whereas only 28% of thick tumors responded to VD. Overall, PDT was effective for the majority of BCC lesions in our study. Tumors most likely to respond can be predicted histologically and by noninvasive 3D morphometry. For PDT-appropriate BCC lesions, neoadjuvant oral Vitamin D represents a safe and beneficial way to accelerate tumor resolution.

**CLINICAL TRANSLATION STATEMENT:** For photodynamic therapy (PDT) of basal cell carcinoma (BCC), a clinical challenge is deciding which tumors to treat since the penetration depth of visible light into the skin is limited. In this clinical trial, noninvasive 3D photography and computer analysis were used to determine the height and volume of BCC tumors and to correlate these calculated parameters with tumor clearance after PDT, with or without the use of oral vitamin D3 (VD) as a neoadjuvant. Two very practical findings emerged. First, tumor height (3DAvHt) was found to correlate with BCC tumor depth and to predict therapeutic response; tumors below a height threshold of 0.13 mm (∼1.5 - 2 mm histological depth) were highly likely to respond. Second, adding VD as a neoadjuvant prior to PDT of appropriate BCC tumors (superficial and thin nodular subtypes) is a safe and effective way to boost PDT efficacy.

## INTRODUCTION

Basal cell carcinomas (BCC), skin tumors which typically arise after decades of chronic sun exposure, is the most common of all human cancers (*1,2*). Although the current standard of care for BCC is surgical excision, post-surgical scars can be very problematic, especially in cosmetically sensitive areas and in patients with a high tumor burden. Examples of the latter are organ transplant recipients on immunosuppressive drugs, and patients with Gorlin syndrome (a heritable condition in which patients develop dozens-to-hundreds of BCC tumors beginning in adolescence)(*3,4*). In Gorlin patients, multiple surgeries over a lifetime are often associated with disfigurement, functional compromise, and even the development of major depression with a high risk of suicide (*5*).

Photodynamic therapy (PDT) is a non-scarring, noninvasive treatment modality for cancer in which tumor-selective accumulation of a photosensitizer (PS), either 5-aminolevulinic acid (ALA) or its methyl ester, is converted into protoporphyrin IX (PpIX) within mitochondria which is then activated using strong visible light (*6–8*). Although PDT can be used for different cancers at various locations in the body (wherever laser light can be delivered using fiber optics). PDT is in fact most commonly used for skin neoplasia because of the relative ease of shining a conventional light source (blue or red lamp or laser) onto the skin (*9*). Most European countries, Canada, and Australia have approved the use of PDT for BCC treatment. PDT is known to work quite well for superficial BCC (sBCC), with 5-year clearance rates (CR) between 71%-88% at 5 years post-treatment (*10,11*) but it is less reliable for nodular BCC (nBCC) in which the CR can be considerably lower (*12*). In general, a barrier to the more widespread use of PDT for BCC is the fact that histological and tumor-size parameters associated with a good BCC response are not yet sufficiently well-defined. In the United States, the FDA has not approved the use of PDT for treating BCC.

To try to improve the overall response of nonmelanoma skin cancer to PDT, we have worked for many years to identify agents that can be combined with PDT (as neoadjuvants) to improve lesion clearance (*13*). As one example, high-dose Vitamin D (VD) was shown to be a safe and effective neoadjuvant with PDT that improves the resolution of cutaneous squamous cell carcinoma (SCC) and actinic keratoses (pre-SCC) in murine models (*14*), and more recently in human patients (*15,16*). In the case of BCC, pretreatment with VD increases ALA-mediated PpIX accumulation in a *ptch1* mutant mouse model of BCC (*17,18*). However, *ptch1*/p53 BCC mice are not a perfect model of human disease, and therefore the question of how size and histological subtype of BCC influence PDT response in patients has remained unanswered. Therefore, we designed a clinical study in patients with BCC to systematically address the following questions: (1) How does the initial size of the BCC correlate with the number of PDT treatments needed to clear it? Can a size threshold for responsiveness be defined? (2) What is the relationship between tumor response after PDT and BCC histological subtype? (3) Does oral VD, administered as a pretreatment prior to PDT, tend to accelerate or otherwise affect the rate of BCC tumor shrinkage? For this study we developed a new high-precision 3D photographic method to accurately monitor changes in the size of individual BCC tumors. This non-invasive analytic technique is described in detail in the report, along with the results of the clinical trial.

## MATERIALS AND METHODS

### Clinical trial design

This clinical trial was registered at ClinicalTrials.gov (NCT03483441) and conducted at two different sites, Cleveland Clinic (Cleveland, Ohio) and US Dermatology Partners (Phoenix, Arizona) under related but separate IRB approvals. For inclusion, all patients were required to have a minimum number of biopsy-proven BCC tumors (2 or 3, depending upon the trial location), and up to 10 lesions per patient could be studied. At Cleveland Clinic, patients with Gorlin syndrome (BCNS; clinically defined in (*3*)) were recruited. Two histologically proven BCC were required for enrollment; any lesion that persisted after 3 rounds of PDT was biopsied. At the Arizona site, primarily chronically sun-damaged patients were recruited; 3 biopsy-proven BCC tumors were required. Exclusion criteria included pregnancy, history of renal disease or porphyria, and treatment with Vismodegib or VD supplementation within the past month.

The primary aim of the clinical study was to test whether high dose oral VD (cholecalciferol; D3), when used in combination with PDT, will hasten the kinetic clearance rate of BCC tumors. The intrapatient comparative study design featured five study visits and three different VD pretreatment conditions (**Fig. 1**). Each patient served as his/her own control for evaluation of VD effects, and overall Vitamin D status was determined at each visit by measuring levels of serum 25-hydroxy vitamin D3 (calcidiol; 25-OH D3). Some blood samples were also used to obtain leukocytes for VD receptor (VDR) gene analysis. The correct content of active VD ingredient (D3) in study drug capsules was verified by laboratory testing (Heartland Assays, Ames, Iowa). The order of administration of agents (either high-dose D3 or placebo capsules) prior to PDT was randomized 1:1 at Visits 2 and 3 by our research pharmacy, as follows. One PDT session was preceded by high dose neoadjuvant D3, 10,000 international units [IU] daily, taken for either 5 or 14 days, depending upon whether the patient’s initial serum 25-OH D3 level was normal or low, respectively. The other PDT session had no D3 pretreatment beforehand. Prior to the third PDT session (Visit 4), patients took D3 2,000 IU daily to achieve a normal serum 25-OH D3 level. Final assessments of lesion clearance were performed at 6 months (Visit 5). Tumors were photographed at every study visit using the 3D camera described below.

**Figure 1.**
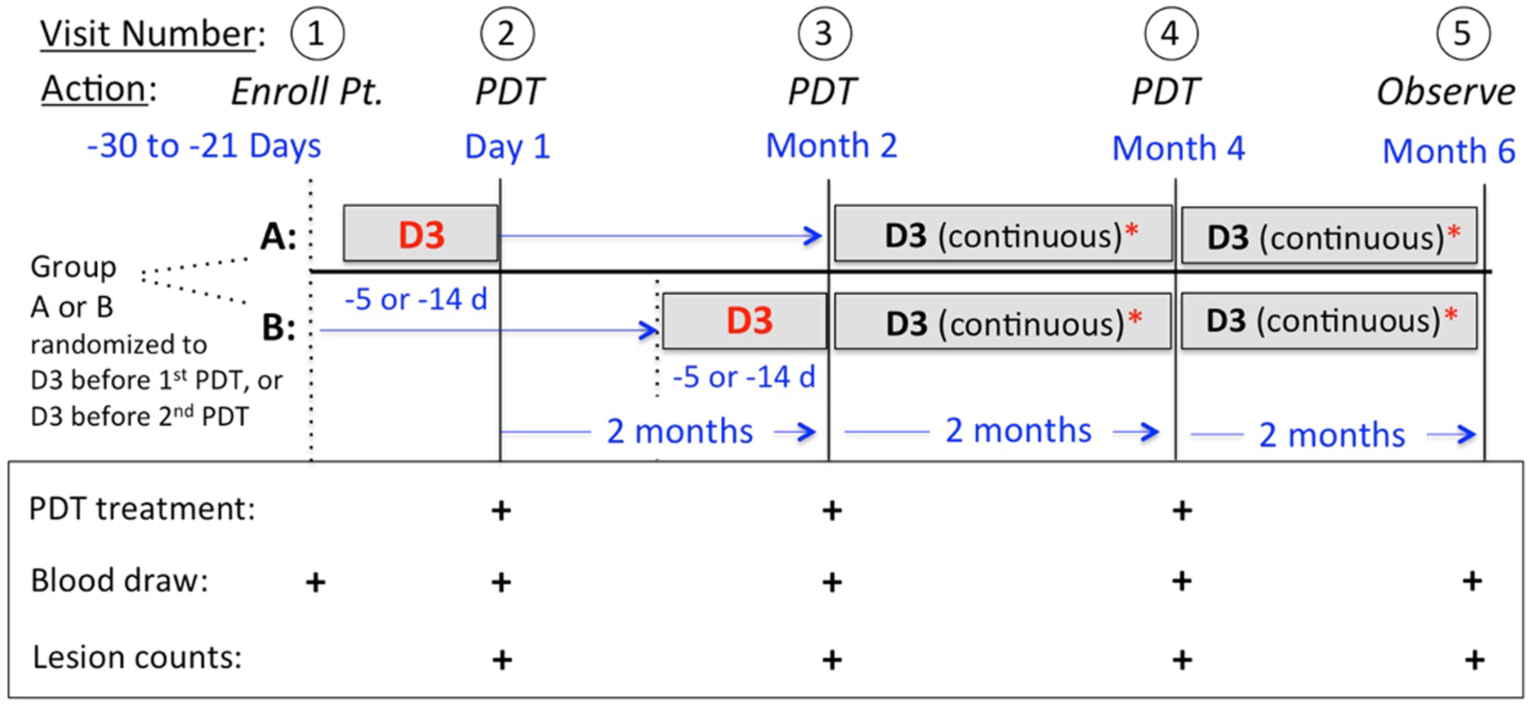
Clinical trial protocol. Patients received 10,000 units of high-dose oral cholecalciferol (“D3”) prior to only one of the first two PDT treatments. For those visits, the delivery of the neoadjuvant (either D3, or a placebo capsule) was randomized as follows. Patients in Group A received D3 prior to PDT at Visit 2, whereas patients in Group B received D3 prior to PDT at Visit 3. All patients then received D3 supplementation (2,000 units daily) prior to their PDT at Visit 4. Any lesion not resolved at Visit 5 was biopsied.

PDT treatments were administered as follows. A solution of 20% ALA (Levulan Kerastick^TM^, Sun Pharmaceuticals) was applied to each tumor and a 5 mm rim of surrounding skin, covered with an occlusive dressing, and 4 hours later illuminated with blue light (Blu-U lamp, 417 nm; 20 J/cm^2^; Sun/DUSA) as described (*19*). Pain relief was provided using ice-cold cloths as needed. Patients were sent home with instructions to avoid direct sun exposure for 48 h.

### Evaluation of BCC tumor volume and thickness using 3-D photographic quantitative analysis

Using a 3D camera (LifeViz Micro) and software (Dermapix) from Quantificare, Inc., pairs of digital images of individual tumors were taken. These were processed to create 3-D mathematical reconstructions from which a variety of tumor dimensions could be calculated. The most useful parameters for us were *absolute volume* (3DAbsVol) and *average height* (3DAvHt). For example, 3DAbsVol was plotted against time as a visual representation of tumor clearance kinetics. These methods are described in detail in **Supplementary Methods 1** *(Supp1)*.

### Evaluation of BCC histologic subtype

For each biopsied lesion, the histological diagnosis was determined by a board-certified dermatopathologist at the respective institutions. To analyze depth of BCC lesions, extra slides were recut from the original paraffin blocks, stained with hematoxylin and eosin (H&E), and the maximum depth (distance from epidermis to deepest aspect of tumor nests) was analyzed using QuPath software (*20*).

### Evaluation of Vitamin D levels and Vitamin D receptor (VDR) status

For all patients (including those from Arizona), serum 25-OH D3 analyses were performed in the clinical pathology department at Cleveland Clinic. For 24 patients, DNA was extracted from blood leukocytes to test genetic alleles of the VDR gene as previously described (*21*).

### Study coordination and data management at multiple study sites

To facilitate the secure and efficient transfer of patient information and data between the Cleveland and Arizona sites, we collaborated with Medocity, Inc. (Parsippany, NJ), a company that develops online tools for sharing of medical information. Medocity’s customized clinical trial management website allowed us to remotely share study patients’ data (recruitment status, 3-D clinical photos, biopsy reports, and lab results) in a safe and protected digital environment. Details about the Medocity platform are provided in **Supplementary Methods 2** (*Supp2)*.

## RESULTS

Raw data from the clinical trial are provided in **Table 1** and **in Supplementary Table S1** (Supp3). As described in Methods, patients were recruited at two different clinical sites, comprising 14 patients at the Cleveland site (including 9 with Gorlin syndrome) and 22 patients with at the Arizona location. Of 36 total subjects enrolled, 35 completed the entire protocol; see enrollment flow chart in **Supplementary Figure S1** (Supp4). Patients were generally older adults (median age, 62 years) with Fitzpatrick skin types I – II and male gender (70%), except for two female Gorlin patients in their mid-twenties. **Table 1** lists the demographic data and lesion counts for all patients. A total of 211 lesions were monitored during the trial. Amongst these, 89 were later excluded due to a subsequent non-BCC diagnosis (SCC; fibrosis/scar) or technical issues that precluded a complete analysis at all five study visits; see lesion flowchart in **Supplementary Figure S2** (Supp 5). Thus, 122 lesions were ultimately available for a complete analysis during the entire study time-course. All data for those 122 tumors and the original 211 lesions is provided in **Supplementary Table S1** (Supp3).

**TABLE 1.**
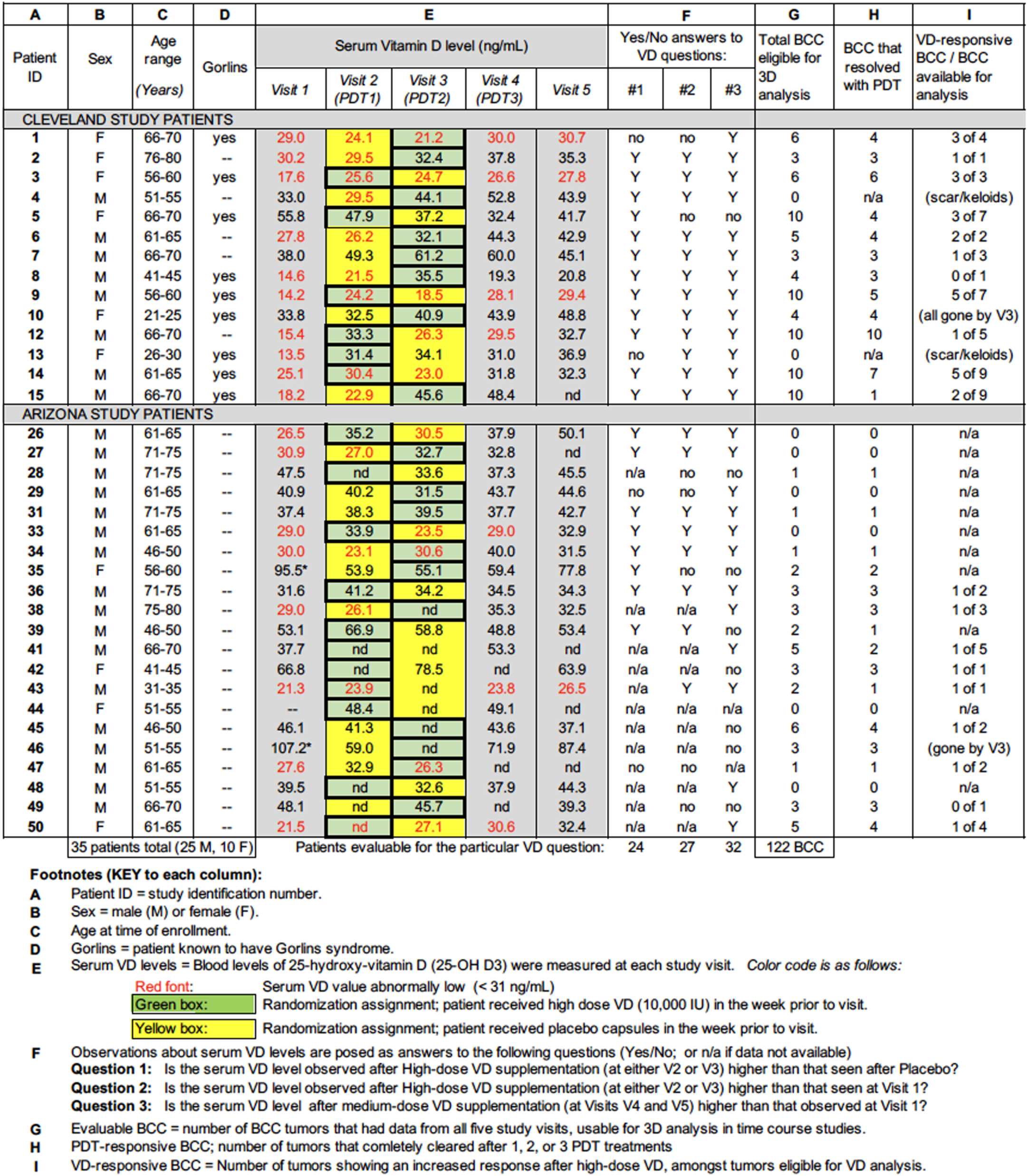
Patient demographics, Vitamin D serum levels, and BCC tumor counts.

### Approach and overall results from analysis of changes in BCC tumor size after PDT

As described in Methods, three-dimensional images (mathematical reconstructions) of tumors from dual high-resolution photographs were used to determine 3DAbsVol and 3DAvHt for each tumor, relative to the horizontal surface plane of the skin. Data were plotted, and kinetic rates of tumor shrinkage were assessed from slopes of the volume-versus-time curve. Examples of typical 3D photographic data are shown in **Figure 2**. Here, we first summarize the overall results. The large majority of BCC lesions resolved after PDT. Specifically, 41% of tumors resolved by Visit 3, 58% by Visit 4, and 70% by Visit 5, leaving 30% of the tumors classified as PDT-resistant. In general, most BCC that resolved after PDT were small, relatively thin tumors (**Fig. 2A and 2B**). Most were superficial BCC (**Fig. 2A**), but also a significant number were nodular BCC (**Fig. 2B**). Other tumors responded initially to PDT (shrinking after one or more treatments) but then grew back (**Fig. 2C** and **2D**). As an important general result, the data show that height really matters. Thus, the height (3D AvHt) of PDT-responsive lesions was typically less than for PDT-resistant lesions, probably reflecting a relationship to depth of photosensitizer and light penetration. However, exceptions to the apparent inverse correlation between tumor height and PDT responsiveness are observed for PDT-resistant tumors with aggressive histological subtypes, such as the one illustrated in **Fig 2E** which showed no inhibition in tumor growth rate after PDT, despite being relatively thin.

**Figure 2.**
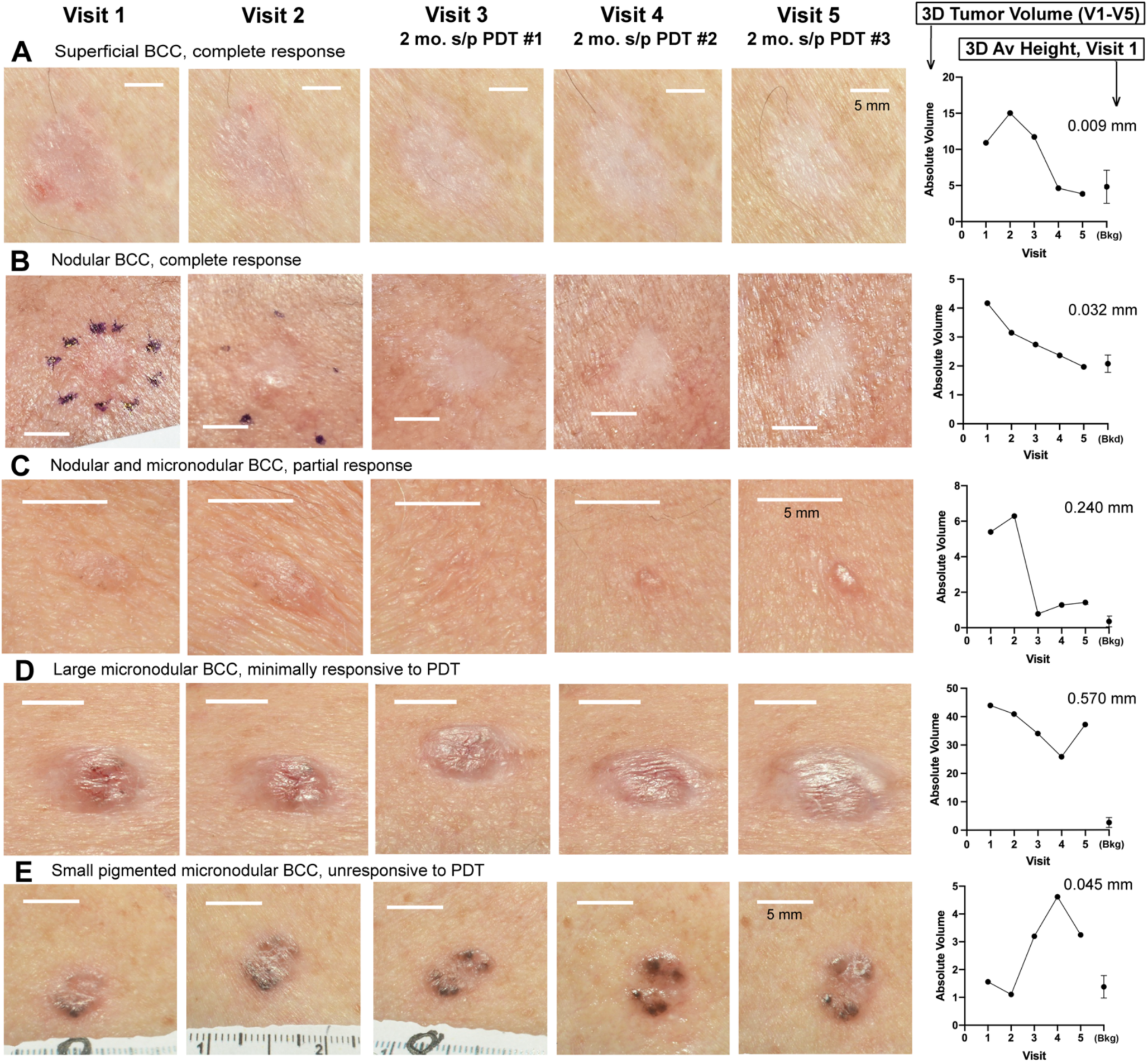
3D reconstructed images of BCC tumors photographed at each of five study visits, to show typical appearances and corresponding changes in absolute tumor volume for each lesion calculated using the DermaPix software. Absolute Volume (in mm^3^) of each tumor, over the time course of 5 study visits, along with the average background value (Bkg), is shown in the graphs. The 3D average height (in mm) of each tumor at Visit 1 is indicated at the upper right of the graphs. Scale bars, 5 mm.

### Influence of BCC tumor size upon responsiveness to PDT

To more precisely define the influence of tumor volume and thickness upon therapeutic response, we focused on two size parameters, *absolute volume* (3DAbsVol) and *3D average height* (3DAvHt). The time course graphs constructed using 3DAbsVol are compiled in **Supplementary Table S1**. Evaluable tumors were grouped into 4 categories, based upon their ultimate clearance status at Visit 5; see **Fig. 3**.

**Fig. 3.**
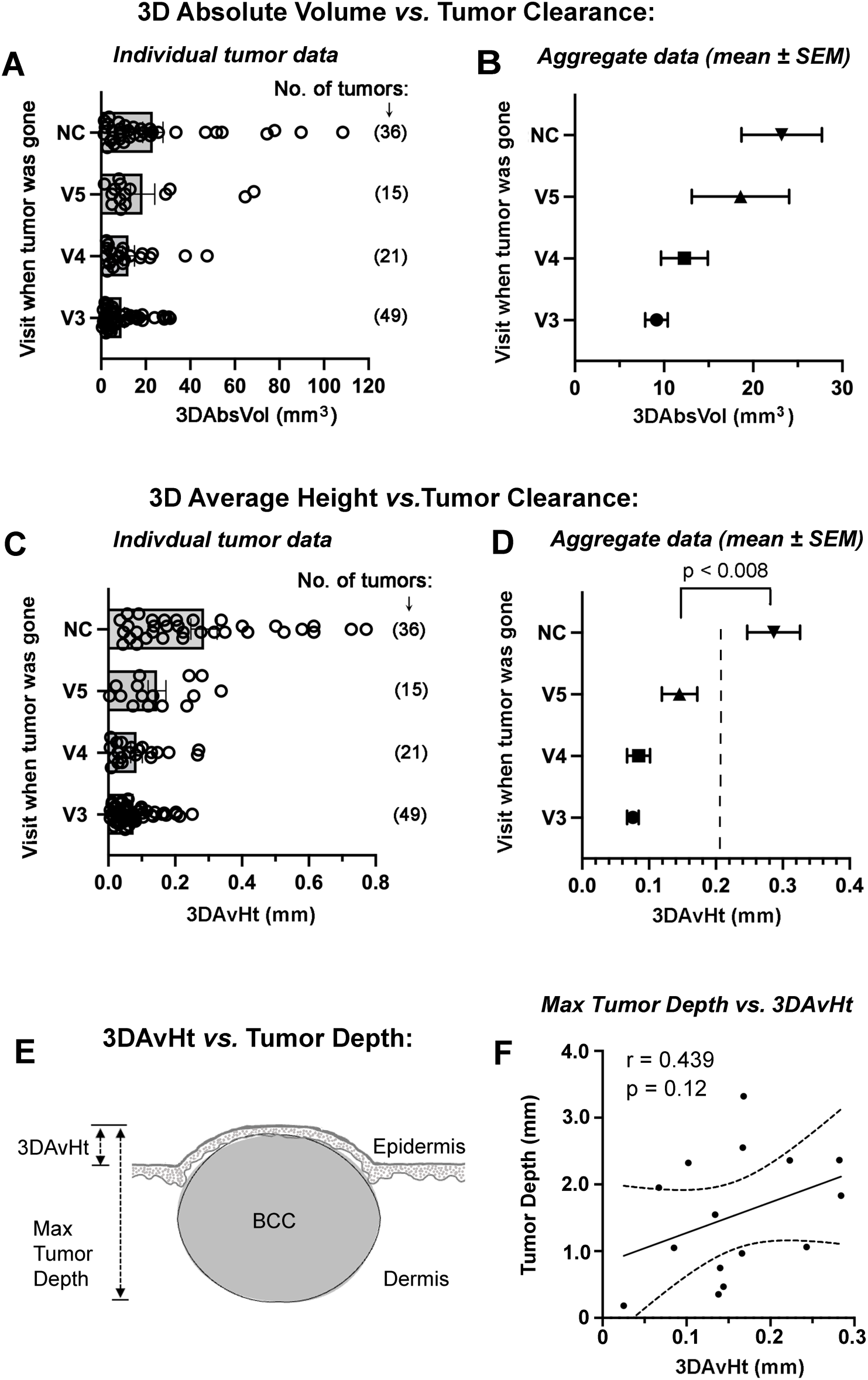
Correlation study between calculated size parameters from 3D analysis of BCC tumors, *versus* tumor clearance as indicated by disappearance of tumor at Visits V3, V4, or V5 (after 1, 2, or 3 PDT treatments, respectively). *NC*, tumors that failed to clear after 3 PDT treatments. (**A, B**), *3D Absolute Volume*, shown as data for individual tumors or in aggregate. (**C, D**) *3D Average height*, shown as data for individual tumors or in aggregate. *Dotted line* in panel ***D*** is apparent threshold of 3DAvHt. Beyond this threshold, BCC are unlikely to respond to PDT. The p-value from Student t-test is shown. (**E**) Schematic diagram showing the definitions of 3DAvHt and histological tumor depth. (**F**) Plot to show the modest correlation between 3DAvHt and histologic depth for a subset of tumors that were biopsied at the same visit; maximum depth of tumor nests was measured from digital images of H&E stained histological sections; depths are in millimeters.

Tumors that cleared completely on clinical exam were grouped by the visit of first disappearance (either V3, V4, or V5). Tumors that persisted at V5 were labelled as *not cleared* (“NC”). Considering absolute volume, a negative relationship between 3DAbsVol and number of PDT treatments to clear the tumor was observed, as expected (**Fig. 3A, B**). However, 3DAbsVol did not provide the ability to unequivocally distinguish between cleared versus persistent tumors. 3DAvHt, on the other hand, revealed large differences between tumors that cleared after 1 - 3 PDT treatments and those that failed to clear (**Fig. 3C, D**). For 3DAvHt, differences between PDT-responsive and PDT-resistant groups were statistically significant. Interestingly, a 3DAvHt value of ∼0.2 mm appears to represent an operational threshold; i.e., tumors with values > 0.2 are unlikely to respond to PDT, whereas those below the threshold have a high likelihood of response (**Fig. 3D**, dotted line).

To understand how 3DAvHt might relate to the true intradermal depth of the tumor (since 3DAvHt is a calculated height and may or may not be related to actual histological depth of the tumor; see **Fig. 3E**), we performed the following analysis. For BCC tumors that were photographed immediately prior to biopsy at V5, the 3DAvHt was compared to the depth of tumor nests observed in the corresponding H&E stained specimen. A roughly linear correlation between 3DAvHt and maximum depth of tumor was observed (**Fig. 3F**). Although 3DAvHt values were only ∼10-20% of the actual tumor depth, a positive correlation between the two parameters was seen, even when tumors were stratified by histological subtype (**Supplementary Table S2**). Thus, we conclude that 3DAvHt represents a surrogate measure of dermal depth for these BCC tumors.

### Influence of BCC histologic subtype upon responsiveness to PDT

From an analysis of 36 tumors diagnosed by pre-treatment biopsy, we examined the relationship between histological subtype and number of treatments required to achieve tumor clearance. Amongst these lesions, 100% of superficial BCCs cleared completely after 1 - 3 PDT sessions (**Fig. 4A**). For nodular BCC, many failed to respond, yet a significant number (61%) did clear (**Fig. 4B**). For tumors that were biopsied because they failed to clear (**Fig. 4C**), histological subtypes (e.g. micronodular; infiltrative) different from the usual superficial and nodular variants (**Supplementary Table S2**) were predominantly found. The numerical distribution ratios for tumors amongst the various histological subtypes, was 1: 6: 12: 3: 3: 1 for Superficial: Nodular: Micronodular: Infiltrative: Trichoepithelial: Adenoid in PDT-resistant tumors (**Supplementary Table S2**), versus 16: 9 for sBCC: nBCC (with no rare histologic subtypes) amongst PDT-responsive tumors (**Table 1**, Arizona patients). To ask whether there was any additional way to demonstrate that rare BCC subtypes have more aggressive biological behavior, we hypothesized that repeated PDT treatments tend to select for smaller, more aggressive tumors. This is because a PDT dose that would kill a typical BCC fails to kill the aggressive tumor which continues to proliferate after PDT, despite being relatively thin and accessible to drug and light. To test this idea, two groups of PDT-resistant BCC (10 nodular *versus* 15 aggressive histological subtypes) were compared to determine their initial sizes at Visit 1; see **Supplementary Table S3 (Supp7**). For these tumors, the mean 3DAbsVol was 3-fold less for aggressive subtypes compared to nodular BCC, and the mean 3D AvHt was 2-fold smaller. This supports the notion that BCC with aggressive histologic subtypes can resist PDT even when relatively small in size.

**Fig. 4.**
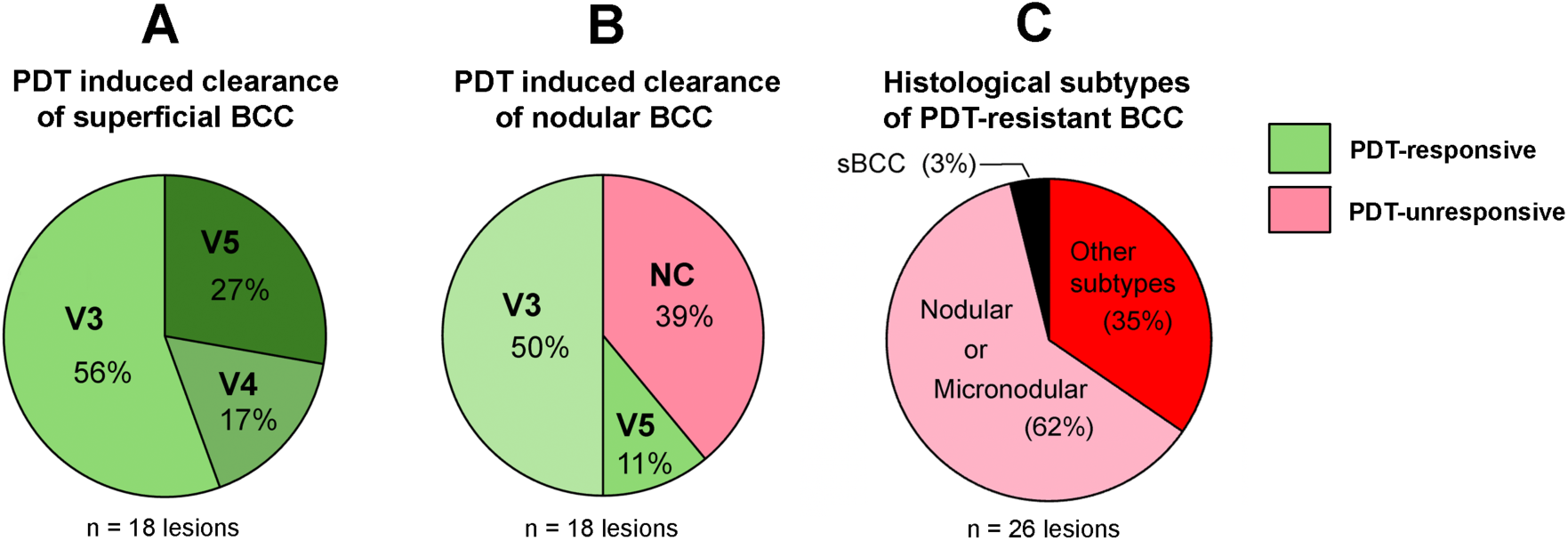
Percentage of tumors that cleared at visits V3, V4, or V5 (*green color*), or failed to respond (*red color*), within categories of superficial (**A**) or nodular (**B**) BCC. Amongst PDT-resistant lesions biopsied at V5 to diagnose the histology of the resistant tumor (**C**), 97% were relatively aggressive BCC subtypes and included a mixture of nodular and micronodular, as well as other rare subtypes including adenoid, infiltrative, and trichoepitheliomatous variants.

### Influence of Vitamin D upon BCC responsiveness to PDT

One of the first observations in Table 1 is that baseline vitamin D status (25-O-D3 levels) were different in the Cleveland and Arizona cohorts, i.e., 71% of Cleveland patients compared to 40% of Arizona patients were Vitamin D deficient at Visit 1. This might be due to a relative lack of sunlight in Cleveland vs. Phoenix, or to the fact that 65% of the Cleveland patients had Gorlin syndrome; the latter is known to be associated with low serum VD levels, for unknown reasons (*22*). Regardless, baseline 25-OH-D3 levels did not seem to have any discernible effect upon PDT outcomes reported in our study.

In the trial, each patient took either a high-dose oral VD capsule, placebo capsule, or low-dose VD capsule prior to each study visit as described in **Fig. 1**. To assess the effects on serum VD levels, blood was sampled at each visit. As shown in **Table 1**, ∼80% of patients, 25-OH D3 serum levels were higher after taking high-dose VD, compared to the prior visit (at which no VD was taken). At their initial study visit, 70% of the Cleveland patients and 40% of the Arizona patients were Vitamin D deficient per our hospital’s standard criteria. Those percentages dropped to 21% and 14%, respectively, by the end of the trial, indicating excellent compliance by our participants when taking their assigned study drugs.

To assess our primary endpoint, we examined the kinetics of tumor regression after PDT with or without high-dose VD pretreatment. A positive VD response was defined as a situation in which neoadjuvant VD combined with PDT caused a faster rate of tumor shrinkage (downward sloping line segment labeled “a” in **Fig. 5A** or **5B**, for example), when compared to the PDT session preceded only by placebo (line segment labelled “b”). Conversely, an absence of VD response was determined if the slope of “a” was the same or greater than the slope of “b” (**Figs. 5 C, D**). A major challenge here was the requirement of having a visible tumor present at 4 study visits to perform a reliable VD kinetic analysis; in fact, a majority of BCCs in the study disappeared before Visit 4. In addition, many tumors from the Arizona dataset cleared more rapidly than we initially expected due to inflammatory effects from prior biopsy (see Discussion). In total only 49 tumors in 11 patients were suitable for kinetic analysis. Amongst the 11 patients, a majority of their tumors responded favorably to VD in 6 of 11 patients (**Supplementary Table S1**, column G). At first glance, this is not very convincing for a meaningful VD effect. However, realizing that a significant proportion of the 49 lesions were thick, non-PDT-responsive tumors that were unlikely to yield any observable effect even with VD present, we performed a stratified analysis; tumors were grouped into “thick” versus “thin” tumors based upon a 3DAvHt threshold (similar to **Fig. 3D**).

**Fig. 5.**
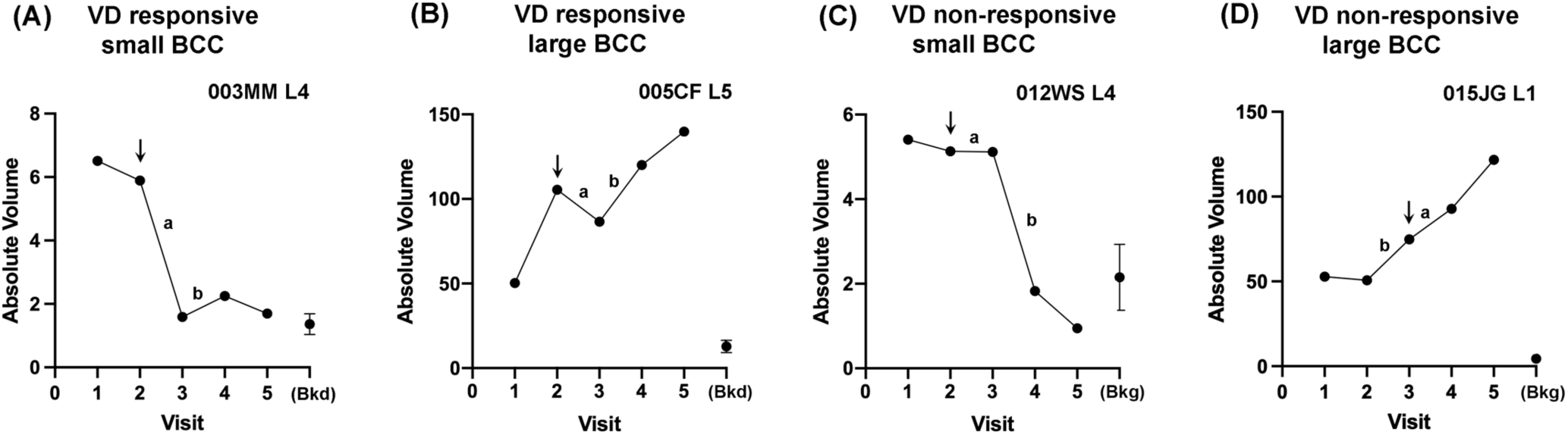
Examples of the changes in BCC tumor volume kinetics used to define Vitamin D-induced PDT responsiveness. *Arrows*, study visit at which patients received neoadjuvant high-dose VD prior to PDT. The slopes of the line segments labelled “a” (occurring after high-dose VD) or “b” (occurring after placebo) were compared; if segment “a” was smaller (more negative) than segment “b”, then the tumor was scored as VD-responsive.

Results shown in **Supplementary Table S4** (Supp8) indicated that for thick tumors, only 28% were responsive to VD compared to 72% that were nonresponsive. In contrast for thin tumors, 65% were responsive to VD. This provides reasonable evidence that neoadjuvant VD may indeed be beneficial as an accelerant of PDT-induced clearance in thin BCC tumors.

## DISCUSSION

Adoption of PDT as an accepted treatment for BCC has been slow in the United States, due to continuing uncertainty about which tumors are likely to respond to PDT versus those likely to persist. In this clinical trial, we systematically examined 122 BCC tumors in 35 patients to ask how (1) tumor size, (2) histologic subtype, and (3) pretreatment with high-dose vitamin D might influence the therapeutic response to PDT. For each question, we report new findings and insights.

In terms of size, we show that tumor volumes and average heights calculated by 3D photographic morphometry can define a size threshold for PDT efficacy which discriminates between PDT-responsive and PDT-resistant tumors (**Fig. 3D**). Because this noninvasive technique provides a surrogate measure of actual tumor size, predicts therapeutic responsiveness, and is relatively easily to perform, it should be helpful in dermatology clinics to identify which BCC are appropriate candidates for PDT. The calculated parameter, 3DAvHt, is roughly correlated with the depth of BCC tumor nests below the surface, even while representing only ∼10-20% of the actual tumor depth (**Fig. 3E**). The threshold value (3DAvHt = ∼0.15 – 0.20 mm) (**Fig. 3D**) corresponds to an approximate tumor depth of 1.5 mm (**Fig. 3F**), which matches quite closely to results from a clinical study by Mosterd in 2008 that comparing PDT outcomes to BCC histological tumor depth (*23*). In that study, PDT responses of 78 BCC tumors with a wide range of thickness values (0.30–3.10 mm) were categorized into two groups with widely different outcomes, i.e., tumors < 1.3 mm thick had a risk of treatment failure of 15.5%, whereas tumors > 1.3 mm thick had a risk of treatment failure of 42.2% % (P = 0.09). Their 1.3 mm depth threshold (*23*) and our estimated 1.5 mm threshold are very similar. Importantly, these values are consistent with European guidelines recommending that PDT be performed only for BCC < 2 mm in depth (*24,25*). Our new 3D photographic method represents an objective, noninvasive way of judging whether a BCC tumor falls within these established clinical guidelines.

The recommendation of 2 mm as a depth threshold originated with a pioneering study by Morton of 53 BCC tumors which demonstrated that thickness significantly affected PDT response, with no response in 4 patients whose tumors exceeded 2 mm in thickness (*26*). Subsequently, most PDT clinical trials have focused on patients with BCC tumors < 2 mm thick, primarily superficial BCC but some with nodular BCC as well (*27*). As a result, many clinicians who perform PDT have learned to visually estimate tumor depth with a surprising degree of accuracy; but the ability to correctly predict the histological BCC subtype is relatively poor, i.e., only 72% in a recent large study (*28*). Surface diameter can be indicative of BCC depth, but only for large tumors. In 235 biopsies of relatively large nodular BCC (mean surface diameter 13.0 ± 8 mm), Takenouchi et al. found that horizontal surface diameter correlated with and was the strongest predictor of tumor depth, followed by histologic subtype (*29*). Our new 3D morphometry technique permits a relative depth evaluation of any BCC tumor, whether large or small, which could be quite helpful for PDT screening purposes. That said, any BCC tumor that is large or has clinically suspicious features should be histologically evaluated before PDT.

An important and fascinating question is how 3DAvHt is able to predict PDT responses when most of the tumor lies below the surface. Here, we need to consider the concept of “phodynamic priming”, in which only a portion of a tumor needs to be damaged by PDT to trigger local events (expression of DAMPs, alteration of tumor stroma, stimulation of innate immune cells) that eventually lead to activation of systemic anti-tumor immunity (*30–33*). Light penetration into the skin is controlled by the laws of physics, and therefore photodynamic efficiency of light at various depths within the skin can be modeled and predicted (*34*). Combining this idea with the concept of photodynamic priming, we postulate that 3DAvHt (a parameter proportionately related to histological tumor depth) is useful because it correlates with the light penetration depth required to successfully induce photodynamic priming.

The above discussion about tumor thickness should not obscure the fact that BCC histological subtype is an overriding determinant of PDT responsiveness. Four BCC growth patterns are thought to have prognostic significance post-treatment: superficial, nodular, micronodular, and infiltrative (*35*). In a study of BCC skin biopsies by Welsch et al. (*36*), superficial (23%) and nodular (59%) subtypes were most prevalent, while micronodular (13%) and infiltrative (5%) subtypes were less common. This compares well with our data in which the distribution of subtypes was superficial (31%), nodular (44%), micronodular (15%), infiltrative (5%), and other (7%). The rank order of mean tumor depths in the Welsch study (*36*) was reported as micronodular (2.01 mm) > infiltrative (1.82 mm) > nodular (1.68 mm) > superficial (0.71 mm), very similar to the rank order in our patients: micronodular (1.26 mm) = infiltrative (1.56 mm) > nodular (0.77 mm) > superficial (0.13 mm). Micronodular and infiltrative subtypes are often reported as the thickest tumors and the most likely to recur, and indeed these were the deepest and most frequent histologic subtypes in our dataset (**Supplemental Table S2, Graph A**).

Now, let us consider how BCC histological subtype affects response to PDT. The most extensive studies have been done with sBCC, and show tumor clearance rates of 95%-100% between 3 months and 1 year (*12,37,38*), falling to 76%-88% at 5 years (*10,11*). Overall results with nBCC are somewhat worse (*39,40*). Thus, in terms of pre-selection of BCC lesions for PDT, the best responses will be achieved with sBCC tumors but excellent results can also be obtained for nBCC if tumors with 3DAvHt < 0.15 mm are selected. On the other hand, micronodular, infiltrative, adenoid, or trichoepitheliomatous BCC are best treated with surgery.

In our study, we compared results from BCC that were biopsy proven versus those that had not undergone biopsy. Amongst 70 biopsy-proven lesions in the Arizona patient cohort, 53% showed no evidence of clinical disease by Visit 2 (before any PDT), whereas patients without any pre-enrollment biopsy showed no lesion disappearance at Visit 2; see **Supplementary Table S5** (Supp 9). This finding suggests the possibility of an anti-tumor immune response triggered after surgical biopsy. Previous studies have shown that over 20% of tumors undergoing Mohs surgery reveal no remaining tumor on intraoperative examination (*41*). Conversely, in patients presenting with no clinically apparent BCC tumor after biopsy, residual tumor may still be found in over 80 % of cases (*42*). To ask whether rates of tumor clearance after PDT were faster in patients with a pre-enrollment biopsy, the percentages of tumor clearance in two groups (biopsy versus no biopsy) were compared (**Supplementary Table S5**). However, no evidence for acceleration in tumor clearance after PDT at Visits 3, 4, and 5 in the biopsy group was found, suggesting that inflammatory or immunological effects that promote tumor clearance post-surgery must be relatively short-lived.

The third question addressed by this trial was whether Vitamin D modifies the therapeutic effects of PDT. The scientific rationale for this idea is strong. Preclinical work in mice with SCC (*14,18*) and recent clinical trials in patients with squamous pre-cancers demonstrated that neoadjuvant VD has a significant positive effect upon PDT response (*15,16,43*). Originally, we attributed this to the fact that VD promotes selective accumulation of PpIX within tumor cells, driving the improved therapeutic response (*14,18*). However, recent evidence in a mouse model of UV-induced squamous precancer revealed significant involvement of the immune system, with striking increases in neutrophils, macrophages, and T-cells into neoplastic lesions within the first few days after PDT (*44*); this immunocyte recruitment was greatly enhanced by pretreatment with VD (*45*). We anticipate that similar mechanisms are at play in BCC, but this remains to be investigated. However, results of the current clinical trial demonstrate that neoadjuvant VD does indeed hasten lesion shrinkage rates for sBCC, and for nBCC tumors thin enough to respond to PDT (although VD does not convert highly PDT-resistant tumors into PDT-sensitive ones). Given the excellent safety profile of vitamin D3, there appears to be a clear potential benefit and no downside of using oral VD together with PDT for BCC.

Most effects of VD are mediated by the Vitamin D receptor (VDR), which can be affected by single-nucleotide polymorphisms (SNPs) within the VDR gene. In a previous study, we had demonstrated a strong association between two of these SNPs (Fok1 and Poly-A) alleles and the development of actinic keratoses and squamous cell carcinoma, and also between the SNPs and 25-OH-D3 serum levels (*21*). Specifically, the presence of Poly-A homozygous short alleles (SS) and Fok1 homozygous dominant alleles (FF) were associated with high levels of serum 25-OH-D3 (*21*). In the current work on BCC, we were intrigued to find a possible involvement of the Poly-A (SS) and Fok1 (FF) alleles in tumor clearance. Patients with 100% tumor clearance in **Supplementary Table S6** (Supp 10) featured primarily a Poly-A (SS) or a Fok1 (FF) genotype, whereas the heterozygous variants had a less complete therapeutic response. This observation, while still preliminary due to the small sample size, is nonetheless interesting because VDR proteins encoded by the Poly-A (SS) and/or Fok1 (FF) gene variants are thought to have a higher-than-average functional activity in cell-based reporter assays designed to measure transcriptional activity of the VDR (discussed in (*21*)). Exactly how different levels of VDR expression might influence BCC growth and involution rates after PDT remains an open question but it may involve effects on the immune system as well as on the tumor itself.

This clinical trial had several limitations. First, the number of patients available for analysis of vitamin D effects was limited, because for comparative kinetics it was necessary to have a visible BCC tumor to observe at 4 or more consecutive visits. During trial planning, we had not anticipated how well PDT would work; 58% of BCC lesions disappeared by Visit 4, leaving only 42% of lesions to analyze. Another limitation was the strong effect of pre-enrollment biopsy upon BCC tumor behavior, such that ∼21% of lesions were already gone by Visit 2 (although the effect is compatible with previous studies, see above). A third issue was that the trial design, as a practical necessity, did not require all lesions to be biopsy-proven. Consequently, of the 211 lesions chosen for monitoring, ∼20% were clinically misdiagnosed and later proved to be either scars, keloids, or SCC.

In summary, the following conclusions can be drawn from this study: *First*, our discovery of a noninvasive way to measure tumor depth, which reflects actual histological depth and predicts tumor responsiveness to PDT, provides a new tool to help reduce uncertainty when choosing which BCC lesions to treat with PDT. *Second*, our data confirm that proper histological categorization is critical to PDT response. Thin nodular BCC tumors respond quite well, but aggressive subtypes (micronodular, infiltrative, adenoid, or trichoepitheliomatous) do not respond to PDT and are best treated with surgery. The combination of histological assessment to exclude aggressive BCC subtypes, combined with 3D photographic screening for appropriate thickness (3DAvHt < 0.2 mm), should make it possible to personalize PDT treatment and maximize favorable results. *Third,* addition of neoadjuvant oral VD is a safe and effective technique to augment PDT-induced therapeutic responses. *Finally,* it is our hope that this report will stimulate optimism about the prospects of using PDT for basal cell carcinoma by making physicians aware that good results are achievable when tumor thickness and subtype are taken into account.

## Data Availability

All data produced in the present work are contained in the manuscript

