## Supplementary Methods 1 for "A Clinical Trial to Determine the Impact of Tumor Size, Histological Subtype, and Vitamin D Status on the Therapeutic Response of Basal Cell Carcinoma to Photodynamic Therapy"

**SUPPLEMENTARY METHODS 1. Analysis of BCC tumor size using 3D software from Quantificare**

**Description of 3D camera and software:**

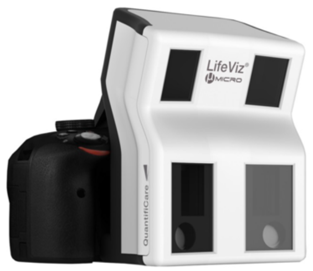

**Fig. A.** The LifeViz Micro camera

The LifeViz Micro camera by Quantificare, Inc. (www.quantificare.com) consists of a standard Nikon digital camera body outfitted with a special housing that contains mirrors and dual lenses spaced ~5 cm apart, in order to simulate stereo vision (**Fig. A**). With this camera, the researcher simultaneously captures a pair of photos of the tumor which are stored digitally. These dual images are then transferred to a laptop computer running Quantificare's DermaPix software (3D image analysis program). After importing the pair of images into the Dermapix analysis program, the pair of images of the tumor are displayed side-by-side in 2-dimensional mode (**Fig. B**). The operator defines the area of interest (ROI) to be analyzed by drawing an outline around the tumor, shown in green in this example. One can toggle back and forth between the 2D drawing mode (**Fig. B**) and 3D visualization mode in which the tumor is displayed as a 3D image (**Fig. C**). The 3D image can be rotated in space and viewed from all angles. It can also be magnified to show very fine details such as skin lines and small ulcerated areas within the tumor, as seen in **Fig. C**.

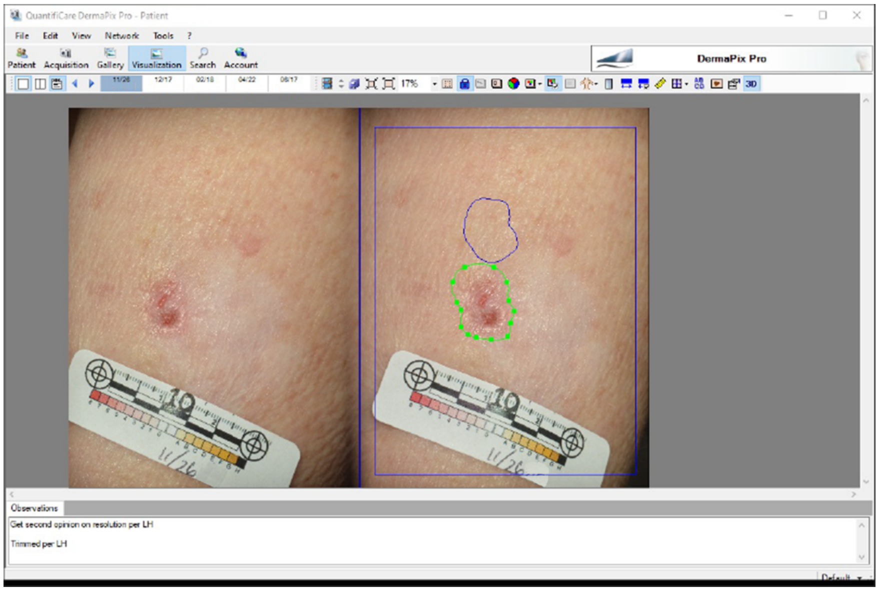


**Fig. B.** Appearance of the pair of images after being imported into the Dermapix software program. Images are displayed here in 2D mode.

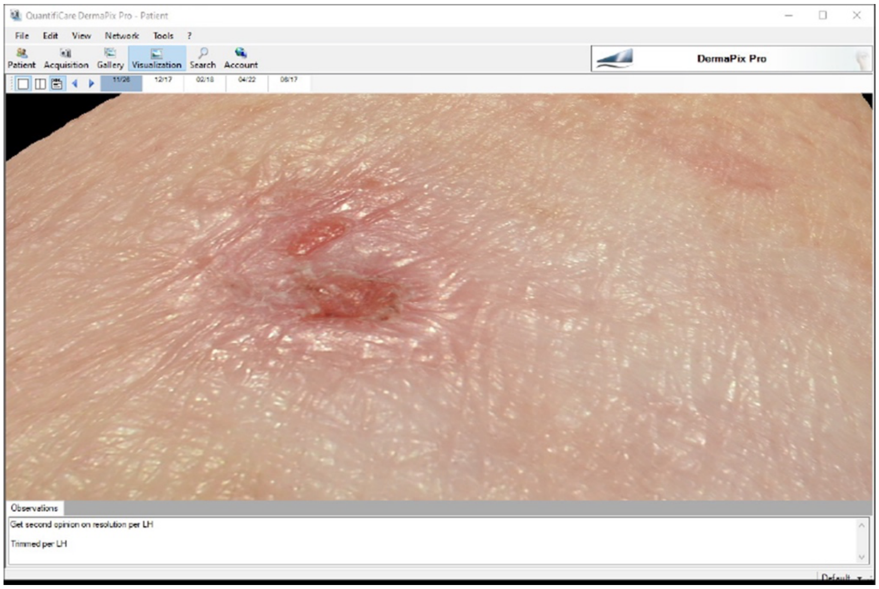


**Fig. C.** The same BCC tumor, now visualized in 3D mode. Note the raised edges, central ulceration, and disruption of normal skin lines.

### Analysis of tumors using the Quantificare 3D camera and Dermapix software:

To perform quantitative analysis, one asks the computer program to convert the 3D image into a 3D mesh network from which various measurements within the ROI can be calculated. In the example shown in **Fig. D**, the ROI is defined as the black outline drawn around the tumor (window on the left.) Within this ROI, various parameters are measured and reported within the report window (lower right). The parameters include *surface area*; *diameter* (of major axis); *perpendicular diameter* (of minor axis); *positive volume* above the surface plane; (iii) *negative volume* below the surface plane; *maximum lesion height*; *average lesion height*; and '*roughness*'. In developing our data, we empirically tried using each parameter for analyses, and ultimately found two of them to be the most consistent and useful for describing changes in tumor size after PDT. The first of these is *Absolute Volume* ("**AbsVol"**), defined as the positive volume plus the absolute value of the negative volume. The second parameter is *Average Lesion Height*, which we abbreviate in the manuscript as "**3D AvHt**". From the report window, parameters of interest are exported to an Excel spreadsheet for further analysis as desired.

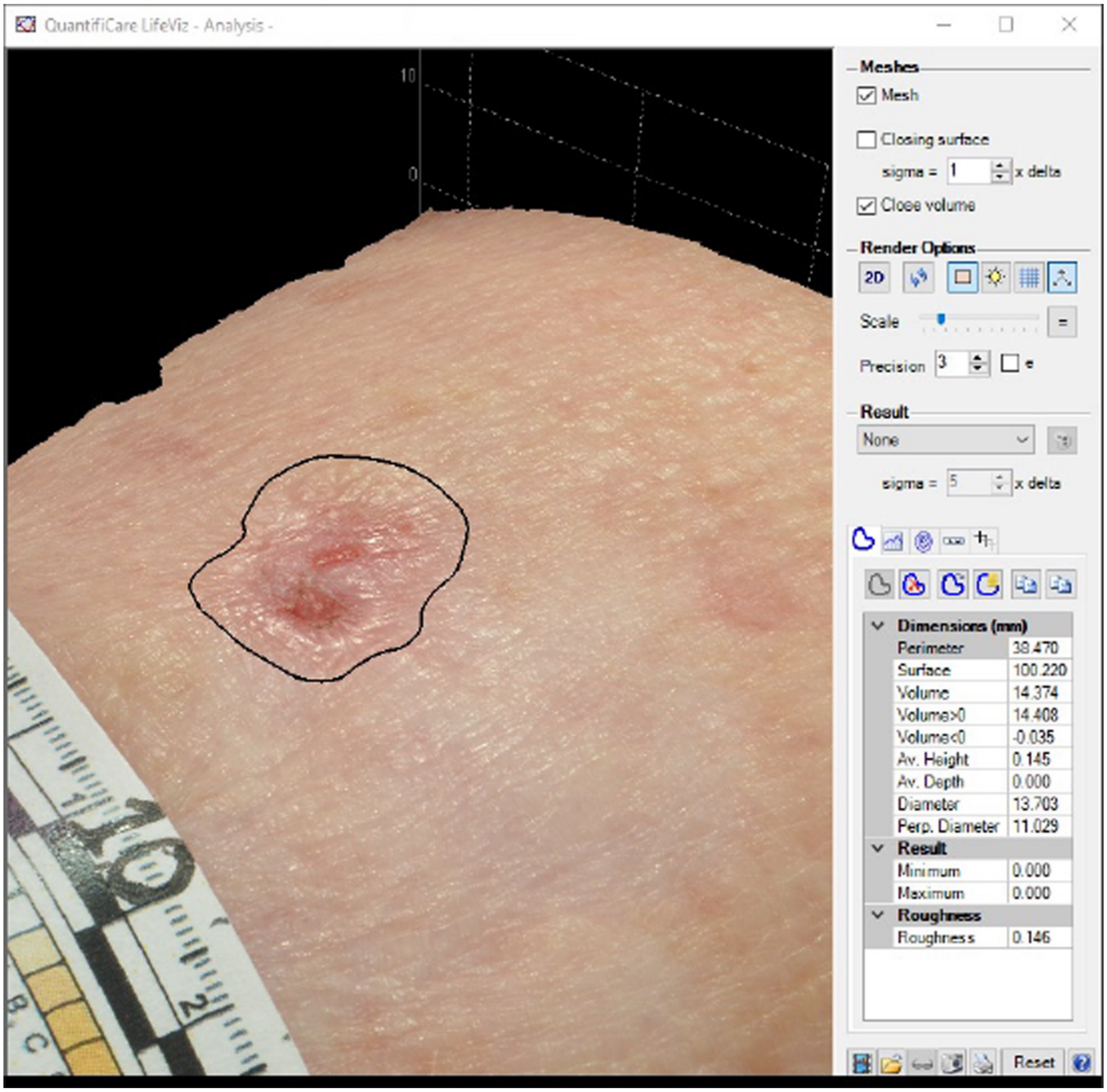


**Fig. D**. Appearance of lesion 3D Analysis Window. On the left is the tumor image after it has been converted into a 3D mesh network to allow calculation of tumor size parameters. (N.B. The white sticker with the 1-cm ruler markers and the lesion number "10" written in ink was placed on the patient's skin when the photograph was taken). On the right, one can see the portion of the 3D analysis window that allows the experimentalist to alter how the tumor is displayed and to select certain measurement thresholds. Once the analysis has been run, the results are shown in the report window on the lower right. These data can be imported directly into a Microsoft Excel spreadsheet.

### Calibration of 3D absolute volumes using known size standards:

The 3D analysis system (3D camera, and associated software) is pre-calibrated at the factory to generate numerical outputs in millimeter units (for lengths), and other calculated parameters in either $mm^2$ (for areas) or $mm^3$ (for volumes). However, due to the complexity of 3D mesh calculations, some inaccuracy is expected. To estimate the true relationship between tumor volume and the measured AbsVol, the following experiment was conducted. First, BCC tumor phantoms were created as follows. Decorative beads, made of epoxy resin and shaped as half-spheres of various sizes, were purchased from a local hobby store. Each bead was weighed on a laboratory balance (Mettler), and then adhered to a piece of cardboard to create a BCC tumor "phantom". Each of the beads was photographed using the Quantificare 3D camera, and their AbsVol were calculated by the DermaPix software as illustrated in **Fig. E**.

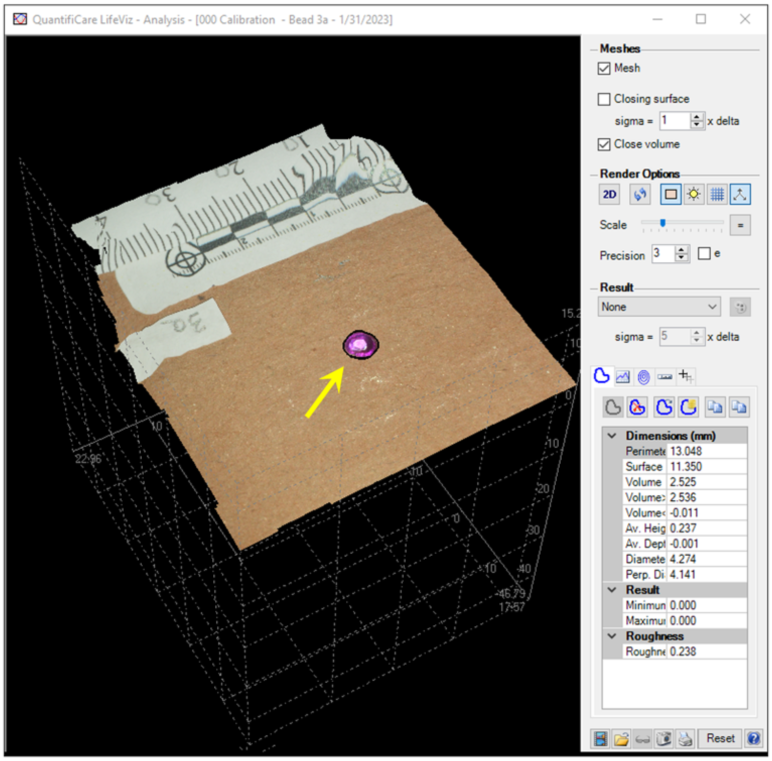


**Fig. E**. Tumor phantom (yellow arrow) visualized in the 3D analysis window.

The calculated AbsVol values were plotted against the actual volumes of the standards, assuming the density of epoxy to be 1.2 mg/mL (or 1.2 $mg/mm^3$). The plot in **Fig. F** demonstrates a linear relationship between AbsVol and the actual volume. The AbsVol calculated by DermaPix for our setup appears to be ~50% less than the actual volume of the tumor phantoms.

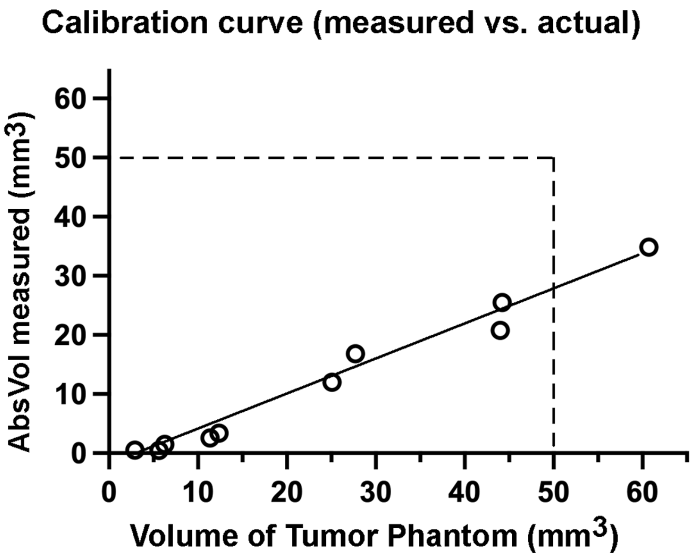


**Fig. F**. Calibration curve, showing values of *AbsVol* versus the *actual volume* of tumor phantoms.
