## Supplementary Methods 2 for "A Clinical Trial to Determine the Impact of Tumor Size, Histological Subtype, and Vitamin D Status on the Therapeutic Response of Basal Cell Carcinoma to Photodynamic Therapy"

### SUPPLEMENTARY METHODS 2. Remote digital information management for the clinical trial

#### The need for digital technology to improve the conduct and management of clinical trials.

In clinical research, two major challenges encountered by investigators are (1) how to identify and recruit patients, and (2) how to maintain tight communication with all study participants, including the patients and all team members throughout the duration of the study so that no information is lost. Up until recently, the major tools available to clinical investigators were printed advertisements (for patient recruitment) and telephone calls (for follow-up). For Dr. Maytin's clinical trial, a particular challenge was reaching a population of patients with a rare condition called Gorlin Syndrome, in which patients develop numerous skin cancers (basal cell carcinomas) beginning in childhood and typically progressing in adulthood so that the patient ultimately develops dozens or hundreds of tumors. The condition is very rare (1/30,000 people), presenting the study team with a serious challenge of how to identify patients with the syndrome and inform them about the existence of our clinical trial. To help with this, we established two study sites at opposite ends of the country, to reduce travel distance barriers. However, this was only a partial solution, and it highlighted another challenge which was how to share clinical information and tissue samples securely and efficiently between the two sites.

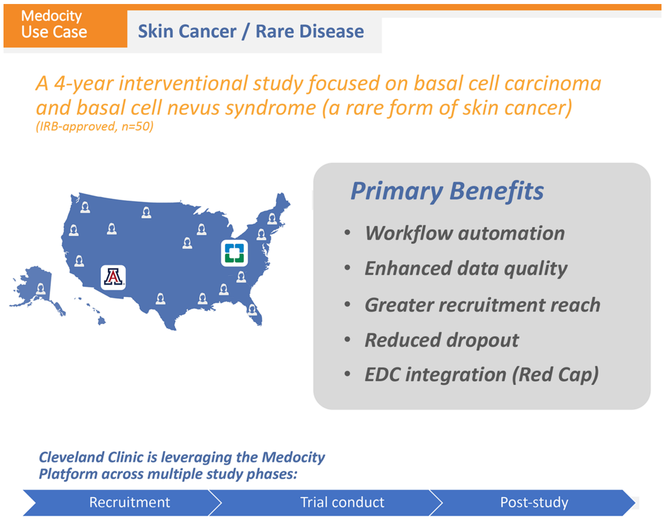


**Fig. A**. The intent of a web-based clinical trial platform is to improve recruitment, enhance data quality, reduce dropout, and allow data integration and data sharing.

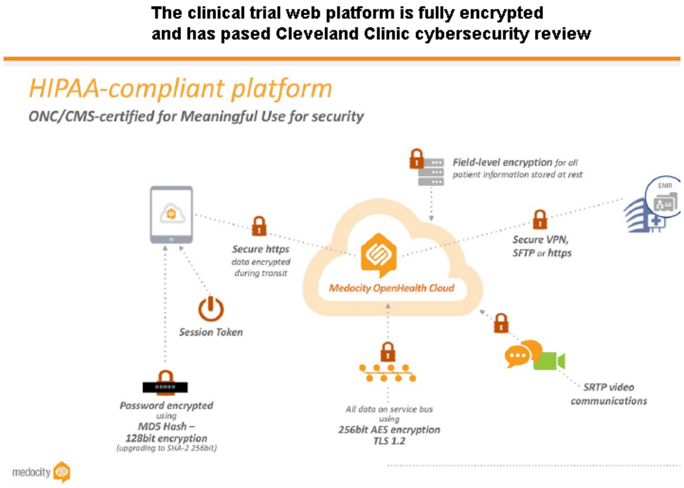


**Fig. B**. The online clinical trials platform from Medocity was password protected and fully encrypted.

#### Collaboration with Medocity for the tailored use of a web-based clinical trials platform.

To overcome these hurdles, Dr. Maytin's team reached out in 2019 to an innovative web-based software company (Medocity, Inc., located in Parsipanny, NJ) that was developing an online platform for communication between patients and healthcare providers. At the time, the company was piloting an innovative web-based application that remains highly novel in the field of clinical research. Their approach offers exceptionally broad versatility as it is compatible with all smart phones (Apple or Android) and with all desktop-based computers (PC and Mac). Dr. Maytin's team entered into a formal collaboration with the software developers at Medocity to configure their online platform to fit our study's needs and to help guide the patients' journey during the clinical trial from beginning to end (**Fig. A**). The collaboration required full IRB review at Cleveland Clinic, including proof that the system is HIPAA-compliant and cybersecure (**Fig. B**). Capabilities provided by the web platform include the following important aspects. (1) Recruitment: Patients who heard about the study from a Google search online or from other types of advertisement could easily access the study-specific web portal to obtain further information by reading about the basic goals and objectives of the study, and to learn about the study sites and personnel (**Fig. C**). If interested, they could fill out a brief online questionnaire to determine whether they might be eligible for the study (**Fig. D**), and request a call-back from our study coordinator for further details. Prospective patients anywhere in the country were able to upload photographs and biopsy reports securely, which the study coordinator could evaluate for potential enrollment eligibility. A telehealth online visit could be arranged if desired. Overall this remote screening feature allowed us to determine patient eligibility without the need for a trip to Cleveland or Arizona. (2) <u>Study visit scheduling and follow-up</u>. Once a patient was enrolled, the web-based software allowed investigators to communicate regularly with patients and send reminders. Patients could view a calendar with their upcoming appointments and receive real-time notifications each time they needed to take their study medication. After every PDT treatment, patients recorded their symptoms and side effects directly in the web portal, thereby enhancing the reliability of the data and reducing the chance for recall errors. Use of the online platform reduced the need for study team members to spend time on the phone performing follow-ups and recording side effects. (3) <u>Secure inter-site communication and transfer of data</u>. The website provided a secure, centralized record-keeping tool. For example, hundreds of digital 3-D photographs were taken at the two sites during the clinical trial, and because the final 3-D software analyses were conducted by the Cleveland team, it was crucial for each team to be able to upload and share the photos along with the associated clinical data. For us, the ability to share confidential clinical information in a secure, password-protected environment was a critical feature of the Medocity platform.

In summary, the use of this digital web-based patient-doctor communication platform allowed us to extend our national outreach for recruitment, improve communication and reliability, and make the research study as convenient as possible for our patients.

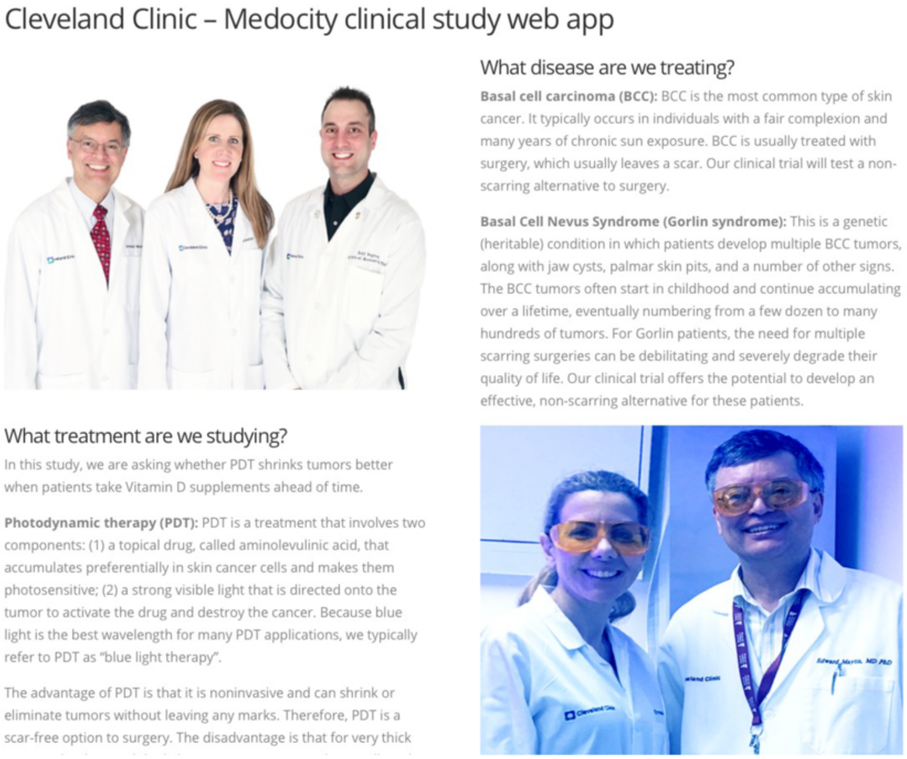


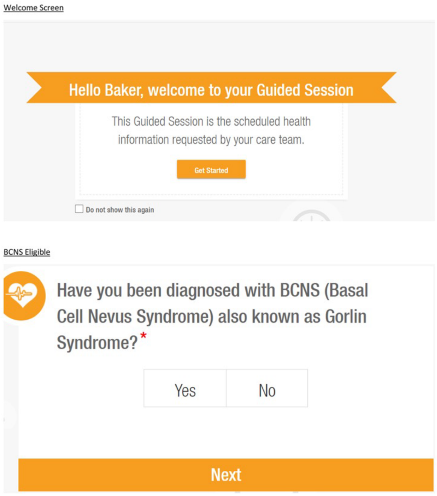


**Fig. C**. Screenshot of a patient-facing informational page on the website.

**Fig. D**. Example of one of the pages presented to patients when filling out the online recruitment questionnarie.
