## Supplementary Table S1 for "A Clinical Trial to Determine the Impact of Tumor Size, Histological Subtype, and Vitamin D Status on the Therapeutic Response of Basal Cell Carcinoma to Photodynamic Therapy"

**SUPPLEMENTARY TABLE S1 Complete Data from Clinical Trial NCT03483441; Responses of Basal Cell Carcinoma to Photodynamic Therapy. Lesional 3D analysis of BCC tumor size changes, responsiveness to Vitamin D, and histological characterization.**

***Explanation of information in each column of the table:***

| | | |
|---|---|---|
| A | **Index Number:** | This is a lesion index number, i.e., a designation for every unique lesion that was photographed at the first study visit. (Note that not every lesion was ultimately shown to be a BCC suitable for 3-D photographic analysis). |
| A | **Subject Number:** | The study I.D. number assigned to each patient in the clinical trial. |
| B | **Lesion Number:** | A number assigned to each suspected BCC lesion (up 10 lesions) under study in any given patient. |
| C | **Body Location:** | The location of the BCC lesion on the body. |
| E | **Diagnosis by Biopsy:** | The histological diagnosis, made by a Board-certified dermatopathologist and listed on the official pathology report. In Cleveland (Gorlins patients), diagnosis of BCC was determined from clinical features; only lesions that failed to clear were biopsied. In Arizona site (chronic UV-exposed patients), patients needed a minimum of three biopsy-proven tumors prior to enrollment. |
| F | **Graphical data:** | Graphs of absolute tumor volume, plotted from yellow-highlighted in the tables to the right. The downward arrows in some graphs indicate the Visit at which high-dose oral Vitamin D3 (10,000 IU) was administered. |
| G | **Response to VitD:** | For each patient, shown at top (blue box) is the randomized assignment for high-dose VitD, i.e., whether given prior to Visit 2 (V2) or Visit 3 (V3). Other text in the column, "VitD=Yes" or "VitD=No", compares the rates of change in tumor size after (PDT+VitD) versus after (PDT+placebo). For tumors that decreased after each PDT treatment, "VitD = Yes" indicates that the downward slope was steeper following (PDT+VitD) than after (PDT+placebo). Lesions that were already gone at V3 could not be used for this analysis. |
| H | **3-D lesion data:** | These are the values generated by 3-D Quantificare Dermapix analysis of the lesion at each study visit. (See manuscript text for details) For every lesion, three background measurements for 3D absolute volume averaged from areas of normal skin (as shown at the right). The average background value is the rightmost data point shown in each of the 3D AbsVol graphs. The tumor dimensions estimated from clinical exam (diameter, height in millimeters) were done for Cleveland patients only. |
| I | **Visit Number:** | Each indicates the study visits at which 3D photographic measurements were taken. |
| J | **Response outcome:** | By comparing 3D AbsVol data (highlighted yellow) with average background from the same lesion, the first visit at which the tumor volume was statistically similar to background was **defined as tumor clearance** and is reported in the orange box as either **V3, V4, or V5**. Tumors that were PDT-resistant and failed to disappear by Visit 5 are termed **"NC" (Not cleared).** |
| K | **VDR gene:** | For about half of the patients, the genotypes of allelic variants at two sites in the Vitamin D Receptor (VDR) gene were determined. These alleles are designated in the blue boxes as "F" versus "f" (for the Fok1 allele), and "Long" versus "Short" (for the Poly A allele). |

### CLEVELAND PATIENTS:

| A | B | C | D | E | F | G | H | I | | | | | J | K |
|---|---|---|---|---|---|---|---|---|---|---|---|---|---|---|
| *Lesion Index #* | Subject # | Lesion # | Body Location | Diagnosis by Biopsy | Graphical data | Notes & Response to VitD | 3-D lesion data | Visit 1 | Visit 2 | Visit 3 | Visit 4 | Visit 5 | Response outcome | VDR gene |
| | | | | | | **High-dose Vit D assignment: before V3** | | Lengths are in (mm); Volumes in (mm^3) | | | | | | **VDR alleles for patient #1** |
| | | | | | | | | | | | | | | Ff \| LS |

#### Lesion 1

| Lesion Index # | Subject # | Lesion # | Body Location | Diagnosis by Biopsy |
|---|---|---|---|---|
| *1* | 1 | 1 | scalp | Biopsy at Visit 5: Prurigo nodule (no tumor) |

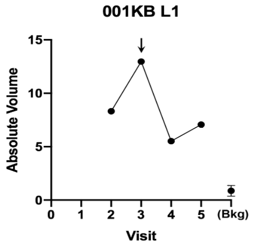

VitD = Yes

Big lesion in scalp.
Still a raised nodule at V5.

| 001KB L1 | V1 | V2 | V3 | V4 | V5 |
|---|---|---|---|---|---|
| Height (exam) | Raised | Raised | raised | raised | Flat |
| Diameter (exam) | 13 x 5 | 20 x 9 | 20 x 9 | 16 x 8 | 14 x 10 |
| 3D diameter | | 18.68 | 21.26 | 17.77 | 16.64 |
| 3D perp diameter | | 11.51 | 12.52 | 10.86 | 10.50 |
| 3D av height | Photo | 0.044 | 0.024 | 0.004 | 0.024 |
| 3D Av Ht/Area * 10^4 | missing | 0.613 | 0.263 | 0.059 | 0.420 |
| 3D volume | | 4.171 | -4.269 | -4.948 | -1.640 |
| 3D absol volume | | 8.314 | 12.975 | 5.522 | 7.069 |

Response outcome: **V5**

Background region at visit: **Average bkdg:**

| V1 | V2 | V5 | MEAN | SD |
|---|---|---|---|---|
| 0.708 | 0.886 | 1.026 | **0.873** | 0.159 |

#### Lesion 2

| Lesion Index # | Subject # | Lesion # | Body Location | Diagnosis by Biopsy |
|---|---|---|---|---|
| *2* | 1 | 2 | temple | Biopsy at Visit 5: Folliculitis (no tumor) |

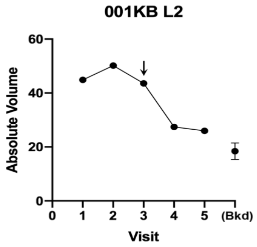

VitD = Yes

Thin BCC near hairline.
Still raised at V5, so we biopsied it, but tumor had resolved.

| 001KB L2 | V1 | V2 | V3 | V4 | V5 |
|---|---|---|---|---|---|
| Height (exam) | raised | raised | raised | raised | Flat |
| Diameter (exam) | 15 x 11 | 20 x 11 | 20 x 11 | 10 x 10 | 10 x 10 |
| 3D diameter | 20.03 | 24.16 | n/d | 15.79 | 14.60 |
| 3D perp diameter | 17.64 | 17.71 | n/d | 15.05 | 12.79 |
| 3D av height | 0.058 | 0.198 | n/d | 0.170 | 0.190 |
| 3D Av Ht/Area * 10^4 | 0.524 | 1.441 | n/d | 2.274 | 3.218 |
| 3D volume | -15.407 | 46.572 | 41.975 | 26.019 | 25.502 |
| 3D absol volume | 44.856 | 50.176 | 43.591 | 27.413 | 25.959 |

Response outcome: **V4**

Background region at visit: **Average bkdg:**

| V1 | V2 | V5 | MEAN | SD |
|---|---|---|---|---|
| 20.734 | 14.957 | 19.544 | **18.411** | 3.050 |

#### Lesion 3

| Lesion Index # | Subject # | Lesion # | Body Location | Diagnosis by Biopsy |
|---|---|---|---|---|
| *3* | 1 | 3 | forehead | Biopsy at Visit 5: BCC, nodular & micronodular |

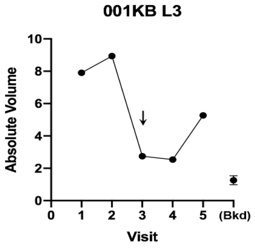

VitD=No

Nodular lesion, shrank then grew
Biopsy showed residual BCC

| 001KB L3 | V1 | V2 | V3 | V4 | V5 |
|---|---|---|---|---|---|
| Height (exam) | Raised | Raised | Raised | Raised | Raised |
| Diameter (exam) | 18 x 18 | 7 x 3 | 7 x 6 | 6 x 5 | 6 x 6 |
| 3D diameter | 7.56 | 8.08 | 7.80 | 5.91 | 6.32 |
| 3D perp diameter | 6.64 | 6.58 | 6.78 | 4.13 | 4.42 |
| 3D av height | 0.081 | 0.100 | 0.109 | 0.092 | 0.166 |
| 3D Av Ht/Area * 10^4 | 5.138 | 5.924 | 6.552 | 11.596 | 18.287 |
| 3D volume | 2.068 | 0.000 | 2.742 | 2.337 | 5.264 |
| 3D absol volume | 7.905 | 8.940 | 2.744 | 2.534 | 5.270 |

Response outcome: **NC**

Background region at visit: **Average bkdg:**

| V1 | V2 | V5 | MEAN | SD |
|---|---|---|---|---|
| 1.579 | 1.036 | 1.180 | **1.265** | 0.281 |

#### Lesion 4

| Lesion Index # | Subject # | Lesion # | Body Location | Diagnosis by Biopsy |
|---|---|---|---|---|
| *4* | 1 | 4 | back | |

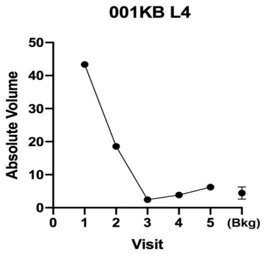

Visually gone by V5

| 001KB L4 | V1 | V2 | V3 | V4 | V5 |
|---|---|---|---|---|---|
| Height (exam) | Raised | Raised | Raised | Raised | invisible |
| Diameter (exam) | 11 x 11 | 7 x 5 | 7 x 5 | 4 x 3 | asurable |
| 3D diameter | 8.92 | 6.95 | 12.67 | 12.70 | 12.80 |
| 3D perp diameter | 7.32 | 4.58 | 9.06 | 9.05 | 8.96 |
| 3D av height | 0.071 | 0.052 | 0.055 | 0.034 | 0.070 |
| 3D Av Ht/Area * 10^4 | 3.430 | 4.992 | 1.482 | 0.915 | 1.880 |
| 3D volume | 42.885 | 18.217 | 4.580 | 2.940 | 5.900 |
| 3D absol volume | 43.334 | 18.573 | 4.920 | 3.848 | 6.241 |

Response outcome: **V3**

Background region at visit: **Average bkdg:**

| V1 | V2 | V5 | MEAN | SD |
|---|---|---|---|---|
| 4.615 | 2.512 | 6.241 | **4.456** | 1.870 |

5 | 1 | 5 | low back | Biopsy at Visit 5: BCC, nodular & micronodular

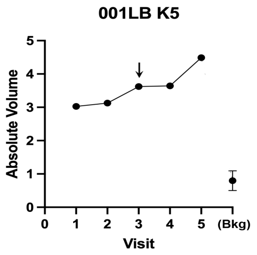

VitD = Yes

Slowed growth after VitD at V3

Biopsy showed residual BCC

| 001KB L5 | V1 | V2 | V3 | V4 | V5 |
|---|---|---|---|---|---|
| Height (exam) | Raised | Raised | Raised | Raised | Raised |
| Diameter (exam) | 6 x 6 | 10 x 5 | 10 x 5 | 4 x 5 | 5 x 4 |
| 3D diameter | 4.95 | 9.28 | 6.18 | 4.39 | 6.47 |
| 3D perp diameter | 4.25 | 6.27 | 4.03 | 4.09 | 4.98 |
| 3D av height | 0.058 | 0.060 | 0.067 | 0.102 | 0.167 |
| 3D Av Ht/Area * 10^4 | 8.771 | 3.169 | 8.196 | 18.014 | 16.266 |
| 3D volume | 2.095 | 2.165 | 3.522 | 3.613 | 4.395 |
| 3D absol volume | 3.029 | 3.127 | 3.625 | 3.645 | 4.488 |

NC

| Background region at visit: | | | Average bkdg: | |
|---|---|---|---|---|
| V1 | V2 | V5 | MEAN | SD |
| 1.140 | 0.601 | 0.656 | **0.799** | 0.297 |

6 | 1 | 6 | forearm

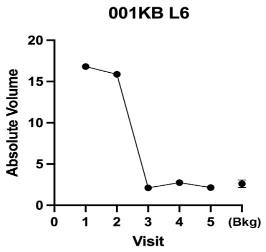

Visually gone by V3

| 001KB L6 | V1 | V2 | V3 | V4 | V5 |
|---|---|---|---|---|---|
| Height (exam) | Slightly R | Slightly ra | Slightly r | Slightly ra | Flat, almost invisible |
| Diameter (exam) | 6 x 6 | 5 x 4 | 5 x 4 | 10 x 8 | 10 x 8 |
| 3D diameter | 7.79 | 5.45 | 8.27 | 8.85 | 8.44 |
| 3D perp diameter | 7.03 | 4.82 | 7.22 | 7.23 | 6.99 |
| 3D av height | 0.047 | 0.039 | 0.051 | 0.056 | 0.051 |
| 3D Av Ht/Area * 10^4 | 2.707 | 4.667 | 2.706 | 2.760 | 2.730 |
| 3D volume | 16.790 | 15.670 | 2.035 | 2.561 | 2.055 |
| 3D absol volume | 16.801 | 15.882 | 2.102 | 2.744 | 2.145 |

V3

| Background region at visit: | | | Average bkdg: | |
|---|---|---|---|---|
| | | | MEAN | SD |
| 2.743 | 2.107 | 2.996 | **2.615** | 0.458 |

7 | 1 | 7 | arm

Lesion so small, doubt it is BCC.
DO NOT USE

| 001KB L7 | V1 | V2 | V3 | V4 | V5 |
|---|---|---|---|---|---|
| Diameter (exam) | 4x4 | 5x5 | 5x3 | 4x3 | Not measurable |
| Height (exam) | Slightly R | Slightly ra | Slightly r | Slightly ra | Flat |
| 3D volume | 0.6413 | 0.2897 | 0.9426 | 0.2381 | 0.1328 |
| 3D absol volume | 0.6413 | 0.29 | 1.029 | 0.38 | 0.3338 |

on V1 l on V2 on V5

8 | 1 | 8 | chest

Lesion at V3-5 not at same site.
DO NOT USE

| 001KB L8 | V1 | V2 | V3 | V4 | V5 |
|---|---|---|---|---|---|
| Diameter (exam) | 5x5 | 7x5 | 4x2 | 3x2 | 3x2 |
| Height (exam) | Flat | Flat | Flat | Flat | Flat |
| 3D absol volume | 0.6874 | 0.451 | 0.8157 | 0.5725 | 0.0462 |

on V1 l on V2 on V5

9 | 1 | 9 | chest

It all looks like folliculitis
DO NOT USE

| 001KB L9 | V1 | V2 | V3 | V4 | V5 |
|---|---|---|---|---|---|
| Diameter (exam) | 3x3 | 5x4 | 2x2 | 2x3 | Not measurable |
| Height (exam) | Raised | Raised | Raised | Raised | Flat |
| 3D volume | 0.6521 | 0.283 | 0.0543 | 0.8743 | |
| 3D absol volume | 0.6544 | 0.2846 | 0.0543 | 0.8743 | |

on V1 l on V2 on V5

10 | 1 | 10 | chest

Different areas pictured in all the visits. Not sure which one is L10.
It all looks like folliculitis
DO NOT USE

| 001KB L10 | V1 | V2 | V3 | V4 | V5 |
|---|---|---|---|---|---|
| Diameter (exam) | 5x5 | 9x4 | 2x3 | 4x5 | Not measurable |
| Height (exam) | Slightly ra | Slightly ra | Slightly r | Slightly ra | Flat |
| 3D volume | 0.6773 | 3.6401 | 0.8471 | | |
| 3D absol volume | 0.6922 | 3.6414 | 0.8471 | | |

on V1 l on V2 on V5

| *Lesion Index #* | Subject # | Lesion # | Body Location | Diagnosis by Biopsy | Graphical data | Notes & Response to VitD | 3-D lesion data | Visit 1 | Visit 2 | Visit 3 | Visit 4 | Visit 5 | Response outcome | VDR gene |
|---|---|---|---|---|---|---|---|---|---|---|---|---|---|---|

**High-dose Vit D assignment: before V3**

Lengths are in (mm); Volumes in (mm^3)

**VDR alleles for patient #2**: FF | LS

*11* | **2** | **1** | hairline

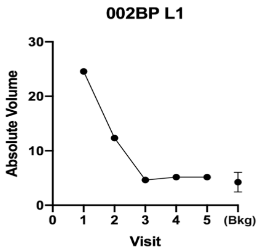

Lesion at hairline

| 002BP L1 | V1 | V2 | V3 | V4 | V5 |
|---|---|---|---|---|---|
| Height (exam) | Raised | n/d | Raised | Flat | ormal skin |
| Diameter (exam) | 15 x 9 | n/d | 15 x 9 | 15 x 9 | visually |
| 3D diameter | 11.42 | 10.12 | 13.65 | 16.10 | n/d |
| 3D perp diameter | 8.81 | 8.69 | 10.41 | 10.13 | n/d |
| 3D av height | 0.392 | 0.017 | 0.022 | 0.150 | n/d |
| 3D Av Ht/Area * 10^4 | 12.191 | 0.604 | 0.492 | 2.767 | n/d |
| 3D volume | 24.101 | -12.009 | 0.236 | 4.732 | n/d |
| 3D absol volume | 24.558 | 12.326 | 4.636 | 5.160 | 4.228 |

Response outcome: **V3**

| Background region at visit: | | | **Average bkdg:** | |
|---|---|---|---|---|
| V1 | V2 | V5 | MEAN | SD |
| 2.877 | 6.300 | 3.508 | **4.228** | 1.821 |

*12* | **2** | **2** | forehead

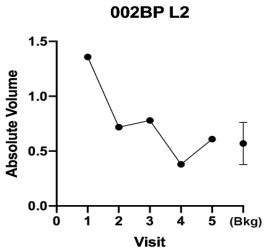

Small lesion at hairline

VitD = Yes

| 002BP L2 | V1 | V2 | V3 | V4 | V5 |
|---|---|---|---|---|---|
| Height (exam) | Raised | n/d | Flat | Flat | mal skin |
| Diameter (exam) | 6 x 4 | n/d | 4 x 4 | 4 x 4 | asurable |
| 3D diameter | 5.35 | 4.71 | lo lesion | No lesion | lo lesion |
| 3D perp diameter | 3.03 | 3.26 | visible | visible | visible |
| 3D av height | 0.047 | 0.016 | -- | -- | -- |
| 3D Av Ht/Area * 10^4 | 8.443 | 3.310 | -- | -- | -- |
| 3D volume | 0.43 | 0.04 | -- | -- | -- |
| 3D absol volume | 1.358 | 0.719 | 0.779 | 0.379 | 0.609 |

Response outcome: **V4**

| Background region at visit: | | | **Average bkdg:** | |
|---|---|---|---|---|
| V1 | V2 | V5 | MEAN | SD |
| 0.774 | 0.539 | 0.395 | **0.569** | 0.191 |

*13* | 2 | 3 | forehead

Excess reflection, maybe not BCC
Lesion at hairline; gone by V3
DO NOT USE

| 002BP L3 | V1 | V2 | V3 | V4 | V5 | | | |
|---|---|---|---|---|---|---|---|---|
| Diameter (exam) | 13x8, 35x30 | | 10x12 | 10x12 | Not measurable | | | |
| Height (exam) | Raised | | Raised | Flat | Normal skin | on V1 | l on V2 | on V5 |
| 3D volume | 11.37 | 0.0246 | -12.35 | 27.725 | | | | |
| 3D absol volume | 19.398 | 0.0398 | 16.668 | 33.502 | | | | |

*14* | 2 | 4 | perinasal

Lesion very unclear in photo
Cannot visualize
DO NOT USE

| 002BP L4 | V1 | V2 | V3 | V4 | V5 | | | |
|---|---|---|---|---|---|---|---|---|
| Diameter (exam) | 4x2 | | 4x2 | 4x2 | Not measurable | | | |
| Height (exam) | Raised | | Depresse | Depresse | Normal skin | on V1 | l on V2 | on V5 |
| 3D volume | -0.4275 | -0.2888 | 0.0808 | 0.0156 | -0.016 | | | |
| 3D absol volume | 0.4301 | 0.3099 | 0.2061 | 0.0171 | 0.0605 | | | |

*15* | **2** | **5** | chin

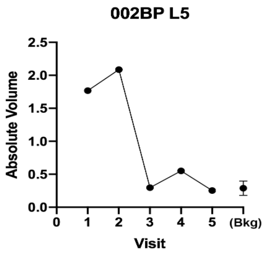

Nodular lesion at the jawline

| 002BP L5 | V1 | V2 | V3 | V4 | V5 |
|---|---|---|---|---|---|
| Height (exam) | Raised | n/d | Sl. raisec | No lesion | No lesion |
| Diameter (exam) | 3 x 3 | n/d | 3 x 3 | visible | lot visible |
| 3D diameter | 3.39 | 3.85 | 2.91 | -- | -- |
| 3D perp diameter | 2.65 | 3.18 | 2.26 | -- | -- |
| 3D av height | 0.242 | 0.260 | 0.032 | -- | -- |
| 3D Av Ht/Area * 10^4 | 84.444 | 66.941 | 15.345 | -- | -- |
| 3D volume | 1.768 | 2.078 | 0.215 | 0.425 | 0.106 |
| 3D absol volume | 1.768 | 2.086 | 0.297 | 0.550 | 0.253 |

Response outcome: **V3**

| Background region at visit: | | | **Average bkdg:** | |
|---|---|---|---|---|
| V1 | V2 | V5 | MEAN | SD |
| 0.413 | 0.243 | 0.208 | **0.288** | 0.110 |

| Lesion Index # | Subject # | Lesion # | Body Location | Diagnosis by Biopsy | Graphical data | Notes & Response to VitD | 3-D lesion data | Visit 1 | Visit 2 | Visit 3 | Visit 4 | Visit 5 | Response outcome | VDR gene |
|---|---|---|---|---|---|---|---|---|---|---|---|---|---|---|

**High-dose Vit D assignment: before V2**

**VDR alleles for patient 3**

| ff | SS |
|---|---|

Lengths are in (mm); Volumes in (mm^3)

| Lesion Index # | Subject # | Lesion # | Body Location |
|---|---|---|---|
| 16 | 3 | 1 | foot |

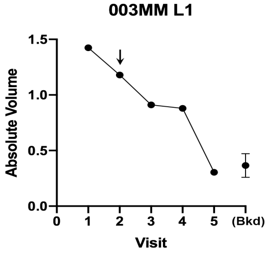

VitD = Yes

| 003MM L1 | V1 | V2 | V3 | V4 | V5 |
|---|---|---|---|---|---|
| Height (exam) | Raised | Raised | Raised | Flat/gone | No lesion |
| Diameter (exam) | 5 x 5 | 5 x 5 | 5 x 5 | 4 x 4 | visible |
| 3D diameter | 5.85 | 5.43 | 5.53 | 5.32 | -- |
| 3D perp diameter | 5.53 | 5.13 | 4.63 | 3.48 | -- |
| 3D av height | 0.004 | 0.003 | 0.005 | 0.006 | -- |
| 3D Av Ht/Area * 10^4 | 0.400 | 0.395 | 0.617 | 0.972 | -- |
| 3D volume | 1.424 | 1.179 | 0.870 | 0.879 | 0.236 |
| 3D absol volume | 1.424 | 1.179 | 0.910 | 0.879 | 0.304 |

Response outcome: **V5**

| Background region at visit: | | | Average bkdg: | |
|---|---|---|---|---|
| V1 | V2 | V5 | MEAN | SD |
| 0.305 | 0.303 | 0.488 | **0.365** | 0.106 |

| Lesion Index # | Subject # | Lesion # | Body Location |
|---|---|---|---|
| 17 | 3 | 2 | Leg |

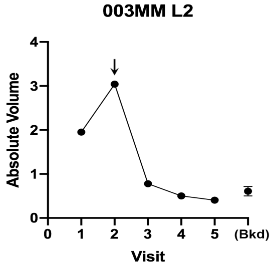

VitD = Yes

| 003MM L2 | V1 | V2 | V3 | V4 | V5 |
|---|---|---|---|---|---|
| Height (exam) | Raised | Raised | Flat/Gon | Flat/Gon | Not visible |
| Diameter (exam) | 5 x 4 | 5 x 4 | 5 x 5 | 5 x 5 | asurable |
| 3D diameter | 5.10 | 5.34 | 1.30 | 1.66 | 1.00 |
| 3D perp diameter | 4.71 | 5.08 | 1.03 | 1.41 | 0.59 |
| 3D av height | 0.143 | 0.134 | 0.014 | 0.028 | 0.019 |
| 3D Av Ht/Area * 10^4 | 18.927 | 15.767 | 33.297 | 37.497 | 96.119 |
| 3D volume | 1.950 | 3.042 | 0.765 | 0.500 | 0.404 |
| 3D absol volume | 1.950 | 3.042 | 0.777 | 0.500 | 0.405 |

Response outcome: **V4**

| Background region at visit: | | | Average bkdg: | |
|---|---|---|---|---|
| V1 | V2 | V5 | MEAN | SD |
| 0.500 | 0.606 | 0.715 | **0.607** | 0.108 |

| Lesion Index # | Subject # | Lesion # | Body Location |
|---|---|---|---|
| 18 | 3 | 3 | ankle |

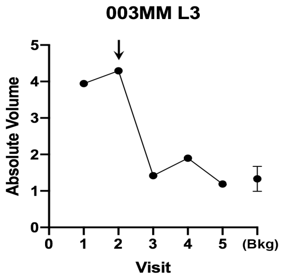

VitD = Yes

| 003MM L3 | V1 | V2 | V3 | V4 | V5 |
|---|---|---|---|---|---|
| Height (exam) | Raised | Raised | Flat | Slightly r | Flat |
| Diameter (exam) | 7 x 7 | 7 x 7 | 6 x 5 | 4 x 4 | 4 x 4 |
| 3D diameter | 5.83 | 8.76 | 4.86 | 5.01 | 4.19 |
| 3D perp diameter | 4.46 | 6.08 | 3.67 | 4.55 | 3.79 |
| 3D av height | 0.040 | 0.063 | 0.033 | 0.043 | 0.017 |
| 3D Av Ht/Area * 10^4 | 4.769 | 3.637 | 5.743 | 6.041 | 3.337 |
| 3D volume | 3.923 | 4.210 | 1.100 | 1.647 | 0.891 |
| 3D absol volume | 3.943 | 4.294 | 1.419 | 1.896 | 1.190 |

Response outcome: **V5**

| Background region at visit: | | | Average bkdg: | |
|---|---|---|---|---|
| V1 | V2 | V5 | MEAN | SD |
| 1.421 | 1.622 | 0.953 | **1.332** | 0.344 |

| Lesion Index # | Subject # | Lesion # | Body Location |
|---|---|---|---|
| 19 | 3 | 4 | leg, popliteal |

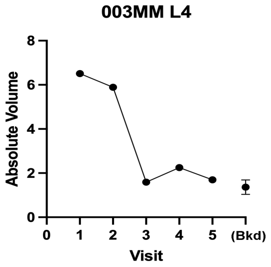

VitD = Yes

| 003MM L4 | V1 | V2 | V3 | V4 | V5 |
|---|---|---|---|---|---|
| Height (exam) | Raised | Raised | Flat | No lesion | No lesion |
| Diameter (exam) | 10 x 5 | 10 x 5 | 7 x 3 | visible | visible |
| 3D diameter | 10.16 | 10.78 | 3.22 | -- | -- |
| 3D perp diameter | 5.83 | 6.57 | 1.84 | -- | -- |
| 3D av height | 0.005 | 0.006 | 0.011 | -- | -- |
| 3D Av Ht/Area * 10^4 | 0.231 | 0.272 | 5.682 | -- | -- |
| 3D volume | 6.474 | 5.754 | 1.229 | 2.172 | -0.079 |
| 3D absol volume | 6.510 | 5.890 | 1.593 | 2.249 | 1.699 |

Response outcome: **V5**

| Background region at visit: | | | Average bkdg: | |
|---|---|---|---|---|
| V1 | V2 | V5 | MEAN | SD |
| 1.073 | 1.437 | 1.589 | **1.366** | 0.265 |

**20** | 3 | 5 | arm

Biopsy at Visit 5: Hyperkeratosis & chronic inflammation (no tumor)

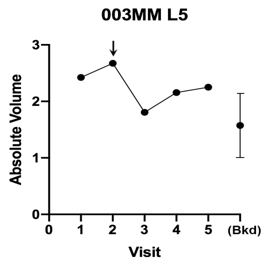

VitD = Yes

Biopsy proved the BCC was gone

| 003MM L5 | V1 | V2 | V3 | V4 | V5 |
|---|---|---|---|---|---|
| Height (exam) | Raised | Raised | Flat | Flat | Flat |
| Diameter (exam) | 10 x 6 | 10 x 6 | 7 x 6 | n/d | 4 x 3 |
| 3D diameter | 9.47 | 10.14 | 4.33 | 4.11 | 4.24 |
| 3D perp diameter | 6.24 | 7.63 | 2.54 | 2.45 | 2.09 |
| 3D av height | 0.053 | 0.061 | 0.024 | 0.008 | 0.031 |
| 3D Av Ht/Area * 10^4 | 2.715 | 2.441 | 6.508 | 2.290 | 9.877 |
| 3D volume | 2.271 | 2.462 | 1.277 | 1.971 | 1.762 |
| 3D absol volume | 2.423 | 2.674 | 1.806 | 2.155 | 2.252 |

**V5**

Background region at visit: **Average bkdg:**

| V1 | V2 | V5 | MEAN | SD |
|---|---|---|---|---|
| 1.337 | 1.160 | 2.221 | **1.573** | 0.568 |

**21** | 3 | 6 | back

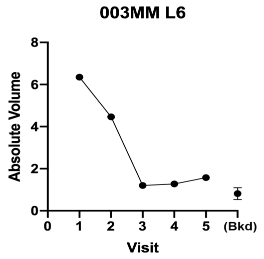

Clinically gone by V3; but a flat scar remains

| 003MM L6 | V1 | V2 | V3 | V4 | V5 |
|---|---|---|---|---|---|
| Height (exam) | Raised | Raised | Slightly r | Flat | Flat |
| Diameter (exam) | 9 x 4 | 9 x 4 | 7 x 4 | 6 x 5 | 7 x 4 |
| 3D diameter | 7.67 | 8.16 | 7.45 | 7.32 | 2.73 |
| 3D perp diameter | 6.03 | 5.47 | 3.90 | 4.97 | 1.02 |
| 3D av height | 0.200 | 0.147 | 0.041 | 0.073 | 0.015 |
| 3D Av Ht/Area * 10^4 | 13.572 | 10.047 | 4.024 | 6.139 | 13.966 |
| 3D volume | 6.335 | 4.377 | 1.084 | 0.147 | -0.200 |
| 3D absol volume | 6.352 | 4.466 | 1.204 | 1.276 | 1.578 |

**V3**

Background region at visit: **Average bkdg:**

| V1 | V2 | V5 | MEAN | SD |
|---|---|---|---|---|
| 0.520 | 1.077 | 0.845 | **0.814** | 0.280 |

| *Lesion Index #* | Subject # | Lesion # | Body Location | Diagnosis by Biopsy | Graphical data | Notes & Response to VitD | 3-D lesion data | Visit 1 | Visit 2 | Visit 3 | Visit 4 | Visit 5 | Response outcome | VDR gene |
|---|---|---|---|---|---|---|---|---|---|---|---|---|---|---|

In retropsect for patient 4, lesions # 1-5 were atrophic shave bipsy scars; lesion #6 was an SCC.

**High-dose Vit D assignment: before V3**

Lengths are in (mm); Volumes in (mm^3)

**VDR alleles for patient 4**

| ff | LS |
|---|---|

**22** | 4 | 1 | leg (knee)

(Did not graph)

Atrophic BCC (negative volumes)

| 004JB L1 | V1 | V2 | V3 | V4 | V5 |
|---|---|---|---|---|---|
| Diameter (exam) | 18x11 | 18x11 | 15x10 | 12x9 | 9x12 |
| Height (exam) | scaly shir | scaly shin | Flat | Flat | Flat |
| 3D diameter | 17.119 | 17.075 | 16.051 | 13.021 | 10.797 |
| 3D perp diameter | 12.963 | 12.134 | 12.118 | 9.875 | 9.283 |
| 3D av height | 0.004 | 0.013 | 0.003 | 0.004 | 0.001 |
| 3D volume | 0.635 | 1.95 | 0.423 | 0.396 | 0.05 |
| 3D absol volume | 18.735 | 17.361 | 17.119 | 8.996 | 8.244 |

Background region at visit: **Average bkdg:**

| V1 | V2 | V5 | MEAN | SD |
|---|---|---|---|---|
| 4.027 | 5.764 | 6.245 | **5.345** | 1.167 |

**23** | 4 | 2 | back

(Did not graph)

Atrophic BCC (negative volumes)

| 004JB L2 | V1 | V2 | V3 | V4 | V5 |
|---|---|---|---|---|---|
| Diameter (exam) | 5x5 | 5x5 | 9x6 | 3x6 | 3x6 |
| Height (exam) | | | Raised | Flat | Looks normal |
| 3D diameter | 6.486 | 7.876 | 5.92 | 6.539 | 6.518 |
| 3D perp diameter | 5.546 | 6.861 | 5.081 | 5.624 | 5.612 |
| 3D av height | 0.029 | 0.008 | 0.009 | 0.016 | 0.009 |
| 3D volume | 0.656 | 0.282 | 0.165 | 0.368 | 0.217 |
| 3D absol volume | 0.776 | 0.876 | 0.878 | 0.438 | 0.406 |

Background region at visit: **Average bkdg:**

| V1 | V2 | V5 | MEAN | SD |
|---|---|---|---|---|
| 0.275 | 0.317 | 0.516 | **0.369** | 0.129 |

**24** | 4 | 3 | back

(Did not graph)

Atrophic BCC (negative volumes)
Lesion became more fibrotic-looking after each PDT Rx.

| 004JB L3 | V1 | V2 | V3 | V4 | V5 |
|---|---|---|---|---|---|
| Diameter (exam) | 10x6 | 8x9 | 9x8 | 8x10 | 8x10 |
| Height (exam) | | | | Depresse | Depressed |
| 3D diameter | 12.592 | 10.352 | 9.247 | 9.904 | 9.75 |
| 3D perp diameter | 8.302 | 7.865 | 7.145 | 7.062 | 7.16 |
| 3D av height | 0 | 0 | 0 | 0 | 0.001 |
| 3D volume | 0 | 0.014 | 0.007 | 0.008 | 0.031 |
| 3D absol volume | 20.665 | 15.361 | 18.422 | 13.218 | 12.183 |

Background region at visit: **Average bkdg:**

| V1 | V2 | V5 | MEAN | SD |
|---|---|---|---|---|
| 1.348 | 1.98 | 1.559 | **1.629** | 0.322 |

**25** | 4 | 4 | back

(Did not graph)

Old sunken biopsy scar; (Shiny surface in a smooth depression in the skin. The 3D quantif'n shows negative vol)

| 004JB L4 | V1 | V2 | V3 | V4 | V5 |
|---|---|---|---|---|---|
| Diameter (exam) | 10x9 | 10x9 | 10x9 | 10x9 | 10x9 |
| Height (exam) | | | | | Depressed |
| 3D diameter | 12.395 | 11.2 | 10.691 | 11.608 | 9.724 |
| 3D perp diameter | 9.87 | 8.591 | 8.183 | 8.436 | 7.603 |
| 3D av height | 0 | 0 | 0 | 0 | 0 |
| 3D Av Ht/area 10^4 | nd | nd | nd | nd | nd |
| 3D volume | 0.014 | 0 | 0 | 0.002 | 0 |
| 3D absol volume | 21.502 | 18.635 | 20.919 | 19.452 | 15.163 |

| Background region at visit: | | | **Average bkdg:** | |
|---|---|---|---|---|
| V1 | V2 | V5 | MEAN | SD |
| 2.575 | 1.222 | 2.042 | **1.946** | 0.682 |

**26** | 4 | 5 | back

(Did not graph)

Sunken nature of depressed lesion only became apparent upon magnification with the 3D images, and neg volumes

| 004JB L5 | V1 | V2 | V3 | V4 | V5 |
|---|---|---|---|---|---|
| Diameter (exam) | 6x6 | 5x7 | 5x10 | 5x10 | 5x10 |
| Height (exam) | | | | | Depressed |
| 3D diameter | 9.772 | 10.064 | 10.151 | 11.058 | 9.667 |
| 3D perp diameter | 8.895 | 6.977 | 6.335 | 7.191 | 5.798 |
| 3D av height | 0.001 | 0.001 | 0 | 0 | 0 |
| 3D Av Ht/area 10^4 | nd | nd | nd | nd | nd |
| 3D volume | 0.027 | 0.03 | 0.001 | 0.025 | 0 |
| 3D absol volume | 8.231 | 11.035 | 15.937 | 11.669 | 7.604 |

| Background region at visit: | | | **Average bkdg:** | |
|---|---|---|---|---|
| V1 | V2 | V5 | MEAN | SD |
| 0.893 | 1.014 | 0.713 | **0.873** | 0.151 |

**27** | 4 | 6 | back

Biopsy at Visit 5: Squamous cell CA

SCC proven by biopsy
DO NOT USE

| 004JB L6 | V1 | V2 | V3 | V4 | V5 |
|---|---|---|---|---|---|
| Height (exam) | | | Not visit | Flat | Flat |
| Diameter (exam) | 4x4 | 4x3 | 4x3 | 2x2 | 2x2 |
| 3D diameter | | | | | |
| 3D perp diameter | | | | | |
| 3D av height | | | | | |
| 3D Av Ht/Area * 10^4 | | | | | |
| 3D volume | 0.9995 | 0.7911 | 0.2171 | 0.2441 | 0.2893 |
| 3D absol volume | 0.9995 | 0.7911 | 0.23 | 0.259 | 0.2896 |

*28* | 4 | 7 | back

Very small, looks like folliculitis AT gone at V3. DO NOT USE

| 004JB L7 | V1 | V2 | V3 | V4 | V5 |
|---|---|---|---|---|---|
| Diameter (exam) | 4x5 | 4x4 | 4x2 | 0x0 | 0x0 |
| Height (exam) | | | Flat | Not visibl | Not visible |
| 3D volume | 0.5448 | -0.077 | -0.006 | -0.038 | 0.2778 |
| 3D absol volume | 0.6109 | 0.2205 | 0.3864 | 0.4493 | 0.3634 |

*29* | 4 | 8 | R shoulder

Very small, questionable whether a
DO NOT USE

| 004JB L8 | V1 | V2 | V3 | V4 | V5 |
|---|---|---|---|---|---|
| Diameter (exam) | 3x3 | 3x3 | 3x3 | 3x4 | 3x4 |
| Height (exam) | | | Raised | Raised | Not visible |
| 3D volume | 0.7346 | 0.506 | 0.7747 | 0.4275 | |
| 3D absol volume | 0.7369 | 0.5464 | 0.9384 | 0.6051 | |

*30* | 4 | 9 | R neck

No lesion visible. DO NOT USE.

| 004JB L9 | V1 | V2 | V3 | V4 | V5 |
|---|---|---|---|---|---|
| Diameter (exam) | | 4x3 | 3x4 | 3x4 | 3x4 |
| Height (exam) | | | Not visit | Not visibl | Not visible |
| 3D volume | | 0.3138 | | | |
| 3D absol volume | | 0.4064 | | | |

| Lesion Index # | Subject # | Lesion # | Body Location | Diagnosis by Biopsy | Graphical data | Notes & Response to VitD | 3-D lesion data | Visit 1 | Visit 2 | Visit 3 | Visit 4 | Visit 5 | Response outcome | VDR gene |
|---|---|---|---|---|---|---|---|---|---|---|---|---|---|---|

**High-dose Vit D assignment: before V2**

Lengths are in (mm); Volumes in (mm^3)

**VDR alleles for patient 5**

| Ff | LS |
|---|---|

*31* | **5** | **1** | breast

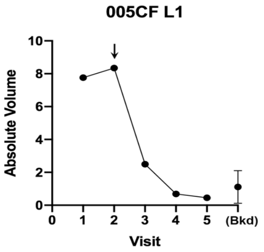

VitD = yes

| 005CF L1 | V1 | V2 | V3 | V4 | V5 |
|---|---|---|---|---|---|
| Height (exam) | | | Flat | Flat | No lesion |
| Diameter (exam) | 13 x 12 | 13 x 12 | 8 x 8 | 5 x 4 | visible |
| 3D diameter | 13.43 | 13.94 | 5.47 | 5.95 | -- |
| 3D perp diameter | 11.99 | 10.85 | 4.83 | 4.65 | -- |
| 3D av height | 0.067 | 0.072 | 0.013 | 0.006 | -- |
| 3D Av Ht/Area * 10^4 | 1.316 | 1.486 | 1.613 | 0.666 | -- |
| 3D volume | 6.347 | 7.977 | 0.957 | -0.219 | 0.034 |
| 3D absol volume | 7.764 | 8.348 | 2.496 | 0.685 | 0.456 |

**V4**

| Background region at visit: | | | Average bkdg: | |
|---|---|---|---|---|
| V1 | V2 | V5 | MEAN | SD |
| 1.462 | 1.182 | 0.691 | **1.112** | **0.995** |

*32* | **5** | **2** | arm

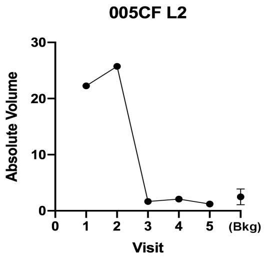

Thin superficial lesion, gone at V3

| 005CF L2 | V1 | V2 | V3 | V4 | V5 |
|---|---|---|---|---|---|
| Height (exam) | | | Flat | Flat | Flat |
| Diameter (exam) | 9 x 7 | 9 x 7 | 5 x 4 | 5 x 5 | 5 x 5 |
| 3D diameter | 8.94 | 8.00 | 7.58 | 7.36 | 7.36 |
| 3D perp diameter | 8.33 | 6.76 | 6.49 | 5.56 | 5.56 |
| 3D av height | 0.012 | 0.030 | 0.052 | 0.036 | 0.004 |
| 3D Av Ht/Area * 10^4 | 0.505 | 1.765 | 3.347 | 2.748 | 0.337 |
| 3D volume | 21.768 | 25.700 | 1.592 | 2.083 | 1.002 |
| 3D absol volume | 22.260 | 25.737 | 1.663 | 2.095 | 1.214 |

**V3**

| Background region at visit: | | | Average bkdg: | |
|---|---|---|---|---|
| V1 | V2 | V5 | MEAN | SD |
| 4.118 | 1.596 | 1.761 | **2.492** | 1.411 |

*33* | **5** | **3** | arm

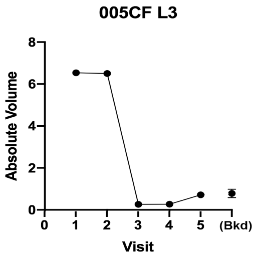

Thin superficial lesion, gone at V3

| 005CF L3 | V1 | V2 | V3 | V4 | V5 |
|---|---|---|---|---|---|
| Height (exam) | | | Flat | No lesion | No lesion |
| Diameter (exam) | 9 x 6 | 9 x 6 | 4 x 5 | visible | visible |
| 3D diameter | 8.31 | 7.71 | 8.27 | -- | -- |
| 3D perp diameter | 7.64 | 6.87 | 5.42 | -- | -- |
| 3D av height | 0.022 | 0.016 | 0.004 | -- | -- |
| 3D Av Ht/Area * 10^4 | 1.125 | 0.978 | 0.241 | -- | -- |
| 3D volume | 6.363 | 6.322 | -0.096 | -0.092 | 0.469 |
| 3D absol volume | 6.535 | 6.505 | 0.264 | 0.272 | 0.716 |

**V3**

| Background region at visit: | | | Average bkdg: | |
|---|---|---|---|---|
| V1 | V2 | V5 | MEAN | SD |
| 0.746 | 0.593 | 1.001 | **0.780** | 0.206 |

*34* | **5** | **4** | back | Biopsy at Visit 5: BCC, nodular

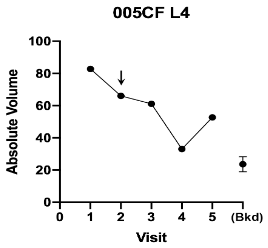

VitD=No

Huge lesion; shrank, then grew

| 005CF L4 | V1 | V2 | V3 | V4 | V5 |
|---|---|---|---|---|---|
| Height (exam) | | | | Mostly fla | Mostly flat |
| Diameter (exam) | 39x15 | 39x15 | 39x22 | 35x20 | 35x20 |
| 3D diameter | 37.58 | 36.48 | 39.08 | 41.24 | 40.29 |
| 3D perp diameter | 25.08 | 24.46 | 21.36 | 23.22 | 19.07 |
| 3D av height | 0.060 | 0.071 | 0.078 | 0.090 | 0.144 |
| 3D Av Ht/Area * 10^4 | 0.193 | 0.243 | 0.273 | 0.275 | 0.521 |
| 3D volume | 80.191 | 57.438 | 34.189 | 23.461 | 40.919 |
| 3D absol volume | 82.831 | 66.033 | 61.138 | 33.065 | 52.698 |

**NC**

| Background region at visit: | | | Average bkdg: | |
|---|---|---|---|---|
| V1 | V2 | V5 | MEAN | SD |
| 25.747 | 18.293 | 26.955 | **23.665** | 4.692 |

| 35 | 5 | 5 | back | Biopsy at Visit 5: BCC, infiltrative |
|---|---|---|---|---|

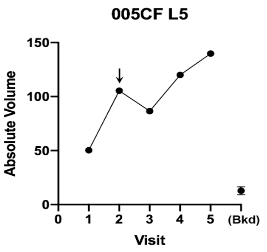

Giant, ulcerated, lesion on back

VitD=Yes

| 005CF L5 | V1 | V2 | V3 | V4 | V5 |
|---|---|---|---|---|---|
| Height (exam) | with | with 6x6 | with | with | with |
| Diameter (exam) | 15 x 12 | 15 x 12 | 17 x 10 | 15 x 12 | 15 x 12 |
| 3D diameter | 13.31 | 17.62 | 20.32 | 20.62 | 20.14 |
| 3D perp diameter | 12.10 | 15.91 | 12.47 | 14.27 | 14.86 |
| 3D av height | 0.079 | 0.272 | 0.308 | 0.483 | 0.318 |
| 3D Av Ht/Area * 10^4 | 1.550 | 3.077 | 3.647 | 5.049 | 3.302 |
| 3D volume | -5.863 | 4.369 | 29.653 | 99.505 | 89.709 |
| 3D absol volume | 50.306 | 105.462 | 86.617 | 120.030 | ######## |

NC

Background region at visit: / Average bkdg:

| V1 | V2 | V5 | MEAN | SD |
|---|---|---|---|---|
| 16.809 | 9.940 | 11.666 | **12.805** | 3.573 |

| 36 | 5 | 6 | scapula | Biopsy at Visit 5: BCC, nodular and infiltrative |
|---|---|---|---|---|

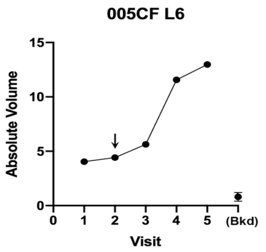

Nodule, grew slower with VitD

(VitD = yes)

| 005CF L6 | V1 | V2 | V3 | V4 | V5 |
|---|---|---|---|---|---|
| Height (exam) | Raised | Raised | Raised | Raised | Raised |
| Diameter (exam) | 9 x 5 | 9 x 5 | 8 x 5 | 8 x 5 | 10 x 5 |
| 3D diameter | 8.01 | 8.42 | 8.08 | 8.90 | 9.09 |
| 3D perp diameter | 4.98 | 5.09 | 4.78 | 5.22 | 6.58 |
| 3D av height | 0.124 | 0.142 | 0.205 | 0.348 | 0.315 |
| 3D Av Ht/Area * 10^4 | 9.363 | 9.912 | 15.761 | 22.208 | 16.310 |
| 3D volume | 3.485 | 4.233 | 5.491 | 11.366 | 12.573 |
| 3D absol volume | 4.038 | 4.421 | 5.631 | 11.567 | 12.980 |

NC

Background region at visit: / Average bkdg:

| V1 | V2 | V5 | MEAN | SD |
|---|---|---|---|---|
| 0.989 | 0.709 | 0.713 | **0.803** | 0.161 |

| 37 | 5 | 7 | scapula | Biopsy at Visit 5: BCC, nodular & and adenoid |
|---|---|---|---|---|

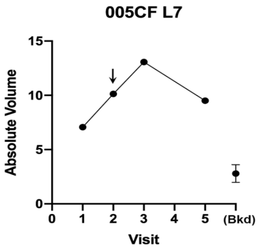

VitD= no

Atrophic BCC; never responded

| 005CF L7 | V1 | V2 | V3 | V4 | V5 |
|---|---|---|---|---|---|
| Height (exam) | Raised | Raised | Raised | Raised | Raised |
| Diameter (exam) | 12 x 10 | 12 x 10 | 10 x 9 | 12 x 6 | 12 x 10 |
| 3D diameter | 11.75 | 12.81 | 13.59 | n/d | 12.40 |
| 3D perp diameter | 8.21 | 7.03 | 7.11 | n/d | 10.84 |
| 3D av height | 0.060 | 0.166 | 0.238 | n/d | 0.067 |
| 3D Av Ht/Area * 10^4 | 1.918 | 5.381 | 7.068 | n/d | 1.591 |
| 3D volume | 1.021 | 7.662 | 12.702 | n/d | 0.815 |
| 3D absol volume | 7.069 | 10.128 | 13.066 | n/d | 9.510 |

NC

Background region at visit: / Average bkdg:

| V1 | V2 | V5 | MEAN | SD |
|---|---|---|---|---|
| 2.409 | 2.239 | 3.733 | **2.794** | 0.818 |

| 38 | 5 | 8 | neck | Biopsy at Visit 5: BCC, micronodular |
|---|---|---|---|---|

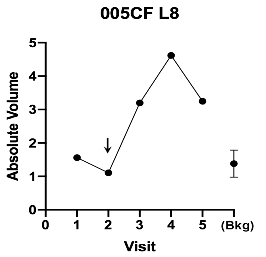

Small pigmented BCC, unresponsive to PDT

VitD= no

| 005CF L8 | V1 | V2 | V3 | V4 | V5 |
|---|---|---|---|---|---|
| Height (exam) | Raised | Raised | Raised | Raised | Raised |
| Diameter (exam) | 5 x 5 | 5 x 5 | 7 x 6 | 7 x 5 | 6 x 5 |
| 3D diameter | 7.24 | 6.56 | 7.88 | 8.23 | 8.21 |
| 3D perp diameter | 5.83 | 4.72 | 5.29 | 5.88 | 5.27 |
| 3D av height | 0.045 | 0.026 | 0.080 | 0.114 | 0.085 |
| 3D Av Ht/Area * 10^4 | 3.347 | 2.588 | 5.857 | 7.325 | 5.969 |
| 3D volume | 0.941 | -0.065 | 1.341 | 3.032 | 2.134 |
| 3D absol volume | 1.561 | 1.106 | 3.199 | 4.618 | 3.246 |

NC

Background region at visit: / Average bkdg:

| V1 | V2 | V5 | MEAN | SD |
|---|---|---|---|---|
| 1.824 | 1.028 | 1.296 | **1.382** | 0.405 |

| 39 | 5 | 9 | arm | Biopsy at Visit 5: micronodular and infiltrative |
|---|---|---|---|---|

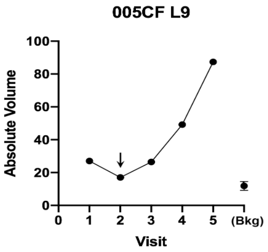

Big annular lesion

VitD= no

| 005CF L9 | V1 | V2 | V3 | V4 | V5 |
|---|---|---|---|---|---|
| Height (exam) | Raised | Raised | Raised | Raised | Raised |
| Diameter (exam) | 18 x 8 | 18 x 8 | 20 x 10 | 20 x 15 | 20 x 15 |
| 3D diameter | 20.68 | 20.10 | 21.39 | 23.10 | 23.47 |
| 3D perp diameter | 10.96 | 10.48 | 13.28 | 14.88 | 15.76 |
| 3D av height | 0.164 | 0.101 | 0.142 | 0.220 | 0.343 |
| 3D Av Ht/Area * 10^4 | 2.083 | 1.373 | 1.509 | 1.944 | 2.840 |
| 3D volume | 27.052 | 15.551 | 26.381 | 49.177 | 87.351 |
| 3D absol volume | 27.065 | 17.004 | 26.444 | 49.244 | 87.405 |

NC

Background region at visit: / Average bkdg:

| V1 | V2 | V5 | MEAN | SD |
|---|---|---|---|---|
| 9.873 | 12.824 | 12.967 | **11.888** | 1.747 |

| Lesion Index # | Subject # | Lesion # | Body Location | Diagnosis by Biopsy |
|---|---|---|---|---|
| *40* | 5 | 10 | arm | Biopsy at Visit 5: Dermal fibrosis, solar elastosis |

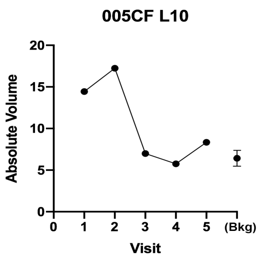

Superficial lesion, Resolved at V3

| 005CF L10 | V1 | V2 | V3 | V4 | V5 |
|---|---|---|---|---|---|
| Height (exam) | Sl. Raised | Sl. Raised | Sl. Raise( | Flat | Slightly raised |
| Diameter (exam) | 10 x 6 | 10 x 6 | 6 x 6 | 5 x 5 | 5 x 5 |
| 3D diameter | 9.91 | 9.08 | -- | -- | -- |
| 3D perp diameter | 7.22 | 7.11 | -- | -- | -- |
| 3D av height | 0.076 | 0.026 | -- | -- | -- |
| 3D Av Ht/Area * 10^4 | 3.296 | 1.266 | -- | -- | -- |
| 3D volume | 14.374 | 17.243 | 6.942 | 5.660 | 8.322 |
| 3D absol volume | 14.443 | 17.244 | 6.993 | 5.751 | 8.341 |

**V3**

| Background region at visit: | | | Average bkdg: | |
|---|---|---|---|---|
| V1 | V2 | V5 | MEAN | SD |
| 6.133 | 7.497 | 5.666 | **6.432** | 0.951 |

| *Lesion Index #* | Subject # | Lesion # | Body Location | Diagnosis by Biopsy | Graphical data | Notes & Response to VitD | 3-D lesion data | Visit 1 | Visit 2 | Visit 3 | Visit 4 | Visit 5 | Response outcome | VDR gene |
|---|---|---|---|---|---|---|---|---|---|---|---|---|---|---|

High-dose Vit D assignment: before V3

Lengths are in (mm); Volumes in (mm^3)

VDR alleles for patient 6

| Ff | LS |
|---|---|

| Lesion Index # | Subject # | Lesion # | Body Location | Diagnosis by Biopsy |
|---|---|---|---|---|
| *41* | 6 | 1 | back | Biopsy at Visit 5: BCC, nodular, microdular, & adenoid |

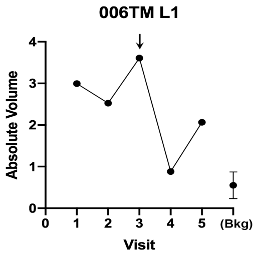

(VitD=Yes)

Ulcerative; a nodular component shrank after VD, but never all gone

| 006TM L1 | V1 | V2 | V3 | V4 | V5 |
|---|---|---|---|---|---|
| Height (exam) | Raised | Raised w/ | Raised | Flat, cent | Flat, central papule |
| Diameter (exam) | 12x17 | 12x17 | 12x17 | 2x5 | 3x5 |
| 3D diameter | 4.63 | 5.26 | 7.02 | 5.68 | 6.82 |
| 3D perp diameter | 2.34 | 4.48 | 3.19 | 2.67 | 3.37 |
| 3D av height | 0.067 | 0.102 | 0.220 | 0.090 | 0.140 |
| 3D Av Ht/Area * 10^4 | 17.625 | 13.659 | 26.832 | 16.482 | 17.132 |
| 3D volume | 2.395 | -0.084 | 3.607 | 0.847 | 2.031 |
| 3D absol volume | 2.996 | 2.526 | 3.609 | 0.882 | 2.066 |

**NC**

| Background region at visit: | | | Average bkdg: | |
|---|---|---|---|---|
| V1 | V2 | V5 | MEAN | SD |
| 0.486 | 0.901 | 0.271 | **0.553** | 0.320 |

| Lesion Index # | Subject # | Lesion # | Body Location |
|---|---|---|---|
| *42* | 6 | 2 | back |

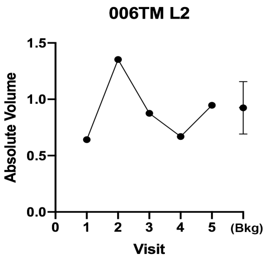

Flat, sunken, looks like scar except papule raised at left pole which was gone by V3
We quantified just the papule

| 006TM L2 | V1 | V2 | V3 | V4 | V5 |
|---|---|---|---|---|---|
| Height (exam) | Raised | Raised | Flat | Flat | Flat |
| Diameter (exam) | 18x15 | 18x15 | 18x15 | 10x10 | 10x10 |
| 3D diameter | 18.24 | 17.63 | 16.42 | -- | -- |
| 3D perp diameter | 12.97 | 12.18 | 12.54 | -- | -- |
| 3D av height | 0.010 | 0.001 | 0.004 | -- | -- |
| 3D Av Ht/Area * 10^4 | 0.131 | 0.020 | 0.058 | -- | -- |
| 3D volume | 0.185 | -1.349 | 0.722 | -0.165 | -0.014 |
| 3D absol volume | 0.642 | 1.353 | 0.875 | 0.669 | 0.947 |

**V3**

| Background region at visit: | | | Average bkdg: | |
|---|---|---|---|---|
| V1 | V2 | V5 | MEAN | SD |
| 1.103 | 0.662 | 1.008 | **0.925** | 0.232 |

| Lesion Index # | Subject # | Lesion # | Body Location |
|---|---|---|---|
| *43* | 6 | 3 | back |

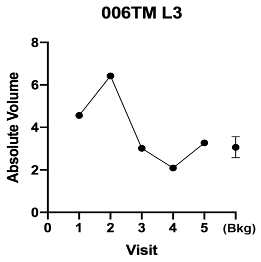

Small lesion, gone by V3

| 006TM L3 | V1 | V2 | V3 | V4 | V5 |
|---|---|---|---|---|---|
| Height (exam) | Raised | Raised | Flat | Flat | Flat |
| Diameter (exam) | 10x10 | 10x10 | 10x10 | 6x9 | 6x9 |
| 3D diameter | 11.32 | 11.76 | 2.39 | 2.80 | 1.36 |
| 3D perp diameter | 7.46 | 8.66 | 2.09 | 2.14 | 1.05 |
| 3D av height | 0.067 | 0.088 | 0.020 | 0.051 | 0.042 |
| 3D Av Ht/Area * 10^4 | 2.410 | 2.681 | 12.432 | 26.447 | 92.123 |
| 3D volume | 4.144 | 5.283 | 1.592 | 0.959 | -1.616 |
| 3D absol volume | 4.562 | 6.422 | 3.018 | 2.092 | 3.267 |

**V3**

| Background region at visit: | | | Average bkdg: | |
|---|---|---|---|---|
| V1 | V2 | V5 | MEAN | SD |
| 2.935 | 2.645 | 3.607 | **3.062** | 0.494 |

| Lesion Index # | Subject # | Lesion # | Body Location |
|---|---|---|---|
| 44 | 6 | 4 | back |

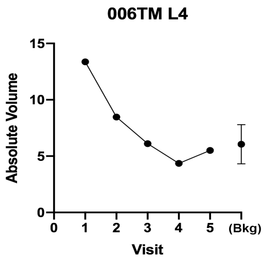

Flat lesion, gone by V3

| 006TM L4 | V1 | V2 | V3 | V4 | V5 |
|---|---|---|---|---|---|
| Height (exam) | Raised | Raised | Flat | Flat | Flat |
| Diameter (exam) | 10x22 | 10x22 | 10x22 | 9x15 | 9x15 |
| 3D diameter | 20.77 | 20.34 | 18.09 | -- | -- |
| 3D perp diameter | 11.60 | 11.61 | 10.91 | -- | -- |
| 3D av height | 0.051 | 0.048 | 0.011 | -- | -- |
| 3D Av Ht/Area * 10^4 | 0.616 | 0.604 | 0.168 | -- | -- |
| 3D volume | 4.183 | 6.463 | -3.085 | -2.390 | -3.263 |
| 3D absol volume | 13.372 | 8.463 | 6.109 | 4.361 | 5.509 |

Response outcome: **V3**

| Background region at visit: | | | Average bkdg: | |
|---|---|---|---|---|
| V1 | V2 | V5 | MEAN | SD |
| 4.146 | 6.458 | 7.555 | **6.053** | 1.740 |

| Lesion Index # | Subject # | Lesion # | Body Location |
|---|---|---|---|
| 45 | 6 | 5 | back |

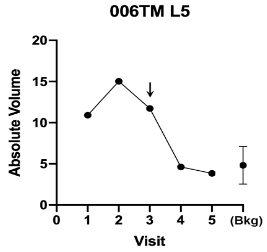

VitD=Yes

Sunken, arising from a scar line; red erosions at one pole at V1, none at later visits. Flat by V4

| 006TM L5 | V1 | V2 | V3 | V4 | V5 |
|---|---|---|---|---|---|
| Height (exam) | Raised | Raised | Flat | Flat | Flat |
| Diameter (exam) | 10x15 | 10x15 | 10x10 | 10x12 | 10x12 |
| 3D diameter | 17.89 | 17.45 | 15.65 | -- | -- |
| 3D perp diameter | 13.12 | 11.94 | 10.16 | -- | -- |
| 3D av height | 0.009 | 0.003 | 0.002 | -- | -- |
| 3D Av Ht/Area * 10^4 | 0.121 | 0.051 | 0.048 | -- | -- |
| 3D volume | -8.281 | -13.947 | -11.154 | -2.229 | 0.200 |
| 3D absol volume | 10.902 | 15.025 | 11.733 | 4.622 | 3.837 |

Response outcome: **V4**

| Background region at visit: | | | Average bkdg: | |
|---|---|---|---|---|
| V1 | V2 | V5 | MEAN | SD |
| 6.262 | 6.020 | 2.200 | **4.827** | 2.279 |

| *Lesion Index #* | Subject # | Lesion # | Body Location | Diagnosis by Biopsy | Graphical data | Notes & Response to VitD | 3-D lesion data | Visit 1 | Visit 2 | Visit 3 | Visit 4 | Visit 5 | Response outcome | VDR gene |
|---|---|---|---|---|---|---|---|---|---|---|---|---|---|---|

**High-dose Vit D assignment: before V3**

Lengths are in (mm); Volumes in (mm^3)

**VDR alleles for patient 7**

| Ff | SS |
|---|---|

| Lesion Index # | Subject # | Lesion # | Body Location | Diagnosis by Biopsy |
|---|---|---|---|---|
| 46 | 7 | 1 | arm | Biopsy at Visit 5: Bx: dermal scar chronic folliclitis |

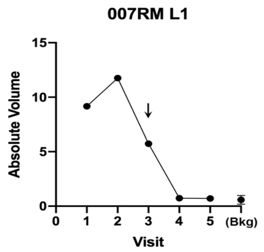

VitD= No

Large flat lesion, gone by V4

| 007RM L1 | V1 | V2 | V3 | V4 | V5 |
|---|---|---|---|---|---|
| Height (exam) | Raised | Raised | Sl. Raise | Sl. Raised | Flat |
| Diameter (exam) | 8x10 | 8x10 | 5x10 | 8x4 | 2x2 |
| 3D diameter | 12.95 | 11.42 | 7.74 | 7.91 | 5.88 |
| 3D perp diameter | 10.07 | 7.41 | 3.51 | 4.56 | 4.17 |
| 3D av height | 0.063 | 0.066 | 0.027 | 0.010 | 0.011 |
| 3D Av Ht/Area * 10^4 | 1.508 | 2.368 | 2.681 | 0.821 | 1.375 |
| 3D volume | 9.110 | 11.703 | 5.639 | 0.233 | -0.247 |
| 3D absol volume | 9.169 | 11.761 | 5.734 | 0.728 | 0.700 |

Response outcome: **V4**

| Background region at visit: | | | Average bkdg: | |
|---|---|---|---|---|
| V1 | V2 | V5 | MEAN | SD |
| 0.439 | 0.590 | 0.734 | **0.587** | 0.147 |

| Lesion Index # | Subject # | Lesion # | Body Location |
|---|---|---|---|
| 47 | 7 | 2 | chest |

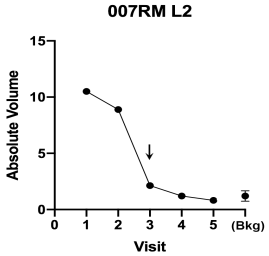

VitD= No

VitD (gone at V4)

| 007RM L2 | V1 | V2 | V3 | V4 | V5 |
|---|---|---|---|---|---|
| Height (exam) | Two dom | Two dom | Two pap | Flat, ill-d | Flat, ill-defined |
| Diameter (exam) | 5x11 | 5x11 | 5x4 | N/A | Not visible |
| 3D diameter | 4.34 | 4.90 | -- | -- | -- |
| 3D perp diameter | 3.60 | 3.93 | -- | -- | -- |
| 3D av height | 0.278 | 0.261 | -- | -- | -- |
| 3D Av Ht/Area * 10^4 | 56.210 | 42.643 | -- | -- | -- |
| 3D volume | 10.241 | 8.117 | 1.552 | -0.862 | 0.157 |
| 3D absol volume | 10.512 | 8.897 | 2.127 | 1.202 | 0.814 |

Response outcome: **V4**

| Background region at visit: | | | Average bkdg: | |
|---|---|---|---|---|
| V1 | V2 | V5 | MEAN | SD |
| 1.743 | 0.912 | 0.964 | **1.207** | 0.465 |

| 48 | 7 | 3 | chest |
|---|---|---|---|

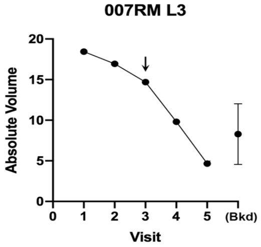

VitD=Yes

Lesion shrank every time but still visible at V5; looks like scar

| 007RM L3 | V1 | V2 | V3 | V4 | V5 |
|---|---|---|---|---|---|
| Height (exam) | Raised | Raised | Depresse | Flat | Flat |
| Diameter (exam) | 10x13 | 10x13 | 12x13 | 15x10 | 15x10 |
| 3D diameter | 14.19 | 13.70 | 12.58 | 11.83 | 13.65 |
| 3D perp diameter | 13.42 | 12.96 | 9.99 | 9.40 | 11.11 |
| 3D av height | 0.126 | 0.079 | 0.082 | 0.076 | 0.042 |
| 3D Av Ht/Area * 10^4 | 2.102 | 1.415 | 2.045 | 2.158 | 0.865 |
| 3D volume | 13.017 | 10.637 | 4.673 | 0.422 | 3.171 |
| 3D absol volume | 18.445 | 16.947 | 14.689 | 9.810 | 4.654 |

**V4**

Background region at visit: **Average bkdg:**

| V1 | V2 | V5 | MEAN | SD |
|---|---|---|---|---|
| 9.230 | 11.466 | 4.186 | **8.294** | 3.729 |

| 49 | 7 | 4 | leg | V1 |
|---|---|---|---|---|
| | | | | Biopsy: SCC |

**SCC, do not include**

| 007RM L4 | V1 | V2 | V3 | V4 | V5 |
|---|---|---|---|---|---|
| Diameter (exam) | 5x12 | 5x12 | 5x12 | Not meas | Not measurable |
| Height (exam) | Raised | Raised | Flat | Nothing v | Nothing visible |
| 3D volume | 6.5017 | 4.0638 | -0.054 | | |
| 3D absol volume | 7.4166 | 4.8476 | 0.2063 | | |

| *Lesion Index #* | Subject # | Lesion # | Body Location | Diagnosis by Biopsy | Graphical data | Notes & Response to VitD | 3-D lesion data | Visit 1 | Visit 2 | Visit 3 | Visit 4 | Visit 5 | Response outcome | VDR gene |
|---|---|---|---|---|---|---|---|---|---|---|---|---|---|---|

**High-dose Vit D assignment: before V3**

Lengths are in (mm); Volumes in (mm^3)

**VDR alleles for patient 8**

| Ff | LS |
|---|---|

| 50 | 8 | 1 | auricular |
|---|---|---|---|

Already gone at V2; DO NOT USE

| 008DM L1 | V1 | V2 | V3 | V4 | V5 |
|---|---|---|---|---|---|
| Height (exam) | Slight rai: | flat | flat, alm | flat, almc | flat, almost invisible |
| Diameter (exam) | 5x5 | 5x5 | 3x3 | 2x2 | 2x2 |
| 3D volume | -0.2897 | -2.5286 | -0.806 | -0.3096 | -2.794 |
| 3D absol volume | 0.5908 | 2.5286 | 0.8132 | 0.3385 | 2.8347 |

| 51 | 8 | 2 | lip, upper |
|---|---|---|---|

Sunken shave biopsy scar DO NOT USE

| 008DM L2 | V1 | V2 | V3 | V4 | V5 |
|---|---|---|---|---|---|
| Diameter (exam) | 4x4 | 4x4 | 4x4 | 4x4 | 4x4 |
| Height (exam) | flat | flat | sunken | flat | sunken |
| 3D volume | -7.4469 | -6.5487 | -6.957 | -6.4586 | -5.367 |
| 3D absol volume | 7.4469 | 6.5487 | 6.9572 | 6.4586 | 5.3669 |

| 52 | 8 | 3 | forearm |
|---|---|---|---|

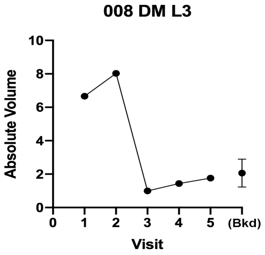

Raised follicular lesion

| 008DM L3 | V1 | V2 | V3 | V4 | V5 |
|---|---|---|---|---|---|
| Height (exam) | | | | | |
| Diameter (exam) | 5x5 | 5x5 | 3x3 | 3x3 | 3x3 |
| 3D diameter | 5.38 | 7.55 | 3.05 | -- | -- |
| 3D perp diameter | 4.89 | 6.34 | 2.33 | -- | -- |
| 3D av height | 0.185 | 0.219 | 0.011 | -- | -- |
| 3D Av Ht/Area * 10^4 | 22.340 | 14.418 | 4.963 | -- | -- |
| 3D volume | 6.280 | 7.369 | -0.098 | 1.350 | 1.725 |
| 3D absol volume | 6.660 | 8.027 | 0.996 | 1.441 | 1.759 |

**V3**

Background region at visit: **Average bkdg:**

| V1 | V2 | V5 | MEAN | SD |
|---|---|---|---|---|
| 2.819 | 2.209 | 1.169 | **2.066** | 0.834 |

| 53 | 8 | 4 | chest |
|---|---|---|---|

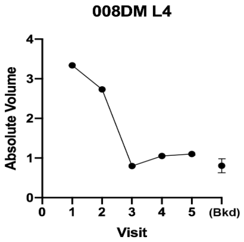

Raised lesion, gone by V3

| 008DM L4 | V1 | V2 | V3 | V4 | V5 |
|---|---|---|---|---|---|
| Height (exam) | raised | raised | flat | flat | flat |
| Diameter (exam) | 9x5 | 8x8 | 2x2 | 1x1 | 1x1 |
| 3D diameter | 6.89 | 7.86 | | | |
| 3D perp diameter | 4.35 | 6.78 | | | |
| 3D av height | 0.048 | 0.047 | | | |
| 3D Av Ht/Area * 10^4 | 4.809 | 2.783 | | | |
| 3D volume | -1.388 | 0.406 | 0.277 | -0.198 | 0.865 |
| 3D absol volume | 3.340 | 2.731 | 0.797 | 1.050 | 1.102 |

**V3**

Background region at visit: **Average bkdg:**

| V1 | V2 | V5 | MEAN | SD |
|---|---|---|---|---|
| 0.601 | 0.907 | 0.907 | **0.805** | 0.177 |

| Lesion Index # | Subject # | Lesion # | Body Location | Diagnosis by Biopsy |
|---|---|---|---|---|
| 54 | 8 | 5 | back | Biopsy at Visit 5: Pityrosporum folliculitis (no BCC) |

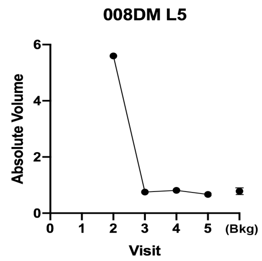

Raised lesion, gone by V3

| 008DM L5 | V1 | V2 | V3 | V4 | V5 |
|---|---|---|---|---|---|
| Height (exam) | flat | raised | flat | flat | flat |
| Diameter (exam) | 10x5 | 15x5 | 3x8 | 4x7 | 4x5 |
| 3D diameter | No photo | 15.38 | 6.22 | 2.44 | 3.45 |
| 3D perp diameter | | 5.99 | 4.60 | 2.24 | 3.09 |
| 3D av height | | 0.042 | 0.017 | 0.023 | 0.005 |
| 3D Av Ht/Area * 10^4 | | 1.173 | 1.899 | 13.191 | 1.600 |
| 3D volume | | -1.477 | 0.082 | 0.352 | 0.084 |
| 3D absol volume | | 5.597 | 0.751 | 0.813 | 0.667 |

Response outcome: **V3**

| Background region at visit: V1 | V2 | V5 | Average bkdg: MEAN | SD |
|---|---|---|---|---|
| 0.907 | 0.776 | 0.667 | **0.783** | 0.120 |

| Lesion Index # | Subject # | Lesion # | Body Location | Diagnosis by Biopsy |
|---|---|---|---|---|
| 55 | 8 | 6 | back | Biopsy at Visit 5: BCC, nodular |

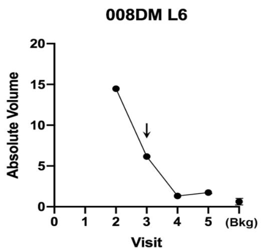

VitD = no

Large, barely-raised plaque at V2 that was still present at V5

| 008DM L6 | V1 | V2 | V3 | V4 | V5 |
|---|---|---|---|---|---|
| Height (exam) | | flat | raised | raised | flat |
| Diameter (exam) | | 10x4 | 10x4 | 10x4 | 10x4 |
| 3D diameter | No photo | 9.43 | 6.92 | 3.05 | 4.03 |
| 3D perp diameter | | 5.47 | 2.59 | 2.17 | 2.59 |
| 3D av height | | 0.028 | 0.132 | 0.133 | 0.138 |
| 3D Av Ht/Area * 10^4 | | 1.583 | 18.523 | 62.181 | 40.076 |
| 3D volume | | -11.977 | -3.538 | 1.125 | 1.205 |
| 3D absol volume | | 14.468 | 6.153 | 1.322 | 1.738 |

Response outcome: **NC**

| Background region at visit: V1 | V2 | V5 | Average bkdg: MEAN | SD |
|---|---|---|---|---|
| 0.606 | 0.669 | 0.614 | **0.630** | 0.034 |

| *Lesion Index #* | Subject # | Lesion # | Body Location | Diagnosis by Biopsy | Graphical data | Notes & Response to VitD | 3-D lesion data | Visit 1 | Visit 2 | Visit 3 | Visit 4 | Visit 5 | Response outcome | VDR gene |
|---|---|---|---|---|---|---|---|---|---|---|---|---|---|---|

**High-dose Vit D assignment: before V2**

Lengths are in (mm); Volumes in (mm^3)

**VDR alleles for patient 9**

| Ff | LS |
|---|---|

| Lesion Index # | Subject # | Lesion # | Body Location | Diagnosis by Biopsy |
|---|---|---|---|---|
| 56 | 9 | 1 | scalp | Biopsy at Visit 5: BCC, nodular |

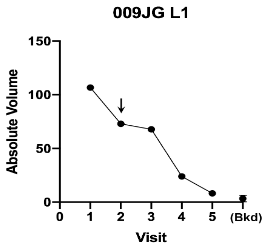

VitD=No

Big nodular BCC, had a good (but incomplete) response

| 009JG L1 | V1 | V2 | V3 | V4 | V5 |
|---|---|---|---|---|---|
| Height (exam) | raised | raised | raised | raised | raised |
| Diameter (exam) | 12x8 | 12x8 | 15x9 | 13x7 | 5x4 |
| 3D diameter | 13.98 | 12.83 | 13.53 | 12.73 | 7.83 |
| 3D perp diameter | 10.48 | 9.43 | 9.40 | 8.52 | 5.43 |
| 3D av height | 0.955 | 0.615 | 0.743 | 0.292 | 0.282 |
| 3D Av Ht/Area * 10^4 | 20.334 | 15.819 | 17.992 | 8.238 | 20.429 |
| 3D volume | 105.484 | 72.749 | 67.820 | 23.957 | 7.792 |
| 3D absol volume | 106.636 | 72.914 | 67.844 | 23.992 | 8.211 |

Response outcome: **NC**

| Background region at visit: V1 | V2 | V5 | Average bkdg: MEAN | SD |
|---|---|---|---|---|
| 3.301 | 5.335 | 1.428 | **3.355** | 1.954 |

| Lesion Index # | Subject # | Lesion # | Body Location | Diagnosis by Biopsy |
|---|---|---|---|---|
| 57 | 9 | 2 | scalp | Biopsy at Visit 5: BCC, nodular and micronodular |

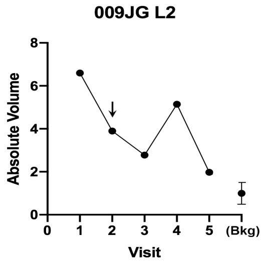

VitD=Yes

Small, hard to quantify; shrank but did not resolve completely

| 009JG L2 | V1 | V2 | V3 | V4 | V5 |
|---|---|---|---|---|---|
| Height (exam) | raised | raised | raised; s | raised | flat ± papules |
| Diameter (exam) | 11x9 | 11x9 | /papules | /papules | 7x8 |
| 3D diameter | 7.36 | 8.39 | 9.55 | 9.47 | 7.99 |
| 3D perp diameter | 6.22 | 7.19 | 7.63 | 4.98 | 7.57 |
| 3D av height | 0.105 | 0.064 | 0.032 | 0.100 | 0.102 |
| 3D Av Ht/Area * 10^4 | 7.256 | 3.382 | 1.368 | 6.077 | 5.356 |
| 3D volume | 6.450 | 3.807 | 1.299 | 5.036 | 1.884 |
| 3D absol volume | 6.599 | 3.897 | 2.777 | 5.142 | 1.977 |

Response outcome: **NC**

| Background region at visit: V1 | V2 | V5 | Average bkdg: MEAN | SD |
|---|---|---|---|---|
| 0.862 | 0.569 | 1.560 | **0.997** | 0.509 |

58 9 3 neck

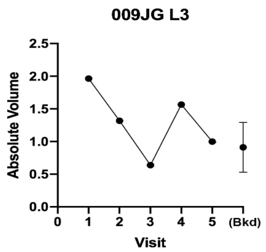

Visually gone at V3

| 009JG L3 | V1 | V2 | V3 | V4 | V5 |
|---|---|---|---|---|---|
| Height (exam) | raised | raised | flat | flat | normal skin |
| Diameter (exam) | 4x5 | 4x5 | 2x2 | 1x1 | 2x2 |
| 3D diameter | 9.36 | 6.39 | -- | -- | -- |
| 3D perp diameter | 5.20 | 4.89 | -- | -- | -- |
| 3D av height | 0.021 | 0.022 | -- | -- | -- |
| 3D Av Ht/Area * 10^4 | 1.279 | 2.246 | -- | -- | -- |
| 3D volume | -0.520 | -0.440 | -0.494 | -1.161 | -0.803 |
| 3D absol volume | 1.964 | 1.317 | 0.640 | 1.566 | 0.998 |

**V3**

Background region at visit: **Average bkdg:**

| V1 | V2 | V5 | MEAN | SD |
|---|---|---|---|---|
| 0.919 | 1.289 | 0.525 | **0.911** | 0.382 |

59 9 4 neck Biopsy at Visit 5:

BCC, nodular and micronodular

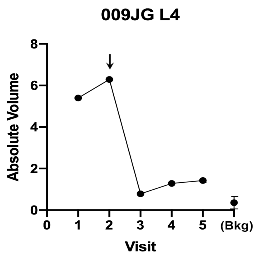

VitD=Yes

| 009JG L4 | V1 | V2 | V3 | V4 | V5 |
|---|---|---|---|---|---|
| Height (exam) | raised | slightly ra | slightly r | slightly ra | slightly raised |
| Diameter (exam) | 5x5 | 5x5 | 2x2 | 2x2 | 2x2 |
| 3D diameter | 7.22 | 7.65 | 6.74 | 4.60 | 4.36 |
| 3D perp diameter | 4.52 | 4.12 | 3.91 | 2.97 | 2.99 |
| 3D av height | 0.240 | 0.314 | 0.029 | 0.135 | 0.168 |
| 3D Av Ht/Area * 10^4 | 22.186 | 28.921 | 3.308 | 29.942 | 39.512 |
| 3D volume | 5.340 | 6.277 | 0.163 | 1.123 | 1.392 |
| 3D absol volume | 5.398 | 6.287 | 0.783 | 1.286 | 1.422 |

**NC**

Background region at visit: **Average bkdg:**

| V1 | V2 | V5 | MEAN | SD |
|---|---|---|---|---|
| 0.345 | 0.445 | 0.270 | **0.353** | 0.088 |

60 9 5 cheek

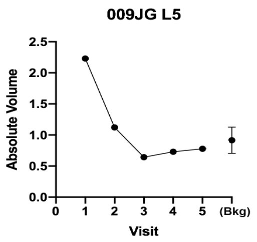

Small lesion, almost gone at V2, completely gone at V3

| 009JG L5 | V1 | V2 | V3 | V4 | V5 |
|---|---|---|---|---|---|
| Height (exam) | raised | raised, rai | none, rai | none, no | none, none |
| Diameter (exam) | 3x3 | 3x3 | 1x2 | No lesion | No lesion |
| 3D diameter | 7.29 | 4.13 | -- | -- | -- |
| 3D perp diameter | 6.07 | 3.25 | -- | -- | -- |
| 3D av height | 0.101 | 0.047 | -- | -- | -- |
| 3D Av Ht/Area * 10^4 | 7.183 | 11.055 | -- | -- | -- |
| 3D volume | 2.212 | 1.111 | 0.418 | 0.263 | 0.606 |
| 3D absol volume | 2.231 | 1.123 | 0.642 | 0.731 | 0.778 |

**V3**

Background region at visit: **Average bkdg:**

| V1 | V2 | V5 | MEAN | SD |
|---|---|---|---|---|
| 1.009 | 0.674 | 1.064 | **0.916** | 0.211 |

61 9 6 back Biopsy at Visit 5:

BCC, micronodular

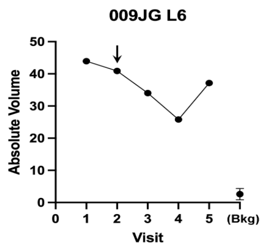

VitD=No

BCC micronodular by biopsy

| 009JG L6 | V1 | V2 | V3 | V4 | V5 |
|---|---|---|---|---|---|
| Height (exam) | soft raise | soft raise | soft raise | soft raise | soft raised |
| Diameter (exam) | 10x10 | 10x10 | 7x11 | 7x11 | 9x12 |
| 3D diameter | 11.52 | 10.62 | 10.96 | 10.83 | 11.79 |
| 3D perp diameter | 9.74 | 9.62 | 9.20 | 8.93 | 9.68 |
| 3D av height | 0.570 | 0.608 | 0.482 | 0.379 | 0.473 |
| 3D Av Ht/Area * 10^4 | nd | nd | nd | nd | nd |
| 3D volume | 43.835 | 40.904 | 34.001 | 25.800 | 37.081 |
| 3D absol volume | 43.923 | 40.905 | 34.001 | 25.813 | 37.130 |

**NC**

Background region at visit: **Average bkdg:**

| V1 | V2 | V5 | MEAN | SD |
|---|---|---|---|---|
| 2.458 | 2.521 | 2.845 | **2.608** | 0.208 |

62 9 7 chest Biopsy at Visit 5:

Benign lichenoid keratosis

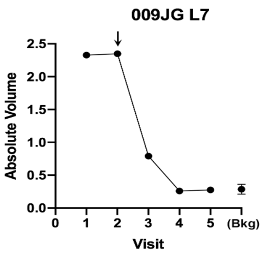

VitD=Yes

Completely gone at V4

| 009JG L7 | V1 | V2 | V3 | V4 | V5 |
|---|---|---|---|---|---|
| Height (exam) | raised | raised | raised | flat | flat |
| Diameter (exam) | 4x5 | 4x5 | 3x3 | 3x3 | 2x2 |
| 3D diameter | 4.75 | 4.48 | 5.61 | 3.12 | 4.26 |
| 3D perp diameter | 4.48 | 4.32 | 3.36 | 2.87 | 3.40 |
| 3D av height | 0.177 | 0.185 | 0.063 | 0.009 | 0.015 |
| 3D Av Ht/Area * 10^4 | 26.472 | 30.312 | 9.974 | 3.086 | 3.304 |
| 3D volume | 2.307 | 2.339 | 0.761 | 0.132 | 0.223 |
| 3D absol volume | 2.327 | 2.348 | 0.790 | 0.259 | 0.276 |

**V4**

Background region at visit: **Average bkdg:**

| V1 | V2 | V5 | MEAN | SD |
|---|---|---|---|---|
| 0.337 | 0.265 | 0.259 | **0.287** | 0.043 |

| Lesion Index # | Subject # | Lesion # | Body Location | Diagnosis by Biopsy |
|---|---|---|---|---|
| *63* | 9 | 8 | chest | |

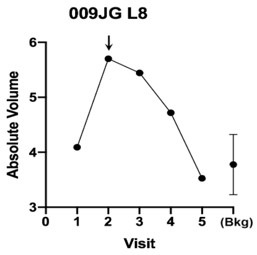

VitD=no

Big superficial BCC; visually gone at V4, digitally gone at V5

| 009JG L8 | V1 | V2 | V3 | V4 | V5 |
|---|---|---|---|---|---|
| Height (exam) | slight rais | scaly raise | nearly fla | nearly fla | nearly flat |
| Diameter (exam) | 18x15 | 18x15 | 18x10 | 14x15 | invisible |
| 3D diameter | 17.827 | 18.051 | 16.520 | 17.817 | 19.095 |
| 3D perp diameter | 11.986 | 12.521 | 13.289 | 12.219 | 13.467 |
| 3D av height | 0.025 | 0.021 | 0.014 | 0.015 | 0.019 |
| 3D Av Ht/Area * 10^4 | 0.356 | 0.281 | 0.195 | 0.209 | 0.228 |
| 3D volume | 2.298 | -0.935 | -2.441 | -1.107 | 0.533 |
| 3D absol volume | 4.091 | 5.701 | 5.443 | 4.718 | 3.526 |

Response outcome: **V5**

| Background region at visit: | | | Average bkdg: | |
|---|---|---|---|---|
| V1 | V2 | V5 | MEAN | SD |
| 3.430 | 3.490 | 4.409 | **3.777** | 0.549 |

| Lesion Index # | Subject # | Lesion # | Body Location | Diagnosis by Biopsy |
|---|---|---|---|---|
| *64* | 9 | 9 | abdomen | Biopsy at Visit 5: BCC, superficial |

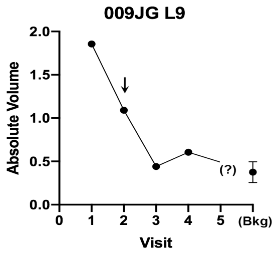

VitD=Yes

Lesion still raised at V4 (and at V5 per clinical notes). Biopsied.

| 009JG L9 | V1 | V2 | V3 | V4 | V5 |
|---|---|---|---|---|---|
| Height (exam) | flat | flat | flat | flat | flat |
| Diameter (exam) | 4x4 | 4x4 | 2x4 | 2x4 | 3x4 |
| 3D diameter | 7.19 | 7.90 | 5.55 | 7.65 | |
| 3D perp diameter | 5.81 | 5.27 | 3.47 | 4.40 | No |
| 3D av height | 0.061 | 0.031 | 0.027 | 0.025 | Photo |
| 3D Av Ht/Area * 10^4 | 4.598 | 2.309 | 4.195 | 2.227 | |
| 3D volume | 1.830 | 0.788 | 0.300 | 0.510 | |
| 3D absol volume | 1.857 | 1.091 | 0.440 | 0.605 | |

Response outcome: **NC**

| Background region at visit: | | | Average bkdg: | |
|---|---|---|---|---|
| V1 | V2 | V5 | MEAN | SD |
| 0.305 | 0.516 | 0.309 | **0.376** | 0.121 |

| Lesion Index # | Subject # | Lesion # | Body Location | Diagnosis by Biopsy |
|---|---|---|---|---|
| *65* | 9 | 10 | back | |

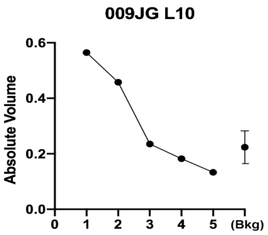

Very small lesion, gone at V3

| 009JG L10 | V1 | V2 | V3 | V4 | V5 |
|---|---|---|---|---|---|
| Height (exam) | flat | flat | flat | flat | flat, faint pink |
| Diameter (exam) | 4x6 | 4x6 | 4x6 | 2x2 | 2x2 |
| 3D diameter | 8.44 | 5.15 | 3.64 | 3.00 | -- |
| 3D perp diameter | 3.40 | 2.96 | 1.62 | 1.61 | -- |
| 3D av height | 0.026 | 0.010 | 0.028 | 0.011 | -- |
| 3D Av Ht/Area * 10^4 | 2.325 | 1.874 | 12.923 | 6.494 | -- |
| 3D volume | 0.244 | 0.032 | 0.054 | 0.088 | 0.006 |
| 3D absol volume | 0.565 | 0.458 | 0.235 | 0.182 | 0.133 |

Response outcome: **V3**

| Background region at visit: | | | Average bkdg: | |
|---|---|---|---|---|
| V1 | V2 | V5 | MEAN | SD |
| 0.290 | 0.178 | 0.202 | **0.224** | 0.059 |

| *Lesion Index #* | Subject # | Lesion # | Body Location | Diagnosis by Biopsy | Graphical data | Notes & Response to VitD | 3-D lesion data | Visit 1 | Visit 2 | Visit 3 | Visit 4 | Visit 5 | Response outcome | VDR gene |
|---|---|---|---|---|---|---|---|---|---|---|---|---|---|---|

**High-dose Vit D assignment: before V3**

Lengths are in (mm); Volumes in (mm^3)

**VDR alleles for patient 10**

| FF | LL |
|---|---|

| Lesion Index # | Subject # | Lesion # | Body Location | Diagnosis by Biopsy |
|---|---|---|---|---|
| *66* | 10 | 1 | temple | |

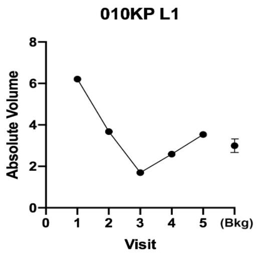

Completely gone at V3

| 010KP L1 | V1 | V2 | V3 | V4 | V5 |
|---|---|---|---|---|---|
| Height (exam) | slightly ra | slightly ra | flat | flat | flat, hard to find |
| Diameter (exam) | 9x10 | 9x10 | 9x10 | 5x8 | 5x8 |
| 3D diameter | 12.01 | 9.23 | -- | -- | -- |
| 3D perp diameter | 9.85 | 7.15 | -- | -- | -- |
| 3D av height | 0.068 | 0.023 | -- | -- | -- |
| 3D Av Ht/Area * 10^4 | 1.820 | 1.094 | -- | -- | -- |
| 3D volume | 5.173 | 2.663 | -0.501 | 1.823 | 3.363 |
| 3D absol volume | 6.212 | 3.676 | 1.699 | 2.588 | 3.534 |

Response outcome: **V3**

| Background region at visit: | | | Average bkdg: | |
|---|---|---|---|---|
| V1 | V2 | V5 | MEAN | SD |
| 2.633 | 3.087 | 3.280 | **3.000** | 0.332 |

67 | 10 | 2 | temple

Biopsy at Visit 5:

Biopsy negative for residual

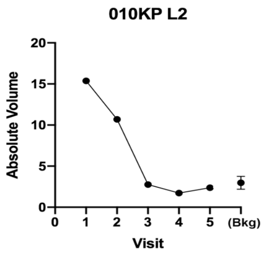

Still erythema V4; all gone at V5

| 010KP L2 | V1 | V2 | V3 | V4 | V5 |
|---|---|---|---|---|---|
| Height (exam) | slightly ra | slightly ra | raised w, flat | | flat |
| Diameter (exam) | 8x15 | 8x15 | 8x15 | 6x8 | 6x8 |
| 3D diameter | 16.11 | 21.12 | 15.97 | -- | -- |
| 3D perp diameter | 7.56 | 11.51 | 8.49 | -- | -- |
| 3D av height | 0.019 | 0.063 | 0.008 | -- | -- |
| 3D Av Ht/Area * 10^4 | 0.422 | 0.756 | 0.177 | -- | -- |
| 3D volume | 10.984 | 7.556 | -1.634 | 0.398 | 1.883 |
| 3D absol volume | 15.372 | 10.692 | 2.763 | 1.721 | 2.376 |

V3

| Background region at visit: | | | Average bkdg: | |
|---|---|---|---|---|
| V1 | V2 | V5 | MEAN | SD |
| 3.860 | 2.386 | 2.662 | **2.970** | 0.783 |

68 | 10 | 3 | cheek

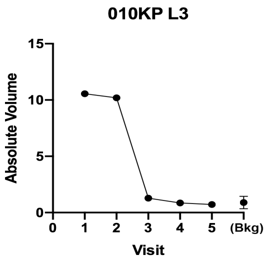

Lesion very unclear in pictures
Gone at V3

| 010KP L3 | V1 | V2 | V3 | V4 | V5 |
|---|---|---|---|---|---|
| Height (exam) | slightly ra | two raise | flat | no visible | no visible lesion |
| Diameter (exam) | 6x12 | 11x20 | 11x20 | 11x20 | 11x20 |
| 3D diameter | 13.72 | 13.72 | -- | -- | -- |
| 3D perp diameter | 5.56 | 3.52 | -- | -- | -- |
| 3D av height | 0.072 | 0.046 | -- | -- | -- |
| 3D Av Ht/Area * 10^4 | 2.467 | 1.956 | -- | -- | -- |
| 3D volume | 9.087 | 8.800 | 1.068 | 0.709 | 0.188 |
| 3D absol volume | 10.551 | 10.199 | 1.281 | 0.862 | 0.714 |

V3

| Background region at visit: | | | Average bkdg: | |
|---|---|---|---|---|
| V1 | V2 | V5 | MEAN | SD |
| 1.287 | 0.764 | 0.613 | **0.888** | 0.354 |

69 | 10 | 4 | mid back

Biopsy at Visit 5:

Normal skin

In retrospect, this raised lesion was never a BCC. In the images it has normal epidermal markings, even though slightly raised.

| 010KP L4 | V1 | V2 | V3 | V4 | V5 |
|---|---|---|---|---|---|
| Diameter (exam) | 8x8 | 8x8 | 8x8 | 8x8 | 8x8 |
| Height (exam) | slightly ra | slightly ra | slightly r | slightly ra | barely |
| 3D volume | 7.6151 | 8.237 | 4.1949 | 3.5581 | 1.3595 |
| 3D absol volume | 7.6151 | 8.28 | 4.2131 | 3.6211 | 1.3778 |

| on V1 | l on V2 | on V5 | **Average bkg controls:** | |
|---|---|---|---|---|
| | | | MEAN | SD |
| 0.7133 | 0.913 | 0.86425 | **0.83** | 0.1041 |

70 | 10 | 5 | temple

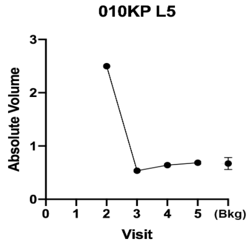

Completely gone at V3

| 010KP L5 | V1 | V2 | V3 | V4 | V5 |
|---|---|---|---|---|---|
| Height (exam) | | raised | no lesior | no lesion | no lesion detected |
| Diameter (exam) | | 3x4 | 3x4 | 3x4 | 3x4 |
| 3D diameter | | 6.19 | -- | -- | -- |
| 3D perp diameter | | 5.69 | -- | -- | -- |
| 3D av height | | 0.089 | -- | -- | -- |
| 3D Av Ht/Area * 10^4 | | 8.036 | -- | -- | -- |
| 3D volume | | 2.343 | 0.176 | -0.429 | 0.392 |
| 3D absol volume | | 2.498 | 0.537 | 0.641 | 0.687 |

V3

| Background region at visit: | | | Average bkdg: | |
|---|---|---|---|---|
| V1 | V2 | V5 | MEAN | SD |
| 0.540 | 0.737 | 0.737 | **0.671** | 0.113 |

71 | 10 | 6 | temple

Question whether BCC, as lesion looks different at V1 than V2 (dry, scarlike papules).
DO NOT USE.

| 010KP L6 | V1 | V2 | V3 | V4 | V5 | |
|---|---|---|---|---|---|---|
| Diameter (exam) | | 4x5 | 4x5 | 4x5 | 4x5 | |
| Height (exam) | | raised | flat | no visible | no visibl | ntr |
| 3D volume | 1.6002 | 3.6768 | 8.6509 | 5.4759 | 2.7451 | |
| 3D absol volume | 1.6252 | 3.6768 | 8.6509 | 5.4923 | 2.7571 | |

on V1 l on V2 on V5

72 | 10 | 7 | neck

Lesion only appeared at Visit 4
DO NOT USE.

| 010KP L7 | V1 | V2 | V3 | V4 | V5 | |
|---|---|---|---|---|---|---|
| Diameter (exam) | | | | 6x5 | 6x5 | |
| Height (exam) | | | | slightly ra | no visibl | ntr |
| 3D volume | | | | 3.7936 | | |
| 3D absol volume | | | | 4.1853 | | |

on V1 l on V2 on V5

| *Lesion Index #* | Subject # | Lesion # | Body Location | Diagnosis by Biopsy | Graphical data | Notes & Response to VitD | 3-D lesion data | Visit 1 | Visit 2 | Visit 3 | Visit 4 | Visit 5 | Response outcome | VDR gene |
|---|---|---|---|---|---|---|---|---|---|---|---|---|---|---|

**11- Screenfail**

| *Lesion Index #* | Subject # | Lesion # | Body Location | Diagnosis by Biopsy | Graphical data | Notes & Response to VitD | 3-D lesion data | Visit 1 | Visit 2 | Visit 3 | Visit 4 | Visit 5 | Response outcome | VDR gene |
|---|---|---|---|---|---|---|---|---|---|---|---|---|---|---|

**High-dose Vit D assignment: before V2**

Lengths are in (mm); Volumes in (mm^3)

VDR alleles: Ff | SS

*73* | **12** | **1** | shoulder

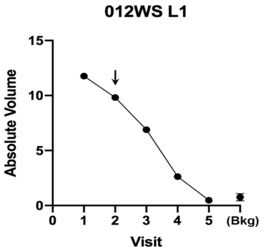

VitD = No

| 012WS L1 | V1 | V2 | V3 | V4 | V5 |
|---|---|---|---|---|---|
| Height (exam) | flat | flat | flat | flat | gone |
| Diameter (exam) | 10x13 | 10x13 | 10x13 | | No lesion |
| 3D diameter | 17.12 | 15.39 | 7.45 | 6.04 | visible |
| 3D perp diameter | 13.74 | 11.90 | 5.43 | 2.84 | -- |
| 3D av height | 0.045 | 0.030 | 0.179 | 0.050 | -- |
| 3D Av Ht/Area * 10^4 | 0.605 | 0.5085 | 13.742 | 8.103 | |
| 3D volume | 10.267 | 6.926 | 3.986 | 0.754 | 0.112 |
| 3D absol volume | 11.767 | 9.815 | 6.893 | 2.637 | 0.475 |

Visually and digitally gone by V5

**V5**

| Background region at visit: | | | Average bkdg: | |
|---|---|---|---|---|
| V1 | V2 | V5 | MEAN | SD |
| 0.659 | 1.121 | 0.475 | **0.752** | 0.333 |

*74* | **12** | **2** | midback | Biopsy at Visit 5: Scar tissue

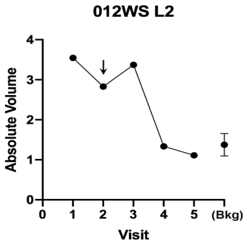

VitD = No

| 012WS L2 | V1 | V2 | V3 | V4 | V5 |
|---|---|---|---|---|---|
| Height (exam) | flat | flat | flat | gone | |
| Diameter (exam) | 10x10 | 10x10 | 7x10 | | |
| 3D diameter | 11.81 | 11.32 | 5.16 | 1.99 | No lesion |
| 3D perp diameter | 9.92 | 9.33 | 3.12 | 1.47 | visible |
| 3D av height | 0.038 | 0.040 | 0.054 | 0.001 | -- |
| 3D Av Ht/Area * 10^4 | 1.027 | 1.182 | 10.031 | 1.538 | -- |
| 3D volume | 2.276 | 2.253 | 0.866 | 0.891 | -0.280 |
| 3D absol volume | 3.544 | 2.828 | 3.373 | 1.334 | 1.110 |

visually gone by V4
Scar by biopsy

**V4**

| Background region at visit: | | | Average bkdg: | |
|---|---|---|---|---|
| V1 | V2 | V5 | MEAN | SD |
| 1.641 | 1.399 | 1.080 | **1.373** | 0.281 |

*75* | **12** | **3** | shoulder

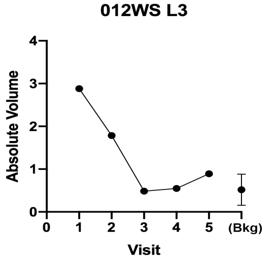

| 012WS L3 | V1 | V2 | V3 | V4 | V5 |
|---|---|---|---|---|---|
| Height (exam) | flat | flat | flat | gone | |
| Diameter (exam) | 9x10 | 9x10 | 3x5 | | |
| 3D diameter | 10.40 | 10.00 | No lesion | No lesion | No lesion |
| 3D perp diameter | 7.65 | 9.41 | visible | visible | visible |
| 3D av height | 0.045 | 0.033 | -- | -- | -- |
| 3D Av Ht/Area * 10^4 | 1.777 | 1.099 | -- | -- | -- |
| 3D volume | 2.588 | 1.674 | 0.307 | -0.357 | -0.713 |
| 3D absol volume | 2.880 | 1.784 | 0.484 | 0.548 | 0.890 |

Visually gone by V3

**V3**

| Background region at visit: | | | Average bkdg: | |
|---|---|---|---|---|
| V1 | V2 | V5 | MEAN | SD |
| 0.376 | 0.249 | 0.931 | **0.519** | 0.362 |

*76* | **12** | **4** | shoulder

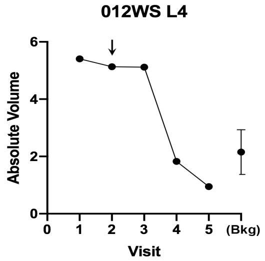

VitD = No

| 012WS L4 | V1 | V2 | V3 | V4 | V5 |
|---|---|---|---|---|---|
| Height (exam) | flat | flat | flat | gone | |
| Diameter (exam) | 15x14 | 15x14 | 8x8 | | |
| 3D diameter | 13.13 | 14.15 | 8.59 | No lesion | No lesion |
| 3D perp diameter | 12.13 | 13.35 | 7.36 | visible | visible |
| 3D av height | 0.047 | 0.011 | 0.059 | -- | -- |
| 3D Av Ht/Area * 10^4 | 0.935 | 0.187 | 2.939 | -- | -- |
| 3D volume | 4.236 | -2.514 | -2.469 | 0.696 | 0.331 |
| 3D absol volume | 5.410 | 5.132 | 5.116 | 1.829 | 0.949 |

visually & digitally gone by V4

**V4**

| Background region at visit: | | | Average bkdg: | |
|---|---|---|---|---|
| V1 | V2 | V5 | MEAN | SD |
| 2.972 | 2.064 | 1.417 | **2.151** | 0.781 |

| | | | |
|---|---|---|---|
| 77 | 12 | 5 | back |

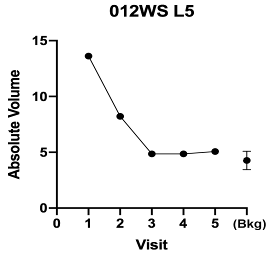

Puzzling lesion (falt, scaly, shiny); became drier amd more fibrotic in appearance.
Visually, lesion resolved at V3-V4

| 012WS L5 | V1 | V2 | V3 | V4 | V5 |
|---|---|---|---|---|---|
| Height (exam) | flat | flat | flat | flat | Normal skin markings |
| Diameter (exam) | 15x12 | 15x12 | 10x12 | 8x10 | 8x10 |
| 3D diameter | 16.57 | 17.06 | 6.69 | No lesion | No lesion |
| 3D perp diameter | 14.88 | 13.28 | 3.50 | visible | visible |
| 3D av height | 0.071 | 0.027 | 0.040 | -- | -- |
| 3D Av Ht/Area * 10^4 | 0.915 | 0.380 | 4.960 | -- | -- |
| 3D volume | 9.276 | -0.292 | -2.513 | -4.295 | -0.637 |
| 3D absol volume | 13.617 | 8.216 | 4.860 | 4.860 | 5.074 |

**V3**

| Background region at visit: | | | Average bkdg: | |
|---|---|---|---|---|
| V1 | V2 | V5 | MEAN | SD |
| 4.626 | 3.327 | 4.870 | **4.274** | 0.829 |

| | | | |
|---|---|---|---|
| 78 | 12 | 6 | back |

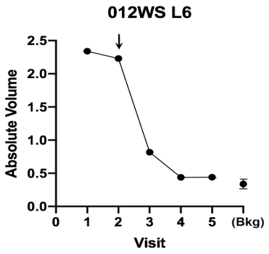

VitD=Yes

Nearly flat, still visible at V3; fibrotic patch at V4, with normal normal epidermis at v4 and V5

| 012WS L6 | V1 | V2 | V3 | V4 | V5 |
|---|---|---|---|---|---|
| Height (exam) | flat | flat | flat | flat | Normal skin markings |
| Diameter (exam) | 11x8 | 11x8 | 7x10 | 6x7 | 8x7 |
| 3D diameter | 11.47 | 10.88 | 1.94 | No lesion | No lesion |
| 3D perp diameter | 8.15 | 7.81 | 1.86 | visible | visible |
| 3D av height | 0.0154 | 0.0184 | 0.0074 | -- | -- |
| 3D Av Ht/Area * 10^4 | 0.509 | 0.669 | 6.584 | -- | -- |
| 3D volume | 0.153 | -0.100 | -0.676 | -0.254 | -0.344 |
| 3D absol volume | 2.341 | 2.230 | 0.818 | 0.439 | 0.440 |

**V4**

| Background region at visit: | | | Average bkdg: | |
|---|---|---|---|---|
| V1 | V2 | V5 | MEAN | SD |
| 0.356 | 0.365 | 0.297 | **0.339** | 0.037 |

| | | | |
|---|---|---|---|
| 79 | 12 | 7 | back |

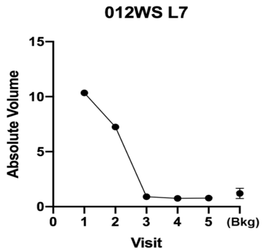

Visually gone by V3 (atrophic)

| 012WS L7 | V1 | V2 | V3 | V4 | V5 |
|---|---|---|---|---|---|
| Height (exam) | flat | flat | flat | slightly d | Slightly depressed |
| Diameter (exam) | 11x7 | 11x7 | 8x11 | 6x9 | 6x9 |
| 3D diameter | 13.48 | 12.80 | No lesion | No lesion | No lesion |
| 3D perp diameter | 10.04 | 9.77 | visible | visible | visible |
| 3D av height | 0.0216 | 0.0279 | -- | -- | -- |
| 3D Av Ht/Area * 10^4 | 0.498 | 0.697 | -- | -- | -- |
| 3D volume | -6.895 | -3.455 | 0.091 | 0.083 | 0.652 |
| 3D absol volume | 10.346 | 7.238 | 0.906 | 0.755 | 0.778 |

**V3**

| Background region at visit: | | | Average bkdg: | |
|---|---|---|---|---|
| V1 | V2 | V5 | MEAN | SD |
| 1.696 | 1.138 | 0.774 | **1.203** | 0.465 |

| | | | |
|---|---|---|---|
| 80 | 12 | 8 | back |

Biopsy at Visit 5:
Scar tissue

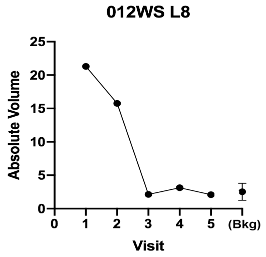

At V1, V2, resembles flat Bx scar with rim of BCC; all gone by V3

| 012WS L8 | V1 | V2 | V3 | V4 | V5 |
|---|---|---|---|---|---|
| Height (exam) | flat | flat | flat | flat | 3x3 slightly raised |
| Diameter (exam) | 15x9 | 15x9 | 7x10 | 7x10 | 7x10 |
| 3D diameter | 16.21 | 16.40 | 12.76 | No lesion | ?Scar |
| 3D perp diameter | 11.75 | 11.93 | 8.87 | visible | -- |
| 3D av height | 0.0065 | 0.0148 | 0.0048 | -- | -- |
| 3D Av Ht/Area * 10^4 | 0.106 | 0.234 | 0.131 | -- | -- |
| 3D volume | -19.531 | -12.004 | -0.760 | 0.963 | 1.536 |
| 3D absol volume | 21.305 | 15.753 | 2.142 | 3.134 | 2.094 |

**V3**

| Background region at visit: | | | Average bkdg: | |
|---|---|---|---|---|
| V1 | V2 | V5 | MEAN | SD |
| 3.984 | 1.993 | 1.594 | **2.524** | 1.280 |

| | | | |
|---|---|---|---|
| 81 | 12 | 9 | leg |

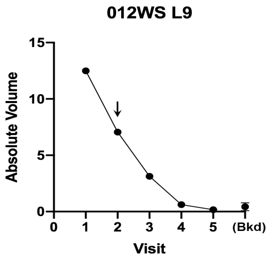

VitD = no

Flat scaly lesion; still there at V3; visually gone by V4

| 012WS L9 | V1 | V2 | V3 | V4 | V5 |
|---|---|---|---|---|---|
| Height (exam) | flat | flat | flat | flat | normal skin |
| Diameter (exam) | 5x8 | 5x8 | 11x9 | 4x5 | 4x5 |
| 3D diameter | 12.12 | 11.81 | 11.20 | No lesion | No lesion |
| 3D perp diameter | 8.80 | 8.92 | 9.09 | visible | visible |
| 3D av height | 0.1601 | 0.0971 | 0.0522 | -- | -- |
| 3D Av Ht/Area * 10^4 | 4.655 | 2.878 | 1.615 | -- | -- |
| 3D volume | 12.406 | 6.953 | 2.965 | 0.607 | 0.169 |
| 3D absol volume | 12.498 | 7.057 | 3.136 | 0.615 | 0.172 |

**V4**

| Background region at visit: | | | Average bkdg: | |
|---|---|---|---|---|
| V1 | V2 | V5 | MEAN | SD |
| 0.573 | 0.623 | 0.084 | **0.427** | 0.298 |

82 | 12 | 10 | leg

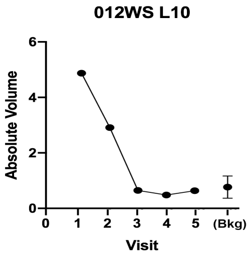

Visually, lesion is gone at V3
normal epidermis and skin color

| 012WS L10 | V1 | V2 | V3 | V4 | V5 |
|---|---|---|---|---|---|
| Height (exam) | flat | flat | flat | flat | normal skin |
| Diameter (exam) | 9x5 | 9x5 | 4x5 | 5x10 | 5x10 |
| 3D diameter | 10.50 | 10.04 | 8.51 | 4.41 | No lesion |
| 3D perp diameter | 6.40 | 8.10 | 5.16 | 2.46 | visible |
| 3D av height | 0.0999 | 0.0488 | 0.0161 | 0.0076 | -- |
| 3D Av Ht/Area * 10^4 | 4.456 | 1.886 | 1.094 | 2.063 | -- |
| 3D volume | 4.739 | 2.037 | 0.392 | -0.147 | 0.509 |
| 3D absol volume | 4.855 | 2.890 | 0.629 | 0.463 | 0.620 |

Response outcome: V3

Background region at visit: Average bkdg:

| V1 | V2 | V5 | MEAN | SD |
|---|---|---|---|---|
| 1.214 | 0.529 | 0.502 | **0.749** | 0.40 |

| *Lesion Index #* | Subject # | Lesion # | Body Location | Diagnosis by Biopsy | Graphical data | Notes & Response to VitD | 3-D lesion data | Visit 1 | Visit 2 | Visit 3 | Visit 4 | Visit 5 | Response outcome | VDR gene |
|---|---|---|---|---|---|---|---|---|---|---|---|---|---|---|
| | | | | | | **High-dose Vit D assignment: before V2** | | | | | | | | |

***In retrospect, this patient's lesions were all keloids (located on shaven legs).*** *In retrospect, this patient's lesions were keloids (all located on shaven legs).*

83 | 13 | 1 | leg, pretibial

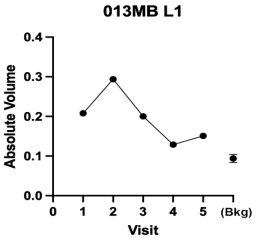

Scar/keloid

Small papule, still present at V5

| 013MB L1 | V1 | V2 | V3 | V4 | V5 |
|---|---|---|---|---|---|
| Diameter (exam) | 3x3 | 2x2 | 2x2 | 1x1 | |
| Height (exam) | Raised | raised | raised | raised | raised |
| 3D diameter | 3.181 | 2.716 | 2.664 | 2.574 | 2.344 |
| 3D perp diameter | 2.380 | 2.504 | 2.037 | 2.209 | 1.919 |
| 3D av height | 0.049 | 0.081 | 0.068 | 0.044 | 0.061 |
| 3D Av Ht/Area * 10^4 | 27.890 | 38.000 | 39.360 | 25.580 | 29.170 |
| 3D volume | 0.208 | 0.294 | 0.199 | 0.129 | 0.151 |
| 3D absol volume | 0.208 | 0.294 | 0.200 | 0.129 | 0.151 |

**VDR alleles:**

| Ff | LL |
|---|---|

Background region at visit: Average bkdg:

| V1 | V2 | V4 | MEAN | SD |
|---|---|---|---|---|
| 0.1 | 0.083 | 0.1 | **0.094** | 0.01 |

84 | 13 | 2 | leg, pretibial | Biopsy at Visit 5: Biopsy: Scar

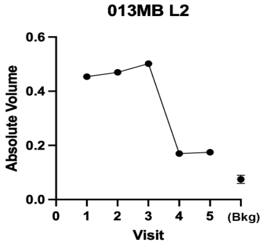

Scar/keloid (Bx-proven)

Small papule, still present at V5

| 013MB L2 | V1 | V2 | V3 | V4 | V5 |
|---|---|---|---|---|---|
| Diameter (exam) | 3x3 | 3x2 | 3x2 | 2x2 | |
| Height (exam) | Raised | raised | raised | raised | raised |
| 3D diameter | 3.115 | 3.487 | 3.277 | 2.875 | 3.116 |
| 3D perp diameter | 2.768 | 3.015 | 2.682 | 2.540 | 2.448 |
| 3D av height | 0.094 | 0.076 | 0.095 | 0.044 | 0.037 |
| 3D Av Ht/Area * 10^4 | 34.620 | 22.740 | 34.180 | 18.940 | 14.170 |
| 3D volume | 0.452 | 0.464 | 0.502 | 0.170 | 0.173 |
| 3D absol volume | 0.454 | 0.470 | 0.502 | 0.170 | 0.175 |

Background region at visit: Average bkdg:

| V1 | V4 | V5 | MEAN | SD |
|---|---|---|---|---|
| 0.072 | 0.067 | 0.086 | **0.075** | 0.01 |

85 | 13 | 3 | leg, pretibial

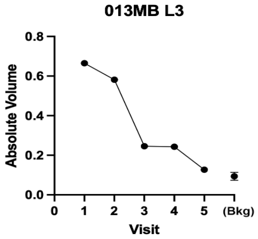

Scar/keloid

Small papule, still present at V5

| 013MB L3 | V1 | V2 | V3 | V4 | V5 |
|---|---|---|---|---|---|
| Diameter (exam) | 3x3 | 3x2 | 3x2 | 2x2 | |
| Height (exam) | Raised | raised | raised | raised | raised |
| 3D diameter | 3.416 | 3.526 | 3.158 | 2.915 | 3.010 |
| 3D perp diameter | 2.929 | 3.092 | 2.598 | 2.635 | 2.610 |
| 3D av height | 0.111 | 0.082 | 0.050 | 0.062 | 0.027 |
| 3D Av Ht/Area * 10^4 | 35.150 | 23.970 | 19.360 | 25.640 | 11.050 |
| 3D volume | 0.663 | 0.563 | 0.245 | 0.243 | 0.120 |
| 3D absol volume | 0.665 | 0.582 | 0.246 | 0.243 | 0.127 |

Background region at visit: Average bkdg:

| V1 | V4 | V5 | MEAN | SD |
|---|---|---|---|---|
| 0.1 | 0.083 | 0.1 | **0.094** | 0.01 |

| | | | | |
|---|---|---|---|---|
| 86 | 13 | 4 | leg, pretibial | Scar/keloid |

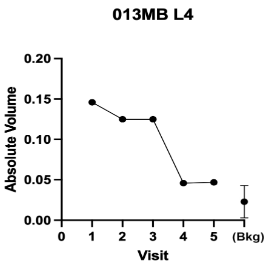

| 013MB L4 | V1 | V2 | V3 | V4 | V5 | | | | | |
|---|---|---|---|---|---|---|---|---|---|---|
| Diameter (exam) | 1x1 | 1x2 | 1x1 | 1x1 | | | | | | |
| Height (exam) | Raised | raised | raised | raised | raised | | | | | |
| 3D diameter | 2.239 | 2.653 | 2.525 | 1.799 | 1.854 | | | | | |
| 3D perp diameter | 1.785 | 1.729 | 1.983 | 1.570 | 1.643 | | | | | |
| 3D av height | 0.065 | 0.049 | 0.037 | 0.034 | 0.031 | | | | | |
| 3D Av Ht/Area * 10^4 | 51.100 | 32.210 | 22.960 | 37.770 | 31.830 | Background region at visit: | | | Average bkdg: | |
| 3D volume | 0.119 | 0.123 | 0.051 | 0.046 | 0.046 | V1 | V4 | V5 | MEAN | SD |
| 3D absol volume | 0.146 | 0.125 | 0.125 | 0.046 | 0.047 | 0.026 | 0.025 | 0.018 | **0.023** | 0.00 |

| | | | | |
|---|---|---|---|---|
| 87 | 13 | 5 | leg, pretibial | Scar/keloid |

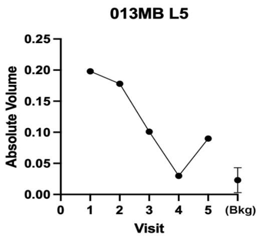

| 013MB L5 | V1 | V2 | V3 | V4 | V5 | | | | | |
|---|---|---|---|---|---|---|---|---|---|---|
| Diameter (exam) | 1x1 | 2x2 | 2x2 | 1x2 | | | | | | |
| Height (exam) | Raised | raised | raised | raised | raised | | | | | |
| 3D diameter | 2.200 | 2.450 | 2.060 | 1.912 | 2.115 | | | | | |
| 3D perp diameter | 1.886 | 2.289 | 1.939 | 1.505 | 1.870 | | | | | |
| 3D av height | 0.087 | 0.052 | 0.046 | 0.028 | 0.042 | | | | | |
| 3D Av Ht/Area * 10^4 | 66.450 | 29.370 | 36.720 | 30.800 | 33.590 | Background region at visit: | | | Average bkdg: | |
| 3D volume | 0.198 | 0.146 | 0.091 | 0.030 | 0.089 | V1 | V4 | V5 | MEAN | SD |
| 3D absol volume | 0.198 | 0.178 | 0.101 | 0.030 | 0.090 | 0.026 | 0.025 | 0.018 | **0.023** | 0.00 |

| | | | | |
|---|---|---|---|---|
| 88 | 13 | 6 | leg, pretibial | Too small, just a hair follicle |

| 013MB L6 | V1 | V2 | V3 | V4 | V5 | | | | | |
|---|---|---|---|---|---|---|---|---|---|---|
| Diameter (exam) | 1x1 | 1x1 | 1x1 | 1x1 | | V1 | V2 | V5 | | |
| Height (exam) | raised | raised | flat | flat | flat | | | | | |
| 3D diameter | 1.509 | 1.687 | 1.616 | 1.229 | 1.418 | 2.227 | 2.165 | 1.874 | | |
| 3D perp diameter | 0.927 | 1.333 | 0.982 | 0.980 | 1.255 | 1.86 | 1.786 | 1.607 | | |
| 3D av height | 0.018 | 0.024 | 0.033 | 0.030 | 0.004 | 0.007 | 0.002 | 0.001 | | |
| 3D volume | 0.008 | 0.018 | 0.020 | 0.012 | 0.001 | 0.017 | 0.003 | 0.002 | MEAN | SD |
| 3D absol volume | 0.009 | 0.018 | 0.020 | 0.012 | 0.006 | 0.026 | 0.025 | 0.018 | **0.023** | 0.00 |

| | | | | | |
|---|---|---|---|---|---|
| 89 | 13 | 7 | leg, pretibial | Biopsy at Visit 5: Biopsy: Scar | Scar/keloid (Bx-proven) |

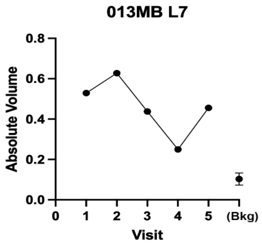

| 013MB L7 | V1 | V2 | V3 | V4 | V5 | | | | | |
|---|---|---|---|---|---|---|---|---|---|---|
| Diameter (exam) | 3x4 | 4x2 | 3x3 | 3x3 | | | | | | |
| Height (exam) | Raised | raised | raised | raised | raised | | | | | |
| 3D diameter | 3.986 | 4.022 | 3.626 | 3.590 | 4.031 | | | | | |
| 3D perp diameter | 2.640 | 2.957 | 2.550 | 2.338 | 3.057 | | | | | |
| 3D av height | 0.092 | 0.093 | 0.083 | 0.060 | 0.063 | | | | | |
| 3D Av Ht/Area * 10^4 | 26.550 | 24.280 | 27.660 | 21.600 | 16.040 | Background region at visit: | | | Average bkdg: | |
| 3D volume | 0.529 | 0.625 | 0.438 | 0.250 | 0.455 | V1 | V4 | V5 | MEAN | SD |
| 3D absol volume | 0.529 | 0.628 | 0.438 | 0.250 | 0.456 | 0.139 | 0.073 | 0.097 | **0.103** | 0.03 |

| | | | | | |
|---|---|---|---|---|---|
| 90 | 13 | 8 | thigh | Biopsy at Visit 5: Biopsy: Scar | Scar/keloid (Bx-proven) |

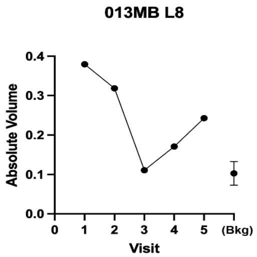

| 013MB L8 | V1 | V2 | V3 | V4 | V5 | | | | | |
|---|---|---|---|---|---|---|---|---|---|---|
| Diameter (exam) | 4x4 | 2x3 | 2x3 | 1x1 | | | | | | |
| Height (exam) | raised | flat | slightly r | slightly raised | | | | | | |
| 3D diameter | 3.8166 | 3.7049 | 3.1588 | 3.5212 | 3.3915 | | | | | |
| 3D perp diameter | 2.8908 | 2.6884 | 2.5624 | 2.3245 | 3.1721 | | | | | |
| 3D av height | 0.0547 | 0.0537 | 0.0237 | 0.037 | 0.0393 | | | | | |
| 3D Av Ht/Area * 10^4 | 15.47 | 16.74 | 9.2 | 13.77 | 11.61 | Background region at visit: | | | Average bkdg: | |
| 3D volume | 0.3794 | 0.3185 | 0.1034 | 0.1708 | 0.238 | V1 | V4 | V5 | MEAN | SD |
| 3D absol volume | 0.3794 | 0.3185 | 0.1109 | 0.1708 | 0.2428 | 0.139 | 0.073 | 0.097 | **0.103** | 0.03 |

| Lesion Index # | Subject # | Lesion # | Body Location | Diagnosis by Biopsy | Notes & Response to VitD |
|---|---|---|---|---|---|
| *91* | 13 | 9 | | Biopsy at Visit 5:<br>Biopsy: Scar | Lesion added at Visit 3<br>Missing visits 1 and 2: EXCLUDE<br>Scar/keloid |

| 013MB L9 | V1 | V2 | V3 | V4 | V5 |
|---|---|---|---|---|---|
| Diameter (exam) | | | 5x5 | 5x6 | |
| Height (exam) | | | slightly r | slightly raised | |
| 3D diameter | | | | | |
| 3D perp diameter | | | | | |
| 3D av height | | | | | |
| 3D Av Ht/Area * 10^4 | | | | | |
| 3D volume | | | 1.9168 | 1.6157 | 1.783 |
| 3D absol volume | | | 1.9176 | 1.6157 | 1.786 |

| *Lesion Index #* | Subject # | Lesion # | Body Location | Diagnosis by Biopsy | Graphical data | Notes & Response to VitD | 3-D lesion data | Visit 1 | Visit 2 | Visit 3 | Visit 4 | Visit 5 | Response outcome | VDR gene |
|---|---|---|---|---|---|---|---|---|---|---|---|---|---|---|

High-dose Vit D assignment: before V2

Lengths are in (mm); Volumes in (mm^3)

VDR alleles: not done

*92* | 14 | 1 | forearm

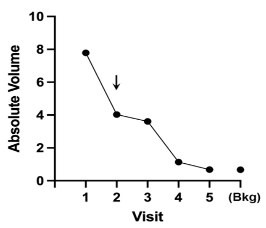

VitD=no

Gone at V5

| 014 BC L1 | V1 | V2 | V3 | V4 | V5 |
|---|---|---|---|---|---|
| Diameter (exam) | | | | | |
| Height (exam) | | | | | |
| 3D diameter | 7.911 | 6.932 | 8.385 | 6.756 | 6.284 |
| 3D perp diameter | 7.67 | 6.538 | 8.104 | 5.998 | 5.44 |
| 3D av height | 0.198 | 0.124 | 0.097 | 0.053 | 0.05 |
| 3D Av Ht/Area * 10^4 | 10.38 | 8.72 | 4.54 | 4.15 | 4.631 |
| 3D volume | 8.071 | 3.663 | 3.683 | 1.16 | 0.848 |
| 3D absol volume | 8.074 | 3.664 | 3.683 | 1.16 | 0.848 |

Response outcome: **V5**

Background region at visit: / **Average bkdg:**

| V1 | V4 | V5 | MEAN | SD |
|---|---|---|---|---|
| 0.733 | 0.382 | 0.728 | **0.614** | 0.20 |

*93* | 14 | 2 | arm, elbow

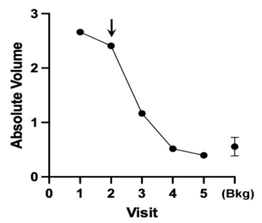

VitD=yes

Gone at V4

| 014 BC L2 | V1 | V2 | V3 | V4 | V5 |
|---|---|---|---|---|---|
| Diameter (exam) | | | | | |
| Height (exam) | | | | | |
| 3D diameter | 11.257 | 11.289 | 6.955 | 6.913 | 7.048 |
| 3D perp diameter | 9.293 | 7.682 | 5.124 | 5.197 | 5.281 |
| 3D av height | 0.036 | 0.044 | 0.001 | 0.001 | 0.005 |
| 3D Av Ht/Area * 10^4 | 1.085 | 1.557 | 0.087 | 0.086 | 0.4188 |
| 3D volume | 2.352 | 2.159 | 0.008 | 0.024 | 0.104 |
| 3D absol volume | 2.66 | 2.408 | 1.165 | 0.515 | 0.395 |

Response outcome: **V4**

Background region at visit: / **Average bkdg:**

| V1 | V4 | V5 | MEAN | SD |
|---|---|---|---|---|
| 0.367 | 0.703 | 0.599 | **0.556** | 0.17 |

*94* | 14 | 3 | back

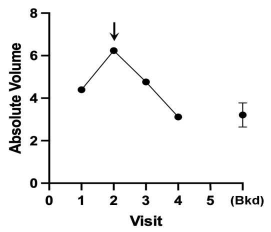

VitD = no

Gone at V4

| 014 BC L3 | V1 | V2 | V3 | V4 | V5 |
|---|---|---|---|---|---|
| Diameter (exam) | | | | | |
| Height (exam) | | | | | |
| 3D diameter | 14.664 | 13.679 | 13.47 | 13.442 | |
| 3D perp diameter | 10.923 | 10.923 | 11.048 | 10.628 | |
| 3D av height | 0.006 | 0.008 | 0.004 | 0.018 | |
| 3D Av Ht/Area * 10^4 | 0.23 | 0.155 | 0.186 | 3.178 | |
| 3D volume | 0.563 | 0.421 | 0.775 | 1.748 | photo |
| 3D absol volume | 4.396 | 6.235 | 4.765 | 3.116 | wrong site |

Response outcome: **V4**

Background region at visit: / **Average bkdg:**

| V1 | V4 | V4 | MEAN | SD |
|---|---|---|---|---|
| 3.761 | 2.629 | 3.241 | **3.210** | 0.57 |

*95* | **14** | **4** | back

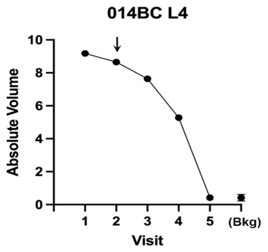

Gone at V5

VitD = no

| 014 BC L4 | V1 | V2 | V3 | V4 | V5 |
|---|---|---|---|---|---|
| Diameter (exam) | | | | | |
| Height (exam) | | | | | |
| 3D diameter | 8.591 | 8.456 | 8.46 | 9.041 | 8.937 |
| 3D perp diameter | 7.426 | 6.325 | 5.963 | 4.608 | 4.546 |
| 3D av height | 0.214 | 0.256 | 0.207 | 0.256 | 0.013 |
| 3D Av Ht/Area * 10^4 | 10.62 | 14.92 | 12.67 | 17.51 | 0.911 |
| 3D volume | 8.316 | 8.642 | 7.582 | 5.274 | 0.267 |
| 3D absol volume | 8.316 | 8.651 | 7.645 | 5.276 | 0.419 |

**V5**

Background region at visit: **Average bkdg:**

| V1 | V4 | V5 | MEAN | SD |
|---|---|---|---|---|
| 0.231 | 0.491 | 0.606 | **0.443** | 0.19 |

*96* | **14** | **5** | back

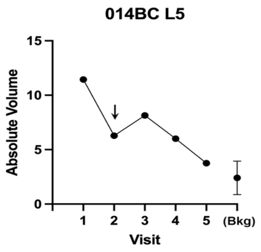

Gone at V5

VitD = no

| 014 BC L5 | V1 | V2 | V3 | V4 | V5 |
|---|---|---|---|---|---|
| Diameter (exam) | | | | | |
| Height (exam) | | | | | |
| 3D diameter | 12.93 | 12.349 | 12.995 | 11.378 | 11.405 |
| 3D perp diameter | 11.309 | 10.744 | 10.808 | 7.489 | 6.76 |
| 3D av height | 0.116 | 0.071 | 0.09 | 0.095 | 0.004 |
| 3D Av Ht/Area * 10^4 | 2.52 | 1.69 | 2.02 | 3.4 | 0.15 |
| 3D volume | 11.431 | 6.097 | 8.017 | 5.829 | 0.18 |
| 3D absol volume | 11.451 | 6.28 | 8.143 | 6.011 | 1.738 |

**V5**

Background region at visit: **Average bkdg:**

| V1 | V4 | V5 | MEAN | SD |
|---|---|---|---|---|
| 0.724 | 2.755 | 3.765 | **2.415** | 1.55 |

*97* | **14** | **6** | arm, axillary

Biopsy at Visit 5:

BCC, superficial, nodular, and micronodular

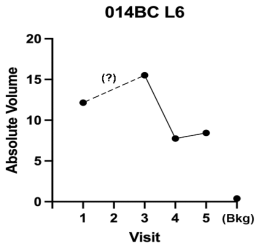

Did not resolve with PDT

VitD = no

| 014 BC L6 | V1 | V2 | V3 | V4 | V5 |
|---|---|---|---|---|---|
| Diameter (exam) | | | | | |
| Height (exam) | | | slightly r | slightly raised | |
| 3D diameter | 9.671 | n/d | 9.291 | 7.727 | 5.329 |
| 3D perp diameter | 6.913 | n/d | 6.129 | 4.47 | 3.857 |
| 3D av height | 0.254 | n/d | 0.448 | 0.328 | 0.608 |
| 3D Av Ht/Area * 10^4 | 11.76 | | 23.99 | 28.07 | 91.74 |
| 3D volume | 12.147 | n/d | 15.394 | 7.745 | 8.453 |
| 3D absol volume | 12.152 | n/d | 15.522 | 7.758 | 8.458 |

**NC**

Background region at visit: **Average bkdg:**

| V1 | V4 | V5 | MEAN | SD |
|---|---|---|---|---|
| 0.429 | 0.45 | 0.288 | **0.389** | 0.09 |

*98* | **14** | **7** | shoulder

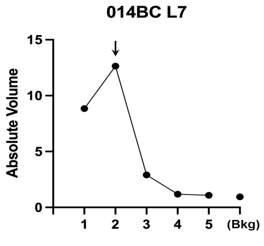

Gone by V5

VitD=yes

| 014 BC L7 | V1 | V2 | V3 | V4 | V5 |
|---|---|---|---|---|---|
| Diameter (exam) | | | | | |
| Height (exam) | | | slightly r | slightly raised | |
| 3D diameter | 14.723 | 14.663 | 11.365 | 7.283 | 7.129 |
| 3D perp diameter | 10.012 | 9.388 | 8.864 | 5.892 | 5.754 |
| 3D av height | 0.095 | 0.144 | 0.047 | 0.022 | 0.035 |
| 3D Av Ht/Area * 10^4 | 1.98 | 3.17 | 1.46 | 1.61 | 2.69 |
| 3D volume | 8.467 | 12.529 | 2.665 | 0.549 | 0.793 |
| 3D absol volume | 8.849 | 12.648 | 2.91 | 1.178 | 1.075 |

**V5**

Background region at visit: **Average bkdg:**

| V1 | V4 | V5 | MEAN | SD |
|---|---|---|---|---|
| 0.941 | 0.951 | 0.643 | **0.845** | 0.18 |

*99* | **14** | **8** | chest

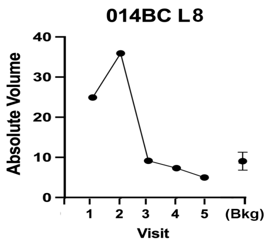

Gone by V3

| 014 BC L8 | V1 | V2 | V3 | V4 | V5 |
|---|---|---|---|---|---|
| Diameter (exam) | | | | | |
| Height (exam) | | | slightly r | slightly raised | |
| 3D diameter | 23.851 | 22.474 | 22.656 | 16.8 | 16.282 |
| 3D perp diameter | 13.972 | 15.821 | 13.124 | 11.304 | 10.95 |
| 3D av height | 0.027 | 0.134 | 0.011 | 0.004 | 0.011 |
| 3D Av Ht/Area * 10^4 | 0.24 | 1.163 | 0.109 | 0.066 | 0.189 |
| 3D volume | 6.98 | 34.911 | 2.238 | 0.498 | 1.288 |
| 3D absol volume | 24.727 | 35.987 | 9.169 | 7.308 | 4.975 |

**V3**

Background region at visit: **Average bkdg:**

| V1 | V4 | V5 | MEAN | SD |
|---|---|---|---|---|
| 10.973 | 9.616 | 6.599 | **9.063** | 2.24 |

| Lesion Index # | Subject # | Lesion # | Body Location | Diagnosis by Biopsy |
|---|---|---|---|---|
| *100* | **14** | **9** | chest | Biopsy at Visit 5: BCC, superficial, nodular, and trichoepithelial |

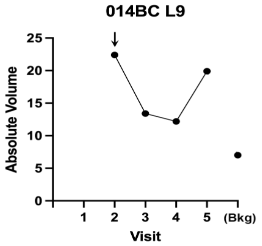

Shrank at first, then grew.
Did not resolve with PDT

**VitD=yes**

| **014 BC L9** | V1 | V2 | V3 | V4 | V5 |
|---|---|---|---|---|---|
| Diameter (exam) | | | | | |
| Height (exam) | | | slightly r | slightly raised | |
| 3D diameter | n/d | 14.392 | 11.354 | 9.711 | 10.996 |
| 3D perp diameter | n/d | 10.75 | 8.76 | 8.325 | 10.036 |
| 3D av height | n/d | 0.216 | 0.191 | 0.232 | 0.284 |
| 3D Av Ht/Area * 10^4 | | 4.35 | 6.01 | 9.08 | 8.17 |
| 3D volume | n/d | 22.367 | 13.406 | 12.224 | 19.87 |
| 3D absol volume | n/d | 22.369 | 13.422 | 12.234 | 19.881 |

**NC**

| Background region at visit: | | | **Average bkdg:** | |
|---|---|---|---|---|
| V1 | V4 | V5 | MEAN | SD |
| 8.259 | 5.147 | 8.584 | **7.330** | 1.90 |

| Lesion Index # | Subject # | Lesion # | Body Location |
|---|---|---|---|
| *101* | **14** | **10** | nose, ala |

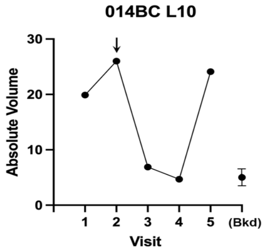

Shrank at first, then grew.
Did not resolve with PDT

**VitD=yes**

| **014 BC L10** | V1 | V2 | V3 | V4 | V5 |
|---|---|---|---|---|---|
| Diameter (exam) | | | | | |
| Height (exam) | | | slightly r | slightly raised | |
| 3D diameter | 12.899 | 12.547 | 9.177 | 8.064 | 12.286 |
| 3D perp diameter | 7.701 | 8.654 | 5.056 | 4.622 | 7.646 |
| 3D av height | 0.297 | 0.378 | 0.233 | 0.171 | 0.381 |
| 3D Av Ht/Area * 10^4 | 8.91 | 10.71 | 14.64 | 13.53 | 12.21 |
| 3D volume | 19.741 | 25.419 | 6.739 | 4.377 | 23.994 |
| 3D absol volume | 19.87 | 26.026 | 6.893 | 4.686 | 24.118 |

**NC**

| Background region at visit: | | | **Average bkdg:** | |
|---|---|---|---|---|
| V1 | V4 | V5 | MEAN | SD |
| 3.309 | 5.793 | 6.022 | **5.041** | 1.50 |

| *Lesion Index #* | Subject # | Lesion # | Body Location | Diagnosis by Biopsy | Graphical data | Notes & Response to VitD | 3-D lesion data | Visit 1 | Visit 2 | Visit 3 | Visit 4 | Visit 5 | Response outcome | VDR gene |
|---|---|---|---|---|---|---|---|---|---|---|---|---|---|---|

**High-dose Vit D assignment: before V3**

Lengths are in (mm); Volumes in (mm^3)

**VDR alleles:** (not done)

| Lesion Index # | Subject # | Lesion # | Body Location |
|---|---|---|---|
| *102* | **15** | **1** | scalp |

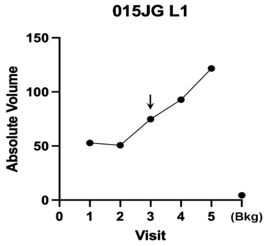

Big dome-shaped nodule,
Unresponsive to PDT

**Vit D= no**

| **015 JG L1** | V1 | V2 | V3 | V4 | V5 |
|---|---|---|---|---|---|
| Diameter (exam) | | | | | |
| Height (exam) | | | slightly r | slightly raised | |
| 3D diameter | 9.967 | 9.976 | 11.114 | 12.056 | 12.2 |
| 3D perp diameter | 8.485 | 9.003 | 10.764 | 10.77 | 11.139 |
| 3D av height | 0.928 | 0.848 | 0.985 | 1.052 | 1.286 |
| 3D Av Ht/Area * 10^4 | 34.71 | 29.98 | 26.2 | 25.7 | 30.06 |
| 3D volume | 52.774 | 50.471 | 74.314 | 92.616 | 121.52 |
| 3D absol volume | 52.803 | 50.639 | 74.795 | 92.754 | 121.68 |

**NC**

| Background region at visit: | | | **Average bkdg:** | |
|---|---|---|---|---|
| V1 | V4 | V5 | MEAN | SD |
| 2.206 | 5.775 | 5.37 | **4.450** | 1.95 |

| Lesion Index # | Subject # | Lesion # | Body Location |
|---|---|---|---|
| *103* | **15** | **2** | scalp |

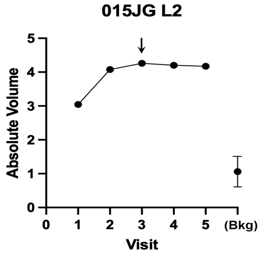

Shiny raised nodule
Unresponsive to PDT

**Vit D= no**

| **015 JG L2** | V1 | V2 | V3 | V4 | V5 |
|---|---|---|---|---|---|
| Diameter (exam) | | | | | |
| Height (exam) | | | slightly r | slightly raised | |
| 3D diameter | 7.272 | 5.924 | 5.987 | 5.14 | 5.293 |
| 3D perp diameter | 6.328 | 4.708 | 5.462 | 4.893 | 5.058 |
| 3D av height | 0.113 | 0.212 | 0.194 | 0.267 | 0.247 |
| 3D Av Ht/Area * 10^4 | 7.78 | 23.89 | 18.84 | 33.77 | 29.35 |
| 3D volume | 3.044 | 4.058 | 4.255 | 4.202 | 4.161 |
| 3D absol volume | 3.044 | 4.076 | 4.259 | 4.202 | 4.174 |

**NC**

| Background region at visit: | | | **Average bkdg:** | |
|---|---|---|---|---|
| V1 | V4 | V5 | MEAN | SD |
| 0.715 | 0.895 | 1.57 | **1.060** | 0.45 |

104 | 15 | 3 | temple | Biopsy at Visit 5: BCC, superficial, nodular, and trichoepitheliom.

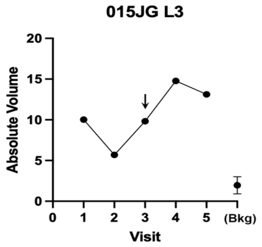

Vit D= no

Raised nodule, continued to grow
Unresponsive to PDT

| 015 JG L3 | V1 | V2 | V3 | V4 | V5 |
|---|---|---|---|---|---|
| Diameter (exam) | | | | | |
| Height (exam) | | | slightly r | slightly raised | |
| 3D diameter | 10.657 | 10.507 | 12.624 | 14.06 | 13.41 |
| 3D perp diameter | 9.429 | 7.219 | 9.245 | 9.081 | 6.851 |
| 3D av height | 0.178 | 0.093 | 0.146 | 0.212 | 0.223 |
| 3D Av Ht/Area * 10^4 | 5.61 | 3.76 | 3.88 | 5.04 | 6.91 |
| 3D volume | 9.78 | 4.312 | 9.113 | 14.772 | 13.102 |
| 3D absol volume | 10.032 | 5.69 | 9.833 | 14.783 | 13.132 |

NC

| Background region at visit: | | | Average bkdg: | |
|---|---|---|---|---|
| V1 | V4 | V5 | MEAN | SD |
| 3.164 | 1.393 | 1.302 | **1.953** | 1.05 |

105 | 15 | 4 | nose, ala

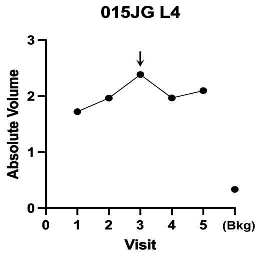

Vit D = yes

Big dome-shaped nodule,
minimal response to PDT

| 015 JG L4 | V1 | V2 | V3 | V4 | V5 |
|---|---|---|---|---|---|
| Diameter (exam) | | | | | |
| Height (exam) | | | | | |
| 3D diameter | 3.998 | 3.767 | 4.108 | 3.672 | 3.606 |
| 3D perp diameter | 3.121 | 3.228 | 3.789 | 3.585 | 3.477 |
| 3D av height | 0.203 | 0.242 | 0.258 | 0.232 | 0.23 |
| 3D Av Ht/Area * 10^4 | | | | | |
| 3D volume | 1.704 | 1.963 | 2.379 | 1.967 | 2.017 |
| 3D absol volume | 1.722 | 1.963 | 2.384 | 1.967 | 2.096 |

NC

| Background region at visit: | | | Average bkdg: | |
|---|---|---|---|---|
| V1 | V4 | V5 | MEAN | SD |
| 0.328 | 0.373 | 0.298 | **0.333** | 0.04 |

106 | 15 | 5 | nose, ala

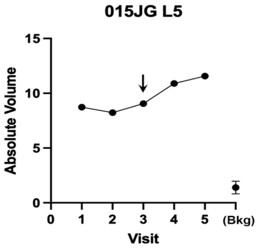

Vit D= no

Dome-shaped nodule,
just kept growing

| 015 JG L5 | V1 | V2 | V3 | V4 | V5 |
|---|---|---|---|---|---|
| Diameter (exam) | | | | | |
| Height (exam) | | | slightly r | slightly raised | |
| 3D diameter | 5.369 | 5.349 | 5.485 | 7.127 | 6.6 |
| 3D perp diameter | 5.149 | 5.247 | 5.105 | 6.06 | 5.975 |
| 3D av height | 0.426 | 0.408 | 0.377 | 0.309 | 0.395 |
| 3D Av Ht/Area * 10^4 | 50.99 | 46.277 | 42.801 | 22.624 | 31.805 |
| 3D volume | 8.712 | 8.176 | 8.11 | 9.726 | 11.385 |
| 3D absol volume | 8.734 | 8.227 | 9.062 | 10.896 | 11.567 |

NC

| Background region at visit: | | | Average bkdg: | |
|---|---|---|---|---|
| V1 | V4 | V5 | MEAN | SD |
| 1.181 | 0.957 | 2.058 | **1.399** | 0.58 |

107 | 15 | 6 | nose, side

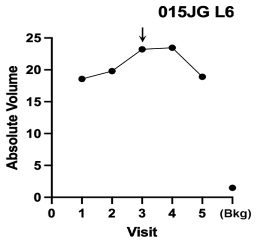

Vit D= no

Dome-shaped nodule,
Unresponsive to PDT

| 015 JG L6 | V1 | V2 | V3 | V4 | V5 |
|---|---|---|---|---|---|
| Diameter (exam) | | | | | |
| Height (exam) | | | slightly r | slightly raised | |
| 3D diameter | 6.48 | 7.311 | 7.634 | 7.793 | 7.35 |
| 3D perp diameter | 6.326 | 6.741 | 6.712 | 6.652 | 6.389 |
| 3D av height | 0.661 | 0.568 | 0.647 | 0.597 | 0.546 |
| 3D Av Ht/Area * 10^4 | 51.32 | 36.62 | 40.03 | 36.43 | 36.83 |
| 3D volume | 18.582 | 19.798 | 23.199 | 23.346 | 18.87 |
| 3D absol volume | 18.582 | 19.798 | 23.228 | 23.464 | 18.912 |

NC

| Background region at visit: | | | Average bkdg: | |
|---|---|---|---|---|
| V1 | V4 | V5 | MEAN | SD |
| 1.988 | 1.246 | 1.235 | **1.490** | 0.43 |

108 | 15 | 7 | ear

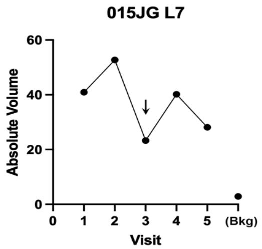

Vit D= no

Large nodule on ear (ulcerated),
partially responsive to PDT

| 015 JG L7 | V1 | V2 | V3 | V4 | V5 |
|---|---|---|---|---|---|
| Diameter (exam) | | | | | |
| Height (exam) | | | slightly r | slightly raised | |
| 3D diameter | 12.026 | 11.952 | 11.443 | 10.897 | 12.386 |
| 3D perp diameter | 6.706 | 7.295 | 6.965 | 7.468 | 7.29 |
| 3D av height | 0.694 | 0.85 | 0.465 | 0.728 | 0.527 |
| 3D Av Ht/Area * 10^4 | 25.18 | 29.21 | 17.47 | 27.48 | 17.33 |
| 3D volume | 40.852 | 52.705 | 23.225 | 40.199 | 27.914 |
| 3D absol volume | 40.915 | 52.705 | 23.283 | 40.199 | 28.108 |

NC

| Background region at visit: | | | Average bkdg: | |
|---|---|---|---|---|
| V1 | V4 | V5 | MEAN | SD |
| 3.169 | 2.027 | 3.444 | **2.880** | 0.75 |

| Lesion Index # | Subject # | Lesion # | Body Location | Diagnosis by Biopsy |
|---|---|---|---|---|
| *109* | 15 | 8 | back | |

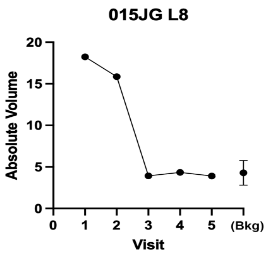

Large flat superficial lesion, gone at V3

| 015 JG L8 | V1 | V2 | V3 | V4 | V5 |
|---|---|---|---|---|---|
| Diameter (exam) | | | | | |
| Height (exam) | | | slightly r | slightly raised | |
| 3D diameter | 16.26 | 16.564 | 11.262 | 11.101 | 11.253 |
| 3D perp diameter | 13.004 | 14.843 | 9.6 | 9.814 | 9.863 |
| 3D av height | 0.11 | 0.088 | 0.029 | 0.055 | 0.049 |
| 3D Av Ht/Area * 10^4 | 1.63 | 1.14 | 0.848 | 1.601 | 1.399 |
| 3D volume | 17.824 | 15.588 | 2.219 | 4.138 | 3.71 |
| 3D absol volume | 18.234 | 15.855 | 3.925 | 4.336 | 3.914 |

**V3**

| Background region at visit: | | | Average bkdg: | |
|---|---|---|---|---|
| V1 | V4 | V5 | MEAN | SD |
| 5.303 | 4.975 | 2.587 | **4.288** | 1.48 |

| Lesion Index # | Subject # | Lesion # | Body Location | Diagnosis by Biopsy |
|---|---|---|---|---|
| *110* | 15 | 9 | shoulder | Biopsy at Visit 5: BCC, superficial, nodular, and micronodular |

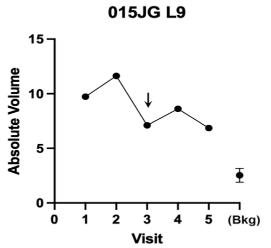

Vit D= no

Dome-shaped nodule, partially responsive to PDT

| 015 JG L9 | V1 | V2 | V3 | V4 | V5 |
|---|---|---|---|---|---|
| Diameter (exam) | | | | | |
| Height (exam) | | | slightly r | slightly raised | |
| 3D diameter | 9.147 | 9.006 | 8.882 | 9.113 | 9.183 |
| 3D perp diameter | 7.344 | 7.198 | 6.974 | 7.233 | 7.118 |
| 3D av height | 0.2 | 0.248 | 0.147 | 0.164 | 0.134 |
| 3D Av Ht/Area * 10^4 | 9.366 | 12.026 | 7.441 | 7.815 | 6.421 |
| 3D volume | 9.723 | 11.616 | 7.063 | 8.601 | 6.809 |
| 3D absol volume | 9.731 | 11.636 | 7.106 | 8.616 | 6.861 |

**NC**

| Background region at visit: | | | Average bkdg: | |
|---|---|---|---|---|
| V1 | V4 | V5 | MEAN | SD |
| 3.243 | 2.002 | 2.334 | **2.526** | 0.64 |

| Lesion Index # | Subject # | Lesion # | Body Location | Diagnosis by Biopsy |
|---|---|---|---|---|
| *111* | 15 | 10 | back | Biopsy at Visit 5: BCC, nodular & trichoepitheliom. |

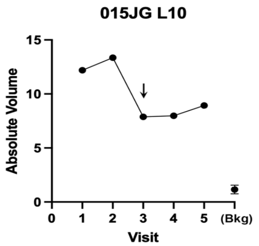

Vit D= no

Fleshy nodule, minimally responsive to PDT

| 015 JG L10 | V1 | V2 | V3 | V4 | V5 |
|---|---|---|---|---|---|
| Diameter (exam) | | | | | |
| Height (exam) | | | slightly r | slightly raised | |
| 3D diameter | 8.957 | 9.079 | 8.925 | 8.586 | 9.329 |
| 3D perp diameter | 7.982 | 6.071 | 5.243 | 5.829 | 6.132 |
| 3D av height | 0.283 | 0.35 | 0.239 | 0.228 | 0.243 |
| 3D Max Ht/Area*10^3 | 12.558 | 19.416 | 15.15 | 13.971 | 12.943 |
| 3D volume | 12.116 | 13.309 | 7.867 | 7.94 | 8.935 |
| 3D absol volume | 12.194 | 13.361 | 7.876 | 7.986 | 8.937 |

**NC**

| Background region at visit: | | | Average bkdg: | |
|---|---|---|---|---|
| V1 | V4 | V5 | MEAN | SD |
| 1.422 | 1.341 | 0.694 | **1.152** | 0.40 |

### ARIZONA PATIENTS:

| *Lesion Index #* | Subject # | Lesion # | Body Location | Diagnosis by Biopsy | Graphical data | Notes & Response to VitD | 3-D lesion data | Visit 1 | Visit 2 | Visit 3 | Visit 4 | Visit 5 | Response outcome | VDR gene |
|---|---|---|---|---|---|---|---|---|---|---|---|---|---|---|

**High-dose Vit D assignment: before V2**

| Lesion Index # | Subject # | Lesion # | Body Location | Diagnosis by Biopsy | Notes & Response to VitD |
|---|---|---|---|---|---|
| *112* | 26 | 1 | cheek | N/A | Visible papule at V1 & V2, gone by V3. |

| 026 DB 1 | V1 | V2 | V3 | V4 | V5 |
|---|---|---|---|---|---|
| 3D diameter | Problem with 3D reconstruction- | | | | |
| 3D perp diameter | Unable to process due to Hair causing artifacts. | | | | |

**VDR alleles:**
(not done)

| Lesion Index # | Subject # | Lesion # | Body Location | Diagnosis by Biopsy | Notes & Response to VitD |
|---|---|---|---|---|---|
| *113* | 26 | 2 | cheek | BCC, nodular | Tumor already gone at V2<br>Nothing present to measure.<br>DO NOT USE |

| 026 DB 2 | V1 | V2 | V3 | V4 | V5 |
|---|---|---|---|---|---|
| 3D diameter | | | | | |
| 3D perp diameter | | | | | |

| Lesion Index # | Subject # | Lesion # | Body Location | Diagnosis by Biopsy | Notes & Response to VitD |
|---|---|---|---|---|---|
| *114* | 26 | 3 | nose, dorsal | N/A | Tumor already gone at V2<br>DO NOT USE |

| 026 DB 3 | V1 | V2 | V3 | V4 | V5 |
|---|---|---|---|---|---|
| 3D diameter | | | | | |
| 3D perp diameter | | | | | |

| | | | | | | | | | | | | | | | |
|---|---|---|---|---|---|---|---|---|---|---|---|---|---|---|---|
| *115* | **26** | **4** | postauricul | BCC, superficial | | | **026 DB 4** | V1 | V2 | V3 | V4 | V5 | | | |
| | | | | | | Tumor already gone at V2 | 3D diameter | | | | | | | | |
| | | | | | | DO NOT USE | 3D perp diameter | | | | | | | | |
| *116* | **26** | **5** | leg, pretibia | BCC, superficial | | | **026 DB 5** | V1 | V2 | V3 | V4 | V5 | | | |
| | | | | | | Tumor already gone at V2 | 3D diameter | | | | | | | | |
| | | | | | | DO NOT USE | 3D perp diameter | | | | | | | | |
| *117* | **26** | **6** | leg, pretibia | N/A | | | **026 DB 6** | V1 | V2 | V3 | V4 | V5 | | | |
| | | | | | INCOMPLETE DATA, CANNOT USE | | 3D diameter | Problem with 3D reconstruction- | | | | | | | |
| | | | | | | | 3D perp diameter | unable to process due to Hair causing artificacts. | | | | | | | |
| | | | | | | Raised plaque, visually gone by V4 | 3D av height | However, clearly gone by Visit 4 | | | | | | | |
| *118* | **26** | **7** | leg, calf | N/A | | | **026 DB 7** | V1 | V2 | V3 | V4 | V5 | | | |
| | | | | | INCOMPLETE DATA, CANNOT USE | | 3D diameter | 9.035 | 8.378 | 9.035 | 0 | 0 | | | |
| | | | | | | | 3D perp diameter | 6.526 | 6.713 | 6.526 | 0 | 0 | | | |
| | | | | | | | 3D av height | 0.08 | 0.083 | 0.08 | 0 | 0 | Background | | |
| | | | | | | | 3D volume | 3.013 | 3.117 | 3.013 | bad 3D reconstruction | | V1 | V2 | V5 |
| | | | | | | Small papule; visually gone by V4 | 3D absol volume | 3.218 | 3.164 | 3.218 | | | 0.429 | 0.656 | 0.429 |
| *119* | **26** | **8** | leg, pretibia | BCC, nodular | | | **026 DB 8** | V1 | V2 | V3 | V4 | V5 | | | |
| | | | | | | | 3D diameter | 9.096 | | | | | | | |
| | | | | | INCOMPLETE DATA, CANNOT USE | | 3D perp diameter | 6.518 | | | | | | | |
| | | | | | | | 3D av height | 0.038 | | | | | | | |
| | | | | | | | 3D volume | 1.394 | | bad 3D reconstruction | | | V1 | V2 | V5 |
| | | | | | | Small papule, visually gone by V3 | 3D absol volume | 1.478 | | | | | 0.835 | | |
| *120* | **26** | **9** | leg, pretibia | BCC, superficial and nodular | | Tumor already gone at V2 | **026 DB 9** | V1 | V2 | V3 | V4 | V5 | | | |
| | | | | | | DO NOT USE | 3D diameter | | | | | | | | |
| | | | | | | | 3D perp diameter | | | | | | | | |

| *Lesion Index #* | Subject # | Lesion # | Body Location | Diagnosis by Biopsy | Graphical data | Notes & Response to VitD | 3-D lesion data | Visit 1 | Visit 2 | Visit 3 | Visit 4 | Visit 5 | | | | Response outcome | VDR gene |
|---|---|---|---|---|---|---|---|---|---|---|---|---|---|---|---|---|---|
| | | | | | | **High-dose Vit D assignment: before V3** | | | | | | | | | | | **VDR alleles:** |
| *121* | **27** | **1** | postauricul | BCC, nodular | | | **027 DK L1** | V1 | V2 | V3 | V4 | V5 | | | | | (not done) |
| | | | | | | Tumor already gone at V2 | 3D diameter | | | | | | | | | | |
| *122* | **27** | **2** | temple | BCC, nodular | | | **027 DK L2** | V1 | V2 | V3 | V4 | V5 | | | | | |
| | | | | | INCOMPLETE DATA, CANNOT USE | | 3D diameter | 8.275 | 0 | 0 | 0 | 0 | | | | | |
| | | | | | | | 3D perp diameter | 7.066 | 0 | 0 | 0 | 0 | | | | | |
| | | | | | | | 3D av height | 0.064 | | | | | Background | | | | |
| | | | | | | | 3D volume | 2.377 | bad 3D reconstruction | | | | V1 | V2 | V5 | | |
| | | | | | | Small lesion, visually gone by V3 | 3D absol volume | 2.516 | | | | | 1.813 | | | | |
| *123* | **27** | **3** | neck | BCC, nodular | | | **027 DK L3** | V1 | V2 | V3 | V4 | V5 | | | | | |
| | | | | | | Tumor already gone at V2 | 3D diameter | | | | | | | | | | |
| | | | | | | Nothing present to measure. | 3D perp diameter | | | | | | | | | | |

| *Lesion Index #* | Subject # | Lesion # | Body Location | Diagnosis by Biopsy | Graphical data | Notes & Response to VitD | 3-D lesion data | Visit 1 | Visit 2 | Visit 3 | Visit 4 | Visit 5 | | | Response outcome | | VDR gene |
|---|---|---|---|---|---|---|---|---|---|---|---|---|---|---|---|---|---|
| | | | | | | **High-dose Vit D assignment: before V2** | | | | | | | | | | | **VDR alleles:** |
| *124* | **28** | **1** | shoulder | BCC, superficial and nodular | | | 028 LM 1 | V1 | V2 | V3 | V4 | V5 | | | | | (not done) |
| | | | | | | Tumor already gone at V2 | 3D diameter | | | | | | | | | | |
| | | | | | | Nothing present to measure. | 3D perp diameter | | | | | | | | | | |
| *125* | **28** | **2** | back | BCC, Superficial | | | 028 LM 2 | V1 | V2 | V3 | V4 | V5 | | | | | |
| | | | | | | | 3D diameter | 9.27 | 9.338 | 9.37 | 9.164 | 9.272 | | | | | |
| | | | | | | | 3D perp diameter | 7.803 | 6.998 | 7.837 | 7.736 | 6.784 | | | | | |
| | | | | | | | 3D av height | 0.062 | 0.032 | 0.002 | 0.015 | 0.018 | | | | | |
| | | | | | | Very small lesion, unhealed Bx | 3D Av Ht/Area * 10^4 | | | | | | Background region at visit: | | | **Average bkdg:** | |
| | | | | | | site replaced by scar at V3; | 3D volume | 2.975 | 1.538 | 0.103 | 0.704 | 0.826 | V1 | V4 | V5 | MEAN | SD |
| | | | | | | | 3D absol volume | 3.442 | 2.09 | 3.335 | 4.039 | 3.296 | 3.937 | 2.816 | 2.065 | **2.939** | 0.94 |
| *126* | **28** | **3** | back | BCC, Superficial | | | **028 LM 3** | V1 | V2 | V3 | V4 | V5 | | | | | |
| | | | | | | **Vit D= no** | 3D diameter | 15.208 | 13.259 | 12.998 | 36.48 | 11.706 | | | **V5** | | |
| | | | | | | | 3D perp diameter | 10.527 | 10.408 | 10.578 | 93.198 | 8.33 | | | | | |
| | | | | | | | 3D av height | 0.015 | 0 | 0.002 | 3.397 | 0.033 | | | | | |
| | | | | | | | 3D Av Ht/Area * 10^4 | | | | | | Background region at visit: | | | **Average bkdg:** | |
| | | | | | | Very small lesion, within Bx site | 3D volume | 1.813 | 0.03 | 0.188 | -0.011 | 2.081 | V1 | V4 | V5 | MEAN | SD |
| | | | | | | Skin appears normal by V5 | 3D absol volume | 12.887 | 16.599 | 20.789 | 13.139 | 3.263 | 8.599 | 5.003 | 7.29 | **6.964** | 1.82 |
| *127* | **28** | **4** | back | N/A | | | 028 LM 4 | V1 | V2 | V3 | V4 | V5 | | | | | |
| | | | | | | White flat scar with small pink mac | 3D diameter | | | | | | | | | | |
| | | | | | | Visually gone by V3 - EXCLUDE | 3D perp diameter | | | | | | | | | | |
| *128* | **28** | **5** | back | N/A | | | 028 LM 5 | V1 | V2 | V3 | V4 | V5 | | | | | |
| | | | | | | Flat red biopsy site at V1 and V2; nc | 3D diameter | | | | | | | | | | |
| | | | | | | EXCLUDE THIS LESION | 3D perp diameter | | | | | | | | | | |
| *129* | **28** | **6** | shoulder | N/A | | | 028 LM 6 | V1 | V2 | V3 | V4 | V5 | | | | | |
| | | | | | | Flat white biopsy scar at V1 and V2; | 3D diameter | | | | | | | | | | |
| | | | | | | EXCLUDE THIS LESION | 3D perp diameter | | | | | | | | | | |
| *130* | **28** | **7** | shoulder | N/A | | | 028 LM 7 | V1 | V2 | V3 | V4 | V5 | | | | | |
| | | | | | | Tumor already gone at V2 | 3D diameter | | | | | | | | | | |
| | | | | | | Nothing present to measure. | 3D perp diameter | | | | | | | | | | |
| *131* | **28** | **8** | sternum | N/A | | | 028 LM 8 | V1 | V2 | V3 | V4 | V5 | | | | | |
| | | | | | | Tumor already gone at V2 | 3D diameter | | | | | | | | | | |
| | | | | | | Nothing present to measure. | 3D perp diameter | | | | | | | | | | |

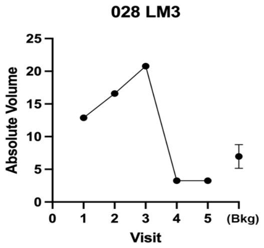

| *Lesion Index #* | Subject # | Lesion # | Body Location | Diagnosis by Biopsy | Graphical data | Notes & Response to VitD | 3-D lesion data | Visit 1 | Visit 2 | Visit 3 | Visit 4 | Visit 5 | | Response outcome | | VDR gene |
|---|---|---|---|---|---|---|---|---|---|---|---|---|---|---|---|---|
| | | | | | | **High-dose Vit D assignment: before V3** | | | | | | | | | | **VDR alleles:** |
| *132* | **29** | **1** | shoulder | BCC, superficial | | | **029 JH L1** | V1 | V2 | V3 | V4 | V5 | | | | (not done) |
| | | | | | | Tumor already gone at V2 | 3D diameter | | | | | | | | | |
| | | | | | | Nothing present to measure. | 3D perp diameter | | | | | | | | | |
| *133* | **29** | **2** | forehead | BCC, superficial and nodular | | | **029 JH L2** | V1 | V2 | V3 | V4 | V5 | | | | |
| | | | | | | Tumor already gone at V2 | 3D diameter | | | | | | | | | |
| | | | | | | Nothing present to measure. | 3D perp diameter | | | | | | | | | |
| *134* | **29** | **3** | forehead | BCC, nodular | | | **029 JH L3** | V1 | V2 | V3 | V4 | V5 | | | | |
| | | | | | | Tumor already gone at V2 | 3D diameter | | | | | | | | | |
| | | | | | | Nothing present to measure. | 3D perp diameter | | | | | | | | | |

| *Lesion Index #* | Subject # | Lesion # | Body Location | Diagnosis by Biopsy | Graphical data | Notes & Response to VitD | 3-D lesion data | Visit 1 | Visit 2 | Visit 3 | Visit 4 | Visit 5 | | Response outcome | | VDR gene |
|---|---|---|---|---|---|---|---|---|---|---|---|---|---|---|---|---|

**30- Patient Withdrew** PATIENT 030 RE-ENROLLED AND BECAME PATIENT 26

Lengths are in (mm); Volumes in (mm^3)

| *Lesion Index #* | Subject # | Lesion # | Body Location | Diagnosis by Biopsy | Graphical data | Notes & Response to VitD | 3-D lesion data | Visit 1 | Visit 2 | Visit 3 | Visit 4 | Visit 5 | | Response outcome | | VDR gene |
|---|---|---|---|---|---|---|---|---|---|---|---|---|---|---|---|---|
| | | | | | | **High-dose Vit D assignment: before V3** | | | | | | | | | | **VDR alleles:** |
| *135* | **31** | **1** | ankle | BCC, nodular | | | **031 RW L1** | V1 | V2 | V3 | V4 | V5 | | | | (not done) |
| | | | | | | Tumor already gone at V2 | 3D diameter | | | | | | | | | |
| | | | | | | Nothing present to measure. | 3D perp diameter | | | | | | | | | |
| *136* | **31** | **2** | leg, calf | BCC, nodular | | | **031 RW L2** | V1 | V2 | V3 | V4 | V5 | | | | |
| | | | | | | Tumor already gone at V2 | 3D diameter | | | | | | | | | |
| | | | | | | Nothing present to measure. | 3D perp diameter | | | | | | | | | |
| *137* | **31** | **3** | leg, calf | N/A | 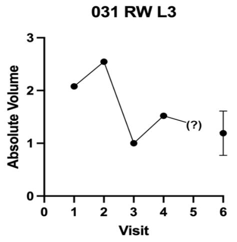 | | **031 RW L3** | V1 | V2 | V3 | V4 | V5 | | **V3** | | |
| | | | | | | | 3D diameter | 9.361 | 9.353 | 8.532 | 8.229 | Corrupted | | | | |
| | | | | | | | 3D perp diameter | 6.497 | 6.885 | 6.104 | 6.241 | image | | | | |
| | | | | | | | 3D av height | 0.051 | 0.059 | 0.031 | 0.047 | 0 | | | | |
| | | | | | | | 3D Av Ht/Area * 10^4 | | | | | | | | | |
| | | | | | | | 3D volume | 2.076 | 2.545 | 0.95 | 1.449 | 0 | | | | |
| | | | | | | Small lesion, visually gone at V3 | 3D absol volume | 2.078 | 2.545 | 1.002 | 1.52 | 0 | | | | |
| *138* | **31** | **4** | leg, calf | N/A | | | **031 RW L4** | V1 | V2 | V3 | V4 | V5 | | | | |
| | | | | | | Tumor already gone at V2 | 3D diameter | | | | | | | | | |
| | | | | | | Nothing present to measure. | 3D perp diameter | | | | | | | | | |

| Background | | | **Average bkdg:** | |
|---|---|---|---|---|
| V1 | V2 | V5 | MEAN | SD |
| 1.599 | 1.204 | 0.769 | **1.191** | 0.42 |

| *Lesion Index #* | Subject # | Lesion # | Body Location | Diagnosis by Biopsy | Graphical data | Notes & Response to VitD | 3-D lesion data | Visit 1 | Visit 2 | Visit 3 | Visit 4 | Visit 5 | | Response outcome | | VDR gene |
|---|---|---|---|---|---|---|---|---|---|---|---|---|---|---|---|---|

**32- Lost to Follow Up**

| *Lesion Index #* | Subject # | Lesion # | Body Location | Diagnosis by Biopsy | Graphical data | Notes & Response to VitD | 3-D lesion data | Visit 1 | Visit 2 | Visit 3 | Visit 4 | Visit 5 | | Response outcome | | VDR gene |
|---|---|---|---|---|---|---|---|---|---|---|---|---|---|---|---|---|
| | | | | | | **High-dose Vit D assignment: before V2** | | | | | | | | | | **VDR alleles:** |
| | | | | | | | | | | | | | | | | (not done) |
| *139* | **33** | **1** | hand | BCC, superficial | | This was small post-biopsy scar -- | **033 BS L1** | V1 | V2 | V3 | V4 | V5 | | | | |
| | | | | | N/A | visible at V1, gone by V2 | Diameter (exam) | | | | | | | | | |
| | | | | | | EXLUDE THIS LESION | Height (exam) | | | | | | | | | |
| *140* | **33** | **2** | ear, helix | BCC, nodular | | This was small post-biopsy scar -- | **033 BS L2** | V1 | V2 | V3 | V4 | V5 | | | | |
| | | | | | N/A | visible at V1, gone by V2 | Diameter (exam) | | | | | | | | | |
| | | | | | | EXLUDE THIS LESION | Height (exam) | | | | | | | | | |
| *141* | **33** | **3** | back | BCC, nodular | | This was small post-biopsy scar -- | **033 BS L3** | V1 | V2 | V3 | V4 | V5 | | | | |
| | | | | | N/A | visible at V1, gone by V2 | Diameter (exam) | | | | | | | | | |
| | | | | | | EXLUDE THIS LESION | Height (exam) | | | | | | | | | |
| *142* | **33** | **4** | neck | BCC, nodular | | This was small post-biopsy scar -- | **033 BS L4** | V1 | V2 | V3 | V4 | V5 | | | | |
| | | | | | N/A | visible at V1, gone by V2 | Diameter (exam) | | | | | | | | | |
| | | | | | | EXLUDE THIS LESION | Height (exam) | | | | | | | | | |

| *Lesion Index #* | Subject # | Lesion # | Body Location | Diagnosis by Biopsy | Graphical data | Notes & Response to VitD | 3-D lesion data | Visit 1 | Visit 2 | Visit 3 | Visit 4 | Visit 5 | | Response outcome | | VDR gene |
|---|---|---|---|---|---|---|---|---|---|---|---|---|---|---|---|---|
| | | | | | | **High-dose Vit D assignment: before V3** | Lengths are in (mm); Volumes in (mm^3) | | | | | | | | | **VDR alleles for patient 34** |
| | | | | | 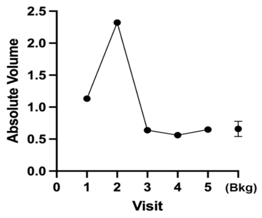 | | **034 MV L1** | V1 | V2 | V3 | V4 | V5 | | | | Ff LL |
| *143* | **34** | **1** | neck | BCC, superficial | | | 3D diameter | 7.426 | 8.842 | 6.829 | 7.393 | 7.218 | | **V3** | | |
| | | | | | | Small flat lesion, gone at V3 | 3D perp diameter | 5.409 | 6.832 | 4.181 | 4.576 | 4.590 | | | | |
| | | | | | | | 3D av height | 0.024 | 0.029 | 0.004 | 0.005 | 0.023 | | | | |
| | | | | | | | 3D Av Ht/Area * 10^4 | 1.860 | 1.500 | | | | | | | |
| | | | | | | | 3D volume | 0.774 | 1.218 | 0.072 | 0.108 | 0.54 | | | | |
| | | | | | | | 3D absol volume | 1.132 | 2.321 | 0.638 | 0.561 | 0.649 | | | | |
| *144* | **34** | **2** | back, upper | BCC, nodular | | | **034MV L2** | V1 | V2 | V3 | V4 | V5 | | | | Ff LL |
| | | | | | | Lesion not visible at V2; | Diameter (exam) | | | | | | | | | |
| | | | | | | EXCLUDE | Height (exam) | | | | | | | | | |
| *145* | **34** | **3** | forearm | BCC, nodular | | | **034MV L3** | V1 | V2 | V3 | V4 | V5 | | | | Ff LL |
| | | | | | | Lesion not visible at V2; | Diameter (exam) | | | | | | | | | |
| | | | | | | EXCLUDE | Height (exam) | | | | | | | | | |

| Background | | | **Average bkdg:** | |
|---|---|---|---|---|
| V1 | V2 | V5 | MEAN | SD |
| 0.647 | 0.54 | 0.789 | **0.659** | 0.12 |

| *Lesion Index #* | Subject # | Lesion # | Body Location | Diagnosis by Biopsy | Graphical data | Notes & Response to VitD | 3-D lesion data | Visit 1 | Visit 2 | Visit 3 | Visit 4 | Visit 5 | Response outcome | VDR gene |
|---|---|---|---|---|---|---|---|---|---|---|---|---|---|---|

**High-dose Vit D assignment: before V3**

Lengths are in (mm); Volumes in (mm^3)

**VDR alleles for patient 35**

| ff | LL |
|---|---|

| *Lesion Index #* | Subject # | Lesion # | Body Location | Diagnosis by Biopsy |
|---|---|---|---|---|
| *146* | **35** | **1** | Shoulder | BCC, superficial |

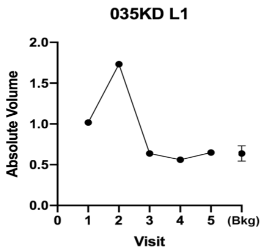

Small flat lesion, gone at V3

| **035KD L1** | V1 | V2 | V3 | V4 | V5 |
|---|---|---|---|---|---|
| 3D diameter | 7.426 | 8.842 | 6.829 | 7.393 | 7.218 |
| 3D perp diameter | 5.409 | 6.832 | 4.181 | 4.576 | 4.59 |
| 3D av height | 0.024 | 0.029 | 0.004 | 0.005 | 0.023 |
| 3D Av Ht/Area * 10^4 | 1.850 | 1.500 | | | |
| 3D volume | 0.774 | 1.218 | 0.072 | 0.108 | 0.54 |
| 3D absol volume | 1.132 | 2.321 | 0.638 | 0.561 | 0.649 |

Response outcome: **V3**

| Background region at visit | | | **Average bkg controls:** | |
|---|---|---|---|---|
| V1 | V2 | V5 | MEAN | SD |
| 0.647 | 0.540 | 0.726 | **0.638** | 0.093 |

| *Lesion Index #* | Subject # | Lesion # | Body Location | Diagnosis by Biopsy |
|---|---|---|---|---|
| *147* | **35** | **2** | Back | BCC, Superficial |

Looks like normal skin even at V2, DO NOT USE

| 035KD L2 | V1 | V2 | V3 | V4 | V5 |
|---|---|---|---|---|---|
| 3D volume | | -0.0053 | -0.07 | -0.1196 | |
| 3D absol volume | | 3.4792 | 0.4523 | 0.1601 | |

| *Lesion Index #* | Subject # | Lesion # | Body Location | Diagnosis by Biopsy |
|---|---|---|---|---|
| *148* | **35** | **3** | Back | BCC, Superficial |

DO NOT USE; small Bx scar at V1 which is already gone at V2.

| 035KD L3 | V1 | V2 | V3 | V4 | V5 |
|---|---|---|---|---|---|
| 3D volume | -2.3659 | -2.2949 | | | |
| 3D absol volume | 2.6174 | 2.3103 | | | |

| *Lesion Index #* | Subject # | Lesion # | Body Location | Diagnosis by Biopsy |
|---|---|---|---|---|
| *149* | **35** | **4** | Back | BCC, superficial |

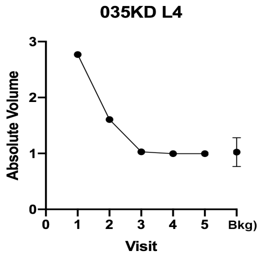

Slightly raised lesion, gone at V3

| **035KD L4** | V1 | V2 | V3 | V4 | V5 |
|---|---|---|---|---|---|
| 3D diameter | 6.85 | 5.86 | 4.51 | 4.12 | 3.05 |
| 3D perp diameter | 5.41 | 4.86 | 3.45 | 3.17 | 2.27 |
| 3D av height | 0.095 | 0.136 | 0.004 | 0.050 | 0.006 |
| 3D Av Ht/Area * 10^4 | 8.030 | 15.070 | | | |
| 3D volume | 1.873 | 1.544 | 0.913 | 0.804 | -0.736 |
| 3D absol volume | 2.768 | 1.607 | 1.028 | 0.995 | 0.966 |

Response outcome: **V3**

| Background region at visit | | | **Average bkg controls:** | |
|---|---|---|---|---|
| V1 | V2 | V5 | MEAN | SD |
| 1.320 | 0.869 | 0.880 | **1.023** | 0.257 |

| *Lesion Index #* | Subject # | Lesion # | Body Location | Diagnosis by Biopsy |
|---|---|---|---|---|
| *150* | **35** | **5** | Elbow | BCC, Superficial |

DO NOT USE

Small erosion (Bx scar?) at V1 already gone at V2.

| 035KD L5 | V1 | V2 | V3 | V4 | V5 |
|---|---|---|---|---|---|
| 3D diameter | x | x | x | x | x |
| 3D perp diameter | | | | | |
| 3D volume | 0.1415 | 0.903 | 0.0564 | 0.0926 | 0.0462 |
| 3D absol volume | 0.229 | 0.9919 | 0.0598 | 0.1661 | 0.0469 |

ff LL

on V1 l on V2 on V5

| *Lesion Index #* | Subject # | Lesion # | Body Location | Diagnosis by Biopsy |
|---|---|---|---|---|
| *151* | **35** | **6** | Clavicle | N/A |

V3 missing
V1 photo does match with others;
DO NOT USE

| 035KD L6 | V1 | V2 | V3 | V4 | V5 |
|---|---|---|---|---|---|
| 3D diameter | x | x | x | x | x |
| 3D perp diameter | | | | | |
| 3D volume | 1.6859 | 1.6581 | | | |
| 3D absol volume | 1.6859 | 1.6818 | | | |

ff LL

on V1 l on V2 on V5

| *Lesion Index #* | Subject # | Lesion # | Body Location | Diagnosis by Biopsy | Graphical data | Notes & Response to VitD | 3-D lesion data | Visit 1 | Visit 2 | Visit 3 | Visit 4 | Visit 5 | Response outcome | VDR gene |
|---|---|---|---|---|---|---|---|---|---|---|---|---|---|---|

**High-dose Vit D assignment: before V2**

Lengths are in (mm); Volumes in (mm^3)

VDR alleles for patient 36: Ff | LS

*152* | **36** | **1** | Shoulder | BCC, superficial

036DM L1

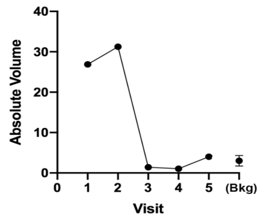

Large flat lesion, gone by V3 (lesions within old shave Bx scar)

| 036DM L1 | V1 | V2 | V3 | V4 | V5 |
|---|---|---|---|---|---|
| 3D diameter | 27.354 | 22.394 | 11.001 | 10.732 | 11.721 |
| 3D perp diameter | 11.96 | 12.798 | 9.247 | 9.096 | 9.357 |
| 3D av height | 0.073 | 0.083 | 0.015 | 0.009 | 0.048 |
| 3D Av Ht/Area * 10^4 | 0.599 | 0.857 | | | |
| 3D volume | 7.309 | 5.209 | 0.288 | -0.092 | 2.486 |
| 3D absol volume | 26.886 | 31.257 | 1.416 | 1.041 | 4.012 |

Response outcome: **V3**

Background region at visit **Average bkg controls:**

| V1 | V2 | V5 | MEAN | SD |
|---|---|---|---|---|
| 2.580 | 4.464 | 1.981 | **3.008** | 1.295 |

*153* | **36** | **2** | Cheek | BCC, superficial

036DM L2

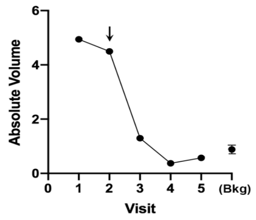

VitD=yes

Slight raised w/ atrophic center, completely gone at V4

| 036DM L2 | V1 | V2 | V3 | V4 | V5 |
|---|---|---|---|---|---|
| 3D diameter | 12.94 | 13.131 | 6.566 | 6.884 | 7.133 |
| 3D perp diameter | 6.791 | 8.137 | 4.445 | 4.497 | 4.654 |
| 3D av height | 0.038 | 0.044 | 0.014 | 0.007 | 0.002 |
| 3D Av Ht/Area * 10^4 | 1.240 | 1.238 | | | |
| 3D volume | 2.076 | 2.798 | 0.285 | 0.142 | 0.038 |
| 3D absol volume | 4.942 | 4.098 | 1.298 | 0.374 | 0.57 |

Response outcome: **V4**

Background region at visit **Average bkg controls:**

| V1 | V2 | V5 | MEAN | SD |
|---|---|---|---|---|
| 1.051 | 0.861 | 0.730 | **0.881** | 0.162 |

*154* | **36** | **3** | Temple | BCC, nodular

036DM L3

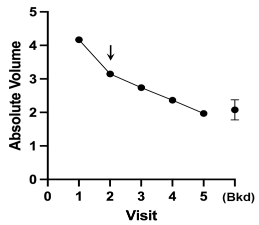

VitD = no

Depressed lesion, gone by V4 (leaving flat scar)

| 036DM L3 | V1 | V2 | V3 | V4 | V5 |
|---|---|---|---|---|---|
| 3D diameter | 10.284 | 12.309 | 9.582 | 9.58 | 9.82 |
| 3D perp diameter | 9.27 | 9.992 | 7.753 | 7.366 | 7.608 |
| 3D av height | 0.032 | 0.025 | 0.027 | 0.044 | 0.016 |
| 3D Av Ht/Area * 10^4 | 1.070 | 0.640 | | | |
| 3D volume | 2.422 | 1.894 | 1.487 | 2.245 | 0.858 |
| 3D absol volume | 4.169 | 3.149 | 2.738 | 2.364 | 1.968 |

Response outcome: **V4**

Background region at visit **Average bkg controls:**

| V1 | V2 | V5 | MEAN | SD |
|---|---|---|---|---|
| 2.208 | 2.291 | 1.731 | **2.077** | 0.302 |

| *Lesion Index #* | Subject # | Lesion # | Body Location | Diagnosis by Biopsy | Graphical data | Notes & Response to VitD | 3-D lesion data | Visit 1 | Visit 2 | Visit 3 | Visit 4 | Visit 5 | Response outcome | VDR gene |
|---|---|---|---|---|---|---|---|---|---|---|---|---|---|---|

**37- Patient lost to Follow Up**

| *Lesion Index #* | Subject # | Lesion # | Body Location | Diagnosis by Biopsy | Graphical data | Notes & Response to VitD | 3-D lesion data | Visit 1 | Visit 2 | Visit 3 | Visit 4 | Visit 5 | Response outcome | VDR gene |
|---|---|---|---|---|---|---|---|---|---|---|---|---|---|---|

**High-dose Vit D assignment: before V3**

Lengths are in (mm); Volumes in (mm^3)

VDR alleles for patient 38: ff | SS

*155* | **38** | **1** | Leg, pretibia | BCC, superficial

038SS L1

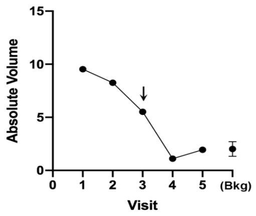

VitD=yes

Superficial lesion, gone by V4

| 038SS L1 | V1 | V2 | V3 | V4 | V5 |
|---|---|---|---|---|---|
| 3D diameter | 11.19 | 12.12 | 11.32 | 7.42 | 7.11 |
| 3D perp diameter | 8.09 | 7.92 | 6.72 | 5.82 | 4.88 |
| 3D av height | 0.173 | 0.121 | 0.098 | 0.029 | 0.048 |
| 3D Av Ht/Area * 10^4 | 5.920 | 3.830 | | | |
| 3D volume | 9.483 | 8.255 | 5.523 | 0.696 | 1.914 |
| 3D absol volume | 9.529 | 8.258 | 5.525 | 1.106 | 1.944 |

Response outcome: **V4**

Background region at visit **Average bkg controls:**

| V1 | V2 | V5 | MEAN | SD |
|---|---|---|---|---|
| 2.617 | 2.190 | 1.260 | **2.022** | 0.694 |

156 | 38 | 2 | Leg, pretibi | BCC, superficial

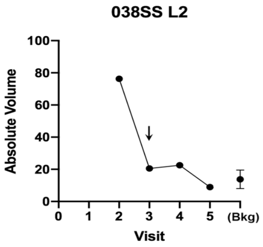

VitD =no

| 038SS L2 | V1 | V2 | V3 | V4 | V5 |
|---|---|---|---|---|---|
| 3D diameter | 19.026 | 22.528 | 19.052 | 20.137 | 17.464 |
| 3D perp diameter | 16.269 | 18.812 | 15.544 | 16.572 | 15.432 |
| 3D av height | 0.2783 | 0.282 | 0.103 | 0.117 | 0.021 |
| 3D Av Ht/Area * 10^4 | 2.850 | 2.100 | | | |
| 3D volume | 58.247 | 76.242 | 19.406 | 22.333 | 3.275 |
| 3D absol volume | 60.787 | 76.32 | 20.503 | 22.581 | 8.924 |

Superficial lesion, gone by V5

**V5**

Background region at visit **Average bkg controls:**

| V1 | V2 | V5 | MEAN | SD | SD |
|---|---|---|---|---|---|
| 16.542 | 7.099 | 17.667 | 16.504 | **14.453** | 4.932 |

157 | 38 | 3 | Leg, pretibia | BCC, superficial

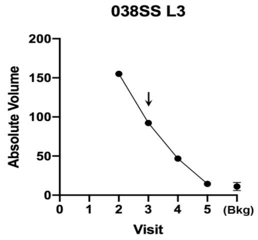

VitD = no

| 038SS L3 | V1 | V2 | V3 | V4 | V5 |
|---|---|---|---|---|---|
| 3D diameter | 21.66 | 21.58 | 20.81 | 19.38 | 18.37 |
| 3D perp diameter | 12.53 | 18.87 | 20.55 | 15.64 | 18.31 |
| 3D av height | 0.338 | 0.575 | 0.372 | 0.223 | 0.114 |
| 3D Av Ht/Area * 10^4 | 3.670 | 4.480 | | | |
| 3D volume | 64.105 | 155.085 | 92.161 | 46.333 | 13.893 |
| 3D absol volume | 64.608 | 155.087 | 92.242 | 46.827 | 14.335 |

Superficial lesion, gone by V5
Exam note says "Clear at visit 5"

**V5**

Background region at visit **Average bkg controls:**

| V1 | V2 | V5 | MEAN | SD | SD |
|---|---|---|---|---|---|
| 9.075 | 9.476 | 12.326 | 13.742 | **11.155** | 2.252 |

158 | 38 | 4 | Leg, pretibia | N/A

| 038SS L4 | V1 | V2 | V3 | V4 | V5 |
|---|---|---|---|---|---|
| 3D diameter | x | x | x | x | x |
| 3D perp diameter | | | | | |
| 3D volume | | 32.668 | 7.315 | -2.8652 | 9.2914 |
| 3D absol volume | | 32.677 | 7.5229 | 7.8033 | 9.3091 |

Raised lesion, still present at V5?
No biopsy done--- DO NOT USE

ff SS

Background region at visit **Average bkg controls:**

| | | | | MEAN | SD |
|---|---|---|---|---|---|
| 3.9923 | 2.71 | 4.3864 | 3.448 | **3.6341** | 0.726 |

| *Lesion Index #* | Subject # | Lesion # | Body Location | Diagnosis by Biopsy | Graphical data | Notes & Response to VitD | 3-D lesion data | Visit 1 | Visit 2 | Visit 3 | Visit 4 | Visit 5 | Response outcome | VDR gene |
|---|---|---|---|---|---|---|---|---|---|---|---|---|---|---|

**High-dose Vit D assignment: before V2**

Lengths are in (mm); Volumes in (mm^3)

**VDR alleles for patient 39**

| FF | LS |
|---|---|

159 | 39 | 1 | Forehead | BCC, nodular and superficial

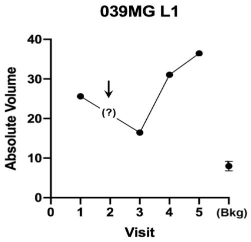

VitD: (Unable to evaluate)

| 039MG L1 | V1 | V2 | V3 | V4 | V5 |
|---|---|---|---|---|---|
| 3D diameter | 15.17 | | 14.73 | 11.17 | 16.39 |
| 3D perp diameter | 11.17 | | 10.86 | 8.85 | 12.95 |
| 3D av height | 0.727 | | 0.426 | 0.506 | 0.669 |
| 3D Av Ht/Area * 10^4 | 1.335 | | | | |
| 3D volume | 25.139 | | 16.393 | 31.047 | 35.810 |
| 3D absol volume | 25.628 | | 16.452 | 31.054 | 36.448 |

Big eroded lesion, unresponsive

**NC**

Background region at visit **Average bkg controls:**

| V1 | V2 | V5 | MEAN | SD |
|---|---|---|---|---|
| 8.009 | 9.191 | 6.794 | **7.998** | 1.199 |

160 | 39 | 2 | Glabella | BCC, nodular

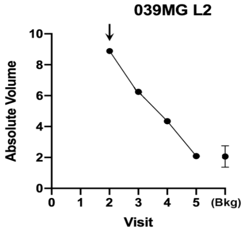

VitD = no

| 039MG L2 | V1 | V2 | V3 | V4 | V5 |
|---|---|---|---|---|---|
| 3D diameter | | 14.207 | 13.483 | 10.443 | 10.073 |
| 3D perp diameter | Photo | 11.299 | 8.57 | 8.459 | 7.832 |
| 3D av height | missing | 0.0742 | 0.075 | 0.065 | 0.021 |
| 3D Av Ht/Area * 10^4 | | 1.452 | 1.960 | 2.316 | 0.836 |
| 3D volume | | 7.030 | 4.849 | 3.095 | 0.006 |
| 3D absol volume | | 8.610 | 6.246 | 4.347 | 2.092 |

Superficial lesion, gone at V5

**V5**

Background region at visit **Average bkg controls:**

| V1 | V2 | V5 | MEAN | SD |
|---|---|---|---|---|
| 2.865 | 1.711 | 1.621 | **2.066** | 0.694 |

| Lesion Index # | Subject # | Lesion # | Body Location | Diagnosis by Biopsy | Notes & Response to VitD |
|---|---|---|---|---|---|
| 161 | 39 | 3 | Forehead | N/A | Hair causes too much volume in every picture<br>Small nodular lesion, still there at V3, visually gone at V4<br>Lots of hair noise (DO NOT USE) |

| 039MG L3 | V1 | V2 | V3 | V4 | V5 |
|---|---|---|---|---|---|
| 3D diameter | x | x | x | x | x |
| 3D perp diameter | | | | | |
| 3D volume | -0.9855 | | -5.244 | -0.7788 | -0.052 |
| 3D absol volume | 2.9299 | | 5.2652 | 1.4621 | 0.0813 |

| control on V1 | control on v3 | control on V5 | Average bkg controls: MEAN | SD | FF | LS |
|---|---|---|---|---|---|---|
| 0.1502 | 1.597 | 0.0869 | **0.611** | 0.8539 | | |

| Lesion Index # | Subject # | Lesion # | Body Location | Diagnosis by Biopsy | Notes & Response to VitD |
|---|---|---|---|---|---|
| 162 | 39 | 4 | Back | N/A | CANNOT USE; missing photos for V2, V3; gone at V4, V5 |

| 039MG L4 | V1 | V2 | V3 | V4 | V5 |
|---|---|---|---|---|---|
| 3D diameter | x | x | x | x | x |
| 3D perp diameter | | | | | |
| 3D volume | 1.8545 | | | | |
| 3D absol volume | 3.1813 | | | | |

FF LS

| *Lesion Index #* | Subject # | Lesion # | Body Location | Diagnosis by Biopsy | Graphical data | Notes & Response to VitD | 3-D lesion data | Visit 1 | Visit 2 | Visit 3 | Visit 4 | Visit 5 | Response outcome | VDR gene |
|---|---|---|---|---|---|---|---|---|---|---|---|---|---|---|

**40- Screen fail**

| *Lesion Index #* | Subject # | Lesion # | Body Location | Diagnosis by Biopsy | Graphical data | Notes & Response to VitD | 3-D lesion data | Visit 1 | Visit 2 | Visit 3 | Visit 4 | Visit 5 | Response outcome | VDR gene |
|---|---|---|---|---|---|---|---|---|---|---|---|---|---|---|

**High-dose Vit D assignment: before V2**

Lengths are in (mm); Volumes in (mm^3)

**VDR alleles for patient 41**

| FF | LL |
|---|---|

| Lesion Index # | Subject # | Lesion # | Body Location | Diagnosis by Biopsy | Notes & Response to VitD | Response outcome |
|---|---|---|---|---|---|---|
| 163 | 41 | 1 | Back, upper | BCC, nodular **(Biopsied at V5)** | VitD=No<br>Nodular lesion at edge of flat scar; not fully resolved after PDT | **NC** |

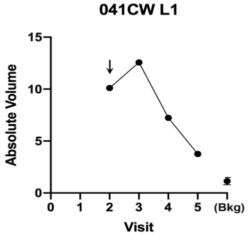

| 041CW L1 | V1 | V2 | V3 | V4 | V5 |
|---|---|---|---|---|---|
| 3D diameter | Photo missing | 10.38 | 10.02 | 8.79 | 13.55 |
| 3D perp diameter | | 8.72 | 7.82 | 5.92 | 9.58 |
| 3D av height | | 0.168 | 0.230 | 0.123 | 0.101 |
| 3D Av Ht/Area * 10^4 | | 5.86 | 9.2 | 7.25 | 2.4 |
| 3D volume | | 9.862 | 12.517 | 6.987 | 3.611 |
| 3D absol volume | | 10.104 | 12.573 | 7.228 | 3.755 |

Background region at visit **Average bkg controls:**

| V1 | V2 | V5 | MEAN | SD |
|---|---|---|---|---|
| 0.912 | 1.547 | 0.960 | **1.140** | 0.354 |

| Lesion Index # | Subject # | Lesion # | Body Location | Diagnosis by Biopsy | Notes & Response to VitD | Response outcome |
|---|---|---|---|---|---|---|
| 164 | 41 | 2 | Shoulder | N/A | VitD= no<br>Thin lesion, visually gone at V3 | **V4** |

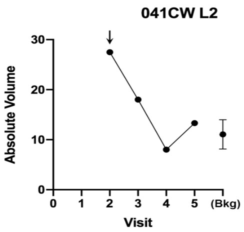

| 041CW L2 | V1 | V2 | V3 | V4 | V5 |
|---|---|---|---|---|---|
| 3D diameter | Photo missing | 23.873 | 17.933 | 19.587 | 19.105 |
| 3D perp diameter | | 19.931 | 15.3 | 17.299 | 18.032 |
| 3D av height | | 0.091 | 0.07 | 0.031 | 0.062 |
| 3D Av Ht/Area * 10^4 | | 0.609 | 0.907 | 0.290 | 0.572 |
| 3D volume | | 25.965 | 13.33 | 5.142 | 12.297 |
| 3D absol volume | | 27.482 | 16.424 | 8.015 | 13.31 |

Background region at visit **Average bkg controls:**

| V1 | V2 | V5 | MEAN | SD |
|---|---|---|---|---|
| 7.738 | 16.3 | 13.196 | **12.410** | 4.333 |

| Lesion Index # | Subject # | Lesion # | Body Location | Diagnosis by Biopsy | Notes & Response to VitD | Response outcome |
|---|---|---|---|---|---|---|
| 165 | 41 | 3 | Neck | BCC, Nodular | VitD=No<br>Large eroded lesion, nodular, continued to grow | **NC** |

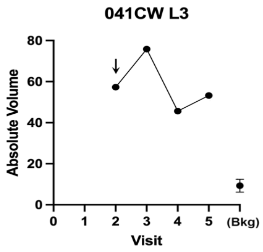

| 041CW L3 | V1 | V2 | V3 | V4 | V5 |
|---|---|---|---|---|---|
| 3D diameter | Photo missing | 17.144 | 20.512 | 17.177 | 16.046 |
| 3D perp diameter | | 13.246 | 14.907 | 13.045 | 12.496 |
| 3D av height | | 0.349 | 0.358 | 0.287 | 0.37 |
| 3D Av Ht/Area * 10^4 | | 4.810 | 3.630 | 4.010 | 5.790 |
| 3D volume | | 56.832 | 75.679 | 45.195 | 53.162 |
| 3D absol volume | | 57.309 | 75.856 | 45.62 | 53.223 |

Background region at visit **Average bkg controls:**

| V1 | V2 | V5 | MEAN | SD |
|---|---|---|---|---|
| 12.778 | 8.337 | 6.792 | **9.302** | 3.108 |

| 166 | 41 | 4 | Shoulder | BCC, Nodular |
|---|---|---|---|---|

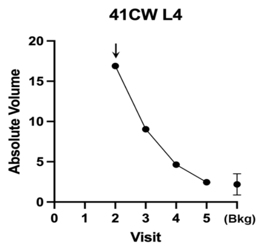

VitD=Yes

Nodular raised lesion; gone by V5

| 041CW L4 | V1 | V2 | V3 | V4 | V5 |
|---|---|---|---|---|---|
| 3D diameter | | 15.11 | 15.646 | 13.907 | 13.42 |
| 3D perp diameter | Photo | 5.566 | 4.486 | 4.453 | 3.67 |
| 3D av height | missing | 0.328 | 0.184 | 0.08 | 0.06 |
| 3D Av Ht/Area * 10^4 | | 9.770 | 5.780 | 3.020 | 2.620 |
| 3D volume | | 16.75 | 8.761 | 3.1 | 1.95 |
| 3D absol volume | | 16.899 | 9.041 | 4.641 | 2.462 |

**V5**

Background region at visit **Average bkg controls:**

| V1 | V2 | V5 | MEAN | SD |
|---|---|---|---|---|
| 3.522 | 2.165 | 0.868 | **2.185** | 1.327 |

| 167 | 41 | 5 | Arm | N/A |
|---|---|---|---|---|

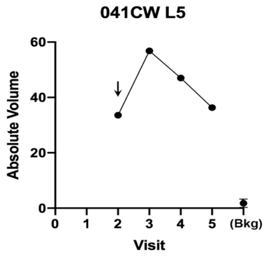

VitD=no

Big nodular lesion with telangiectasia; unresponsive.

| 041CW L5 | V1 | V2 | V3 | V4 | V5 |
|---|---|---|---|---|---|
| 3D diameter | | 13.05 | 13.37 | 14.85 | 13.43 |
| 3D perp diameter | Photo | 11.43 | 10.91 | 9.97 | 9.21 |
| 3D av height | missing | 0.501 | 0.505 | 0.391 | 0.806 |
| 3D Av Ht/Area * 10^4 | | 10.640 | 10.910 | 8.070 | 20.010 |
| 3D volume | | 33.240 | 56.346 | 46.942 | 36.291 |
| 3D absol volume | | 33.578 | 56.818 | 47.035 | 36.322 |

**NC**

Background region at visit **Average bkg controls:**

| V1 | V2 | V5 | MEAN | SD |
|---|---|---|---|---|
| 2.063 | 1.419 | 1.891 | **1.791** | 0.333 |

| *Lesion Index #* | Subject # | Lesion # | Body Location | Diagnosis by Biopsy | Graphical data | Notes & Response to VitD | 3-D lesion data | Visit 1 | Visit 2 | Visit 3 | Visit 4 | Visit 5 | Response outcome | VDR gene |
|---|---|---|---|---|---|---|---|---|---|---|---|---|---|---|

**High-dose Vit D assignment: before V2**

Lengths are in (mm); Volumes in (mm^3)

**VDR alleles for patient 42**

| FF | LS |
|---|---|

| 168 | 42 | 1 | Shoulder | BCC, superficial |
|---|---|---|---|---|

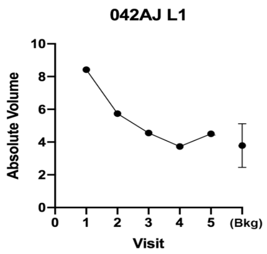

Superficial flat lesion, gone at V3 with thin scar at V4, V5

| 042AJ L1 | V1 | V2 | V3 | V4 | V5 |
|---|---|---|---|---|---|
| 3D diameter | 10.55 | 9.87 | 8.62 | 8.75 | No lesion |
| 3D perp diameter | 9.60 | 8.57 | 7.57 | 8.15 | visible |
| 3D av height | 0.121 | 0.085 | 0.082 | 0.089 | -- |
| 3D Av Ht/Area * 10^4 | 3.790 | 3.200 | 3.980 | 3.970 | -- |
| 3D volume | 7.801 | 3.971 | 4.289 | 3.727 | 3.845 |
| 3D absol volume | 8.421 | 5.739 | 4.554 | 3.732 | 4.503 |

**V3**

Background region at visit **Average bkg controls:**

| V1 | V2 | V5 | MEAN | SD |
|---|---|---|---|---|
| 4.026 | 6.979 | 3.458 | **4.821** | 1.890 |

| 169 | 42 | 2 | Chest | BCC, superficial |
|---|---|---|---|---|

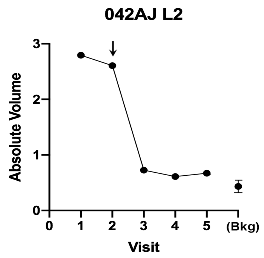

VitD = yes

Gone at V4, raised but with normal epidermis V4, V5 )

| 042AJ L2 | V1 | V2 | V3 | V4 | V5 |
|---|---|---|---|---|---|
| 3D diameter | 5.32 | 6.20 | 3.22 | 6.006 | 6.241 |
| 3D perp diameter | 4.06 | 4.00 | 2.97 | 4.345 | 3.785 |
| 3D av height | 0.087 | 0.076 | 0.037 | 0.039 | 0.035 |
| 3D Av Ht/Area * 10^4 | 12.580 | 9.310 | 12.300 | 4.630 | 4.430 |
| 3D volume | 2.784 | 2.591 | 0.646 | 0.65 | 0.604 |
| 3D absol volume | 2.794 | 2.607 | 0.727 | 0.668 | 0.67 |

**V4**

Background region at visit **Average bkg controls:**

| V1 | V2 | V5 | MEAN | SD |
|---|---|---|---|---|
| 0.554 | 0.61 | 0.522 | **0.562** | 0.045 |

| 170 | 42 | 3 | Chest | BCC, superficial |
|---|---|---|---|---|

Gone at V2 (perhaps due to inflamm from biopsy?) EXCLUDE

| 042AJ L3 | V1 | V2 | V3 | V4 | V5 |
|---|---|---|---|---|---|
| 3D diameter | 5.18 | 5.11 | | | |
| 3D perp diameter | 4.21 | 4.80 | | | |
| 3D volume | -0.356 | -0.162 | 0.259 | -0.021 | 0.259 |
| 3D absol volume | 0.482 | 0.411 | 0.425 | 0.396 | 0.425 |

FF LS

| V1 | V2 | V5 | MEAN | SD |
|---|---|---|---|---|
| 0.381 | 0.460 | 0.448 | **0.430** | 0.042 |

| Lesion Index # | Subject # | Lesion # | Body Location | Diagnosis by Biopsy | Notes & Response to VitD |
|---|---|---|---|---|---|
| *171* | **42** | **4** | Chest | BCC, superficial | Gone at V2 (perhaps due to inflamm from biopsy?) EXCLUDE |

| 042AJ L4 | V1 | V2 | V3 | V4 | V5 | | | | | |
|---|---|---|---|---|---|---|---|---|---|---|
| | | | | | | | | | FF | LS |
| 3D diameter | 6.71 | 6.42 | 5.51 | 2.17 | No lesion | | | | | |
| 3D perp diameter | 5.52 | 5.31 | 2.29 | 1.47 | visible | | | | | |
| 3D volume | 0.207 | 1.094 | 0.818 | 0.929 | 0.366 | V1 | V2 | V5 | MEAN | SD |
| 3D absol volume | 0.762 | 1.099 | 1.114 | 1.035 | 0.624 | 0.498 | 0.616 | 0.725 | **0.613** | 0.113 |

| Lesion Index # | Subject # | Lesion # | Body Location | Diagnosis by Biopsy | Notes & Response to VitD |
|---|---|---|---|---|---|
| *172* | **42** | **5** | Chest | BCC, superficial | Small flat lesion, gone at V3 |

**042AJ L5**

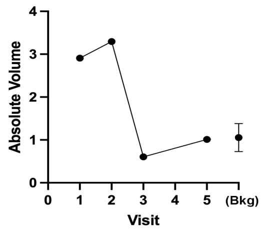

| 042AJ L5 | V1 | V2 | V3 | V4 | V5 |
|---|---|---|---|---|---|
| 3D diameter | 7.36 | 6.71 | 3.42 | | No lesion |
| 3D perp diameter | 6.89 | 6.18 | 2.43 | Photo | visible |
| 3D av height | 0.194 | 0.234 | 0.064 | missing | -- |
| 3D Av Ht/Area * 10^4 | 12.140 | 17.940 | 23.890 | | -- |
| 3D volume | 2.897 | 3.278 | -0.233 | | 0.912 |
| 3D absol volume | 2.907 | 3.295 | 0.604 | | 1.011 |

Response outcome: **V3**

Background region at visit **Average bkg controls:**

| V1 | V2 | V5 | MEAN | SD |
|---|---|---|---|---|
| 1.432 | 0.837 | 0.895 | **1.055** | 0.328 |

| *Lesion Index #* | Subject # | Lesion # | Body Location | Diagnosis by Biopsy | Graphical data | Notes & Response to VitD | 3-D lesion data | Visit 1 | Visit 2 | Visit 3 | Visit 4 | Visit 5 | | Response outcome | | VDR gene |
|---|---|---|---|---|---|---|---|---|---|---|---|---|---|---|---|---|

**High-dose Vit D assignment: before V2**

Lengths are in (mm); Volumes in (mm^3)

**VDR alleles for patient 43**

| Ff | LS |
|---|---|

| Lesion Index # | Subject # | Lesion # | Body Location | Diagnosis by Biopsy | Notes & Response to VitD |
|---|---|---|---|---|---|
| *173* | 43 | 1 | Lip, upper | BCC, Nodular | Sunken tumor or biopsy scar; redness gone V3, then no change. DO NOT USE |

| 043MG L1 | V1 | V2 | V3 | V4 | V5 | | | |
|---|---|---|---|---|---|---|---|---|
| 3D max height | 0.1292 | 0.0948 | 0.4246 | 0.0658 | 0.2262 | | | |
| 3D min height | x | x | x | x | x | | | |
| 3D volume | -3.9322 | -3.8454 | -2.571 | -2.8744 | -2.464 | | | |
| 3D absol volume | 4.1504 | 3.934 | 2.9095 | 2.8932 | 2.5188 | 0.4161 | 0.562 | 0.50395 |

| Lesion Index # | Subject # | Lesion # | Body Location | Diagnosis by Biopsy | Notes & Response to VitD |
|---|---|---|---|---|---|
| *174* | 43 | 2 | Forehead | BCC, Nodular | V5 image is missing<br>Looks like just a biopsy scar. Did not change from V1 thru V5. DO NOT USE |

| 043MG L2 | V1 | V2 | V3 | V4 | V5 | | | | Ff | LS |
|---|---|---|---|---|---|---|---|---|---|---|
| Diameter (exam) | x | x | x | x | x | | | | | |
| Height (exam) | | | | | | on V1 | l on V2 | on V5 | | |
| 3D volume | -1.5293 | -1.4981 | -2.713 | -2.0847 | | | | | | |
| 3D absol volume | 2.0603 | 2.1955 | 3.0632 | 2.4875 | | | | | | |

| Lesion Index # | Subject # | Lesion # | Body Location | Diagnosis by Biopsy | Notes & Response to VitD |
|---|---|---|---|---|---|
| *175* | 43 | 3 | Cheek | BCC, Nodular | Looks like just a biopsy scar. Did not change from V1 thru V5. DO NOT USE |

| 043MG L3 | V1 | V2 | V3 | V4 | V5 | | | 2 | Ff | LS |
|---|---|---|---|---|---|---|---|---|---|---|
| Diameter (exam) | x | x | x | x | x | | | | | |
| Height (exam) | | | | | | on V1 | l on V2 | on V5 | | |
| 3D volume | -1.5435 | | | | | | | | | |
| 3D absol volume | 1.5701 | | | | | | | | | |

| Lesion Index # | Subject # | Lesion # | Body Location | Diagnosis by Biopsy | Notes & Response to VitD |
|---|---|---|---|---|---|
| *176* | **43** | **4A** | (Ear) Post-auricular | BCC, Nodular | VitD=Yes<br>Lesion lies in a crevice<br>Big nodule; initially responded then grew at V4 and V5 |

**043MG L4A**

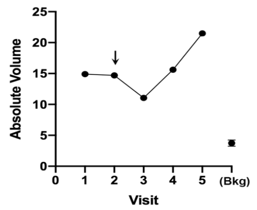

| 043MG L4 A | V1 | V2 | V3 | V4 | V5 |
|---|---|---|---|---|---|
| 3D diameter | 8.49 | 7.70 | 7.40 | 8.07 | 9.35 |
| 3D perp diameter | 6.99 | 6.94 | 5.68 | 7.07 | 6.17 |
| 3D av height | 0.414 | 0.388 | 0.363 | 0.399 | 0.459 |
| 3D Av Ht/Area * 10^4 | 21.99 | 23.07 | | | |
| 3D volume | 14.816 | 13.639 | 11.034 | 15.312 | 21.254 |
| 3D absol volume | 14.897 | 14.706 | 11.055 | 15.621 | 21.498 |

Response outcome: **NC**

Background region at visit **Average bkg controls:**

| V1 | V2 | V5 | MEAN | SD |
|---|---|---|---|---|
| 4.261 | 3.28 | 3.684 | **3.742** | 0.493 |

| Lesion Index # | Subject # | Lesion # | Body Location | Diagnosis by Biopsy |
|---|---|---|---|---|
| *177* | **43** | **4B** | (Ear) Post-auricular | BCC, Nodular |

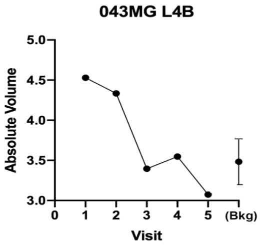

Lesion lies in a crevice
Flat (atrophic), resolved at V3

| 043MG L4 B | V1 | V2 | V3 | V4 | V5 |
|---|---|---|---|---|---|
| 3D diameter | 8.48 | 8.44 | 8.79 | No lesion visible | No lesion visible |
| 3D perp diameter | 7.04 | 6.30 | 5.03 | | |
| 3D av height | 0.041 | 0.035 | 0.004 | -- | -- |
| 3D Av Ht/Area * 10^4 | 2.160 | 2.030 | | -- | -- |
| 3D volume | -1.048 | -2.084 | -3.202 | -2.903 | -2.188 |
| 3D absol volume | 4.529 | 4.334 | 3.396 | 3.548 | 3.075 |

Response outcome: **V3**

| Background region at visit | | | Average bkg controls: | |
|---|---|---|---|---|
| V1 | V2 | V5 | MEAN | SD |
| 4.261 | 3.280 | 3.684 | **3.742** | 0.493 |

| Lesion Index # | Subject # | Lesion # | Body Location | Diagnosis by Biopsy |
|---|---|---|---|---|
| *178* | 43 | 5 | Scalp | N/A |

Bx scar at V1, hemorrhagic papule at V2; nothing to see at V3-V5
DO NOT USE

| 043MG L5 | V1 | V2 | V3 | V4 | V5 |
|---|---|---|---|---|---|
| 3D diameter | x | x | x | x | x |
| 3D perp diameter | | | | | |
| 3D volume | | | | | |
| 3D absol volume | | | | | |

| | | | Ff | LS |
|---|---|---|---|---|
| on V1 | l on V2 | on V5 | | |

| *Lesion Index #* | Subject # | Lesion # | Body Location | Diagnosis by Biopsy | Graphical data | Notes & Response to VitD | 3-D lesion data | Visit 1 | Visit 2 | Visit 3 | Visit 4 | Visit 5 | Response outcome | VDR gene |
|---|---|---|---|---|---|---|---|---|---|---|---|---|---|---|

**High-dose Vit D assignment: before V2**

**VDR alleles for patient 44**

| FF | LS |
|---|---|

| Lesion Index # | Subject # | Lesion # | Body Location | Diagnosis by Biopsy |
|---|---|---|---|---|
| *179* | 44 | 1 | Cheek | BCC, Superficial and Nodular |

Tumor already gone at V2.
DO NOT USE

| 044LG L1 | V1 | V2 | V3 | V4 | V5 |
|---|---|---|---|---|---|
| Diameter (exam) | x | x | x | x | x |
| Height (exam) | | | | | |

| Lesion Index # | Subject # | Lesion # | Body Location | Diagnosis by Biopsy |
|---|---|---|---|---|
| *180* | 44 | 2 | Clavicle | BCC, Nodular |

Tumor already gone at V2.
DO NOT USE

| 044LG L2 | V1 | V2 | V3 | V4 | V5 |
|---|---|---|---|---|---|
| Diameter (exam) | x | x | x | x | x |
| Height (exam) | | | | | |
| 3D volume | 8.6634 | -0.2574 | | | |
| 3D absol volume | 8.6634 | 0.723 | | | |

| N | 3 | FF | LS |
|---|---|---|---|

| Lesion Index # | Subject # | Lesion # | Body Location | Diagnosis by Biopsy |
|---|---|---|---|---|
| *181* | 44 | 3 | Neck | BCC, Superficial |

Tumor already gone at V2.
DO NOT USE

| 044LG L3 | V1 | V2 | V3 | V4 | V5 |
|---|---|---|---|---|---|
| Diameter (exam) | x | x | x | x | x |
| Height (exam) | | | | | |

| N | 3 | FF | LS |
|---|---|---|---|

| *Lesion Index #* | Subject # | Lesion # | Body Location | Diagnosis by Biopsy | Graphical data | Notes & Response to VitD | 3-D lesion data | Visit 1 | Visit 2 | Visit 3 | Visit 4 | Visit 5 | Response outcome | VDR gene |
|---|---|---|---|---|---|---|---|---|---|---|---|---|---|---|

**High-dose Vit D assignment: before V3**

Lengths are in (mm); Volumes in (mm^3)

**VDR alleles for patient 45**

| FF | LL |
|---|---|

| Lesion Index # | Subject # | Lesion # | Body Location | Diagnosis by Biopsy |
|---|---|---|---|---|
| *182* | **45** | **1** | Shoulder | BCC, nodular |

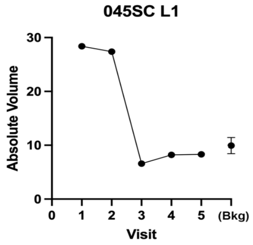

Raised lesion near white scar, visually gone at V3

| 045SC L1 | V1 | V2 | V3 | V4 | V5 |
|---|---|---|---|---|---|
| 3D diameter | 18.499 | 19.704 | 16.584 | 17.034 | 17.866 |
| 3D perp diameter | 12.901 | 15.039 | 8.875 | 8.903 | 9.32 |
| 3D av height | 0.189 | 0.133 | 0.063 | 0.069 | 0.049 |
| 3D Av Ht/Area * 10^4 | 2.440 | 1.400 | 1.240 | 1.310 | 0.844 |
| 3D volume | 28.146 | 25.051 | 5.825 | 6.597 | 5.209 |
| 3D absol volume | 28.385 | 27.4 | 6.586 | 8.211 | 8.311 |

Response outcome: **V3**

| Background region at visit | | | Average bkg controls: | |
|---|---|---|---|---|
| V1 | V2 | V5 | MEAN | SD |
| 9.833 | 10 | 9.054 | **9.630** | 0.506 |

183 | 45 | 2 | Arm | BCC, superficial and nodular

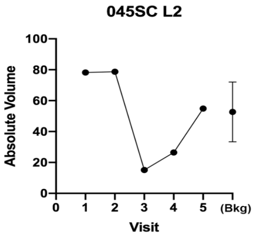

Very large, flat superficial lesion (too large for 3D to work well); visually gone at V3

| 045SC L2 | V1 | V2 | V3 | V4 | V5 |
|---|---|---|---|---|---|
| 3D diameter | 24.692 | | 22.932 | 24.275 | 23.902 |
| 3D perp diameter | 20.263 | | 20.045 | 20.873 | 21.356 |
| 3D av height | 0.167 | | 0.12 | 0.163 | 0.19 |
| 3D Av Ht/Area * 10^4 | 1.050 | | 0.827 | 1.018 | 1.181 |
| 3D volume | 75.013 | 73.135 | 12.263 | 24.293 | 54.477 |
| 3D absol volume | 78.238 | 78.676 | 15.126 | 26.478 | 54.858 |

**V3**

Background region at visit **Average bkg controls:**

| V1 | V2 | V5 | MEAN | SD |
|---|---|---|---|---|
| 37.608 | 46.04 | 74.5979 | **52.75** | 19.386 |

184 | 45 | 3 | Shoulder | BCC, nodular (Biopsied at V5)

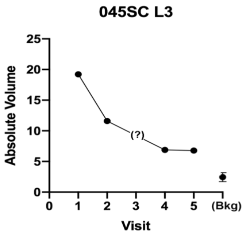

**VitD: Uable to evaluate**

Nodular, generally unresponsive

BIOPSY done on residual lesion

| 045SC L3 | V1 | V2 | V3 | V4 | V5 |
|---|---|---|---|---|---|
| 3D diameter | 13.124 | 12.85 | | Too | 8.744 |
| 3D perp diameter | 9.716 | 9.662 | | much | 6.784 |
| 3D av height | 0.227 | 0.19 | Photo | hair | 0.178 |
| 3D Av Ht/Area * 10^4 | 5.540 | 4.770 | missing | (noise) | 9.400 |
| 3D volume | 18.976 | 15.423 | | | 7.236 |
| 3D absol volume | 19.223 | 15.652 | | | 7.267 |

**NC**

Background region at visit **Average bkg controls:**

| V1 | V2 | V5 | MEAN | SD |
|---|---|---|---|---|
| 3.035 | 2.328 | 1.832 | **2.398** | 0.605 |

185 | 45 | 4 | shoulder | N/A

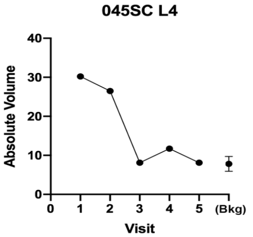

**VitD=No**

Superficial eroded lesion, leaving an atrophic scar at V3. No longer scaly at V4

| 045SC L4 | V1 | V2 | V3 | V4 | V5 |
|---|---|---|---|---|---|
| 3D diameter | 17.796 | 18.474 | 15.672 | 11.732 | 12.829 |
| 3D perp diameter | 17.047 | 16.361 | 9.148 | 8.047 | 8.715 |
| 3D av height | 0.15 | 0.117 | 0.049 | 0.003 | 0.029 |
| 3D Av Ht/Area * 10^4 | 1.573 | 1.280 | 1.012 | 0.098 | 0.796 |
| 3D volume | 27.193 | 22.288 | 4.553 | 0.17 | 2.347 |
| 3D absol volume | 30.224 | 26.489 | 8.115 | 11.725 | 8.108 |

**V3**

Background region at visit **Average bkg controls:**

| V1 | V2 | V5 | MEAN | SD |
|---|---|---|---|---|
| 13.428 | 8.558 | 7.067 | **9.684** | 3.327 |

186 | 45 | 5 | Back | N/A

V2-5 missing

ED&C was done at V1, to stop bleeding; DO NOT USE

| 045SC L5 | V1 | V2 | V3 | V4 | V5 |
|---|---|---|---|---|---|
| 3D diameter | | | | | |
| 3D perp diameter | x | x | x | x | x |
| 3D volume | 5.7455 | | | | |
| 3D absol volume | 7.6171 | | | | |

FF LL

on V1 l on V2 on V5

187 | 45 | 6 | Back | N/A

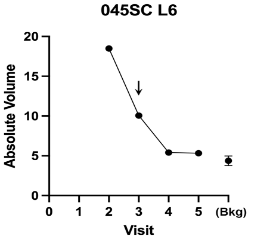

**VitD=No**

Raised lesion that shrank at V3, completely gone by V4

| 045SC L6 | V1 | V2 | V3 | V4 | V5 |
|---|---|---|---|---|---|
| 3D diameter | | 17.012 | 12.675 | 13.351 | 11.712 |
| 3D perp diameter | | 16.358 | 8.863 | 8.954 | 8.562 |
| 3D av height | | 0.025 | 0.112 | 0.024 | 0.039 |
| 3D Av Ht/Area * 10^4 | | 0.286 | 3.074 | 0.614 | 1.208 |
| 3D volume | | 4.047 | 8.168 | 1.818 | 2.624 |
| 3D absol volume | | 18.499 | 10.052 | 5.402 | 5.329 |

**V4**

Background region at visit **Average bkg controls:**

| V1 | V2 | V5 | MEAN | SD |
|---|---|---|---|---|
| 3.758 | 4.981 | 4.406 | **4.382** | 0.612 |

188 | 45 | 7 | Back | BCC, Nodular (Biopsied at V5)

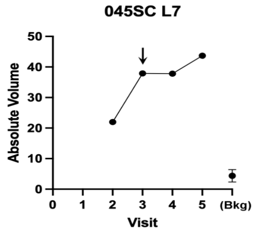

VitD=Yes

Nodular lesion that did not respond except with VitD!

| 045SC L7 | V1 | V2 | V3 | V4 | V5 |
|---|---|---|---|---|---|
| 3D diameter | | 9.845 | 11.474 | 11.665 | 13.281 |
| 3D perp diameter | | 5.858 | 7.2493 | 7.2042 | 8.1183 |
| 3D av height | | 0.526 | 0.7632 | 0.1564 | 0.5458 |
| 3D Av Ht/Area * 10^4 | | 27.160 | 27.700 | 5.590 | 15.180 |
| 3D volume | | 21.961 | 37.556 | 37.78 | 43.37 |
| 3D absol volume | | 21.967 | 37.746 | 37.82 | 43.69 |

**NC**

Background region at visit **Average bkg controls:**

| V1 | V2 | V5 | MEAN | SD |
|---|---|---|---|---|
| 5.156 | 3.271 | 4.66 | **4.362** | 0.977 |

| Lesion Index # | Subject # | Lesion # | Body Location | Diagnosis by Biopsy | Graphical data | Notes & Response to VitD | 3-D lesion data | Visit 1 | Visit 2 | Visit 3 | Visit 4 | Visit 5 | Response outcome | VDR gene |
|---|---|---|---|---|---|---|---|---|---|---|---|---|---|---|

**High-dose Vit D assignment: before V3**

Lengths are in (mm); Volumes in (mm^3)

**VDR alleles for patient 46**: FF | LS

| Lesion Index # | Subject # | Lesion # | Body Location | Diagnosis by Biopsy | Notes & Response to VitD | Response outcome |
|---|---|---|---|---|---|---|
| 189 | 46 | 1 | Shoulder | BCC, Superficial | **VitD: Uable to evaluate** Slightly raised lesion, less at V3, and V5 where it looks like scar but cannot be totally sure | V5 |

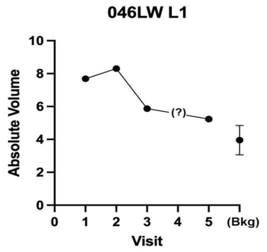

| **046LW L1** | V1 | V2 | V3 | V4 | V5 |
|---|---|---|---|---|---|
| 3D diameter | 12.664 | 10.86 | 11.244 | photo | 11.67 |
| 3D perp diameter | 12.333 | 9.901 | 9.489 | missing | 11.35 |
| 3D av height | 0.065 | 0.108 | 0.089 | | 0.065 |
| 3D Av Ht/Area * 10^4 | 1.324 | 3.190 | | | |
| 3D volume | 6.438 | 8.063 | 5.512 | | 12.474 |
| 3D absol volume | 7.691 | 8.303 | 5.867 | | 5.241 |

Background region at visit **Average bkg controls:**

| V1 | V2 | V5 | MEAN | SD |
|---|---|---|---|---|
| 3.555 | 3.336 | 4.981 | **3.957** | 0.893 |

| Lesion Index # | Subject # | Lesion # | Body Location | Diagnosis by Biopsy | Notes & Response to VitD | Response outcome |
|---|---|---|---|---|---|---|
| 190 | 46 | 2 | Shoulder | N/A | Slightly raised lesion gone at V3, replaced by a depressed scar | V3 |

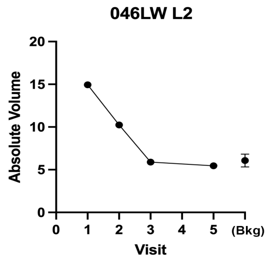

| **046LW L2** | V1 | V2 | V3 | V4 | V5 |
|---|---|---|---|---|---|
| 3D diameter | 13.179 | 13.238 | 13.165 | | 13.32 |
| 3D perp diameter | 12.07 | 9.747 | 9.96 | photo | 9.579 |
| 3D av height | 0.153 | 0.104 | 0.03 | missing | 0.046 |
| 3D Av Ht/Area * 10^4 | 3.055 | 2.506 | 0.714 | | 1.117 |
| 3D volume | 14.914 | 10.16 | 2.607 | | 4.562 |
| 3D absol volume | 14.944 | 10.244 | 5.885 | | 5.455 |

Background region at visit **Average bkg controls:**

| V1 | V2 | V5 | MEAN | SD |
|---|---|---|---|---|
| 5.292 | 6.791 | 6.162 | **6.082** | 0.753 |

| Lesion Index # | Subject # | Lesion # | Body Location | Diagnosis by Biopsy | Notes & Response to VitD |
|---|---|---|---|---|---|
| 191 | 46 | 3 | Shoulder | N/A | Thin, sunken lesion. No change. Is it just a shave biopsy scar? DO NOT USE |

| 046LW L3 | V1 | V2 | V3 | V4 | V5 |
|---|---|---|---|---|---|
| Diameter (exam) | x | x | x | x | x |
| 3D volume | -11.272 | -9.9089 | -11.02 | photo | -18.43 |
| 3D absol volume | 11.88 | 10.481 | 12.271 | missing | 18.819 |

Average bkd controls

| | | | mean | SD |
|---|---|---|---|---|
| 5.0306 | 3.822 | 4.97776 | **4.61** | 0.6831 |

| Lesion Index # | Subject # | Lesion # | Body Location | Diagnosis by Biopsy | Notes & Response to VitD | Response outcome |
|---|---|---|---|---|---|---|
| 192 | 46 | 4 | Back | BCC, nodular | Flat scaly lesion; gone by V3 | V3 |

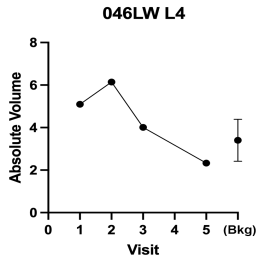

| **046LW L4** | V1 | V2 | V3 | V4 | V5 |
|---|---|---|---|---|---|
| 3D diameter | 11.79 | 12.492 | 13.07 | | 12.124 |
| 3D perp diameter | 9.757 | 8.748 | 8.809 | Photo | 8.424 |
| 3D av height | 0.057 | 0.076 | 0.098 | missing | 0.009 |
| 3D Av Ht/Area * 10^4 | 1.563 | 2.145 | 2.607 | | |
| 3D volume | 4.408 | 5.826 | 3.1 | | 0.675 |
| 3D absol volume | 5.099 | 6.145 | 4.01 | | 2.328 |

Background region at visit **Average bkg controls:**

| V1 | V2 | V5 | MEAN | SD |
|---|---|---|---|---|
| 3.958 | 2.267 | 3.991 | **3.405** | 0.986 |

| Lesion Index # | Subject # | Lesion # | Body Location | Diagnosis by Biopsy | Graphical data | Notes & Response to VitD | 3-D lesion data | Visit 1 | Visit 2 | Visit 3 | Visit 4 | Visit 5 | Response outcome | VDR gene |
|---|---|---|---|---|---|---|---|---|---|---|---|---|---|---|

**High-dose Vit D assignment: before V3**

Lengths are in (mm); Volumes in (mm^3)

**VDR alleles for patient 47**: Ff | LS

| Lesion Index # | Subject # | Lesion # | Body Location | Diagnosis by Biopsy | Notes & Response to VitD | Response outcome |
|---|---|---|---|---|---|---|
| 193 | 47 | 1 | Leg, calf | BCC, superficial and nodular | Raised lesion, gone at V3 | V3 |

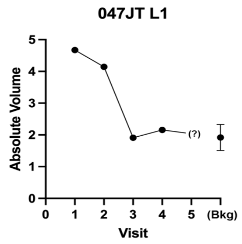

| **047JT L1** | V1 | V2 | V3 | V4 | V5 |
|---|---|---|---|---|---|
| 3D diameter | 9.687 | 8.647 | 9.116 | 9.086 | |
| 3D perp diameter | 7.888 | 5.515 | 5.661 | 5.798 | |
| 3D av height | 0.067 | 0.141 | 0.046 | 0.023 | |
| 3D Av Ht/Area * 10^4 | 2.76 | 8.95 | 2.68 | 1.32 | |
| 3D volume | 2.938 | 4.11 | 1.424 | 0.765 | photo |
| 3D absol volume | 4.672 | 4.144 | 1.909 | 2.158 | missing |

Background region at visit **Average bkg controls:**

| V1 | V2 | V5 | MEAN | SD |
|---|---|---|---|---|
| 1.525 | 2.344 | 1.886 | **1.918** | 0.410 |

| Lesion Index # | Subject # | Lesion # | Body Location | Diagnosis by Biopsy |
|---|---|---|---|---|
| 194 | 47 | 2A | Leg, calf | BCC, superficial |

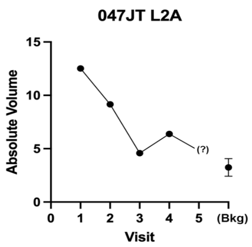

VitD = no

Raised lesion; still present at V4

| 047JT L2A | V1 | V2 | V3 | V4 | V5 |
|---|---|---|---|---|---|
| 3D diameter | 10.068 | 10.395 | 10.966 | 11.276 | |
| 3D perp diameter | 8.916 | 7.452 | 7.634 | 7.535 | |
| 3D av height | 0.177 | 0.158 | 0.069 | 0.088 | |
| 3D Av Ht/Area * 10^4 | 6.250 | 6.316 | 2.539 | 3.166 | |
| 3D volume | 11.715 | 9.042 | 4.316 | 5.911 | photo |
| 3D absol volume | 12.513 | 9.157 | 4.583 | 6.383 | missing |

*Inconclusive (no V5)*

Background region at visit **Average bkg controls:**

| V1 | V2 | V5 | MEAN | SD |
|---|---|---|---|---|
| 2.424 | 3.235 | 4.072 | **3.244** | 0.824 |

| Lesion Index # | Subject # | Lesion # | Body Location | Diagnosis by Biopsy |
|---|---|---|---|---|
| 195 | 47 | 2B | Leg, calf | BCC, superficial |

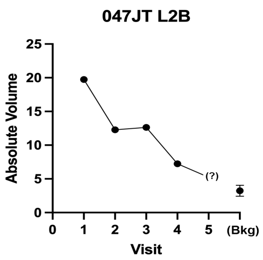

VitD=yes

Raised lesion; still present at V4;

| 047JT L2B | V1 | V2 | V3 | V4 | V5 |
|---|---|---|---|---|---|
| 3D diameter | 14.537 | 9.708 | 10.541 | 9.836 | |
| 3D perp diameter | 9.975 | 7.712 | 7.3322 | 7.924 | |
| 3D av height | 0.187 | 0.242 | 0.2679 | 0.131 | |
| 3D Av Ht/Area * 10^4 | 3.962 | 10.159 | 10.680 | 5.290 | |
| 3D volume | 19.401 | 12.264 | 12.544 | 7.203 | photo |
| 3D absol volume | 19.737 | 12.286 | 12.644 | 7.234 | missing |

*Inconclusive (no V5)*

Background region at visit **Average bkg controls:**

| V1 | V2 | V5 | MEAN | SD |
|---|---|---|---|---|
| 2.424 | 3.235 | 4.072 | **3.244** | 0.824 |

| Lesion Index # | Subject # | Lesion # | Body Location | Diagnosis by Biopsy |
|---|---|---|---|---|
| 196 | 47 | 3 | Knee | N/A |

Doubt this was BCC, as it has normal epidermal markings
DO NOT USE

| 047JT L3 | V1 | V2 | V3 | V4 | V5 |
|---|---|---|---|---|---|
| 3D diameter | x | x | x | x | x |
| 3D perp diameter | | | | | |
| 3D volume | -0.0183 | photo | -0.105 | 0.0101 | photo |
| 3D absol volume | 0.0924 | missing | 0.3632 | 0.2929 | missing |

| | | | Ff | LS |
|---|---|---|---|---|
| on V1 | l on V2 | on V5 | | |

| Lesion Index # | Subject # | Lesion # | Body Location | Diagnosis by Biopsy |
|---|---|---|---|---|
| 197 | 47 | 4 | Leg | N/A |

V5 missing
Also not be same lesion at all visits
DO NOT USE

| 047JT L4 | V1 | V2 | V3 | V4 | V5 |
|---|---|---|---|---|---|
| 3D diameter | x | x | x | x | x |
| 3D perp diameter | | | | | |
| 3D volume | 0.1297 | 0.4389 | 0.4111 | | photo |
| 3D absol volume | 0.1325 | 0.4389 | 0.4111 | | missing |

| | | | Ff | LS |
|---|---|---|---|---|
| on V1 | l on V2 | on V5 | | |

| *Lesion Index #* | Subject # | Lesion # | Body Location | Diagnosis by Biopsy | Graphical data | Notes & Response to VitD | 3-D lesion data | Visit 1 | Visit 2 | Visit 3 | Visit 4 | Visit 5 | Response outcome | VDR gene |
|---|---|---|---|---|---|---|---|---|---|---|---|---|---|---|

THIS PATIENT IS NOT USABLE--- ALL LESIONA ARE SCARS

**High-dose Vit D assignment: before V2**

**VDR alleles for patient 48**

| FF | LL |
|---|---|

| Lesion Index # | Subject # | Lesion # | Body Location | Diagnosis by Biopsy |
|---|---|---|---|---|
| 198 | 48 | 1 | Glabella | BCC, nodular |

Flat & normal at V4
looks like a biopsy scar

| 048DM L1 | V1 | V2 | V3 | V4 | V5 |
|---|---|---|---|---|---|
| 3D diameter | x | x | x | x | x |
| 3D perp diameter | | | | | |
| 3D volume | -0.5649 | -0.9481 | -0.991 | -0.5834 | 0.4858 |
| 3D absol volume | 1.0902 | 0.9586 | 1.005 | 0.6082 | 0.5362 |

| | | | FF | LL |
|---|---|---|---|---|
| on V1 | l on V2 | on V5 | | |
| 0.6373 | 0.594 | 0.9604 | | |

| Lesion Index # | Subject # | Lesion # | Body Location | Diagnosis by Biopsy |
|---|---|---|---|---|
| 199 | 48 | 2 | Cheek | BCC, nodular |

Lesion is partly covered in V1
Lesion difficult to make out
Sunken scar; I don't see any tumor
DO NOT USE

| 048DM L2 | V1 | V2 | V3 | V4 | V5 |
|---|---|---|---|---|---|
| 3D diameter | x | x | x | x | x |
| 3D perp diameter | | | | | |
| 3D volume | -1.5103 | -0.0089 | -1.523 | -1.3329 | -1.373 |
| 3D absol volume | 2.8426 | 0.588 | 2.0652 | 1.4339 | 1.4484 |

| | | | FF | LL |
|---|---|---|---|---|
| on V1 | l on V2 | on V5 | | |

| | | | | | | | | | | | |
|---|---|---|---|---|---|---|---|---|---|---|---|
| *200* | 48 | 3 | Back | BCC, superficial | Scar<br>Looks like scar; no tumor<br>DO NOT USE | 048DM L3 | V1 | V2 | V3 | V4 | V5 |
| | | | | | | 3D diameter | x | x | x | x | x |
| | | | | | | 3D perp diameter | | | | | |
| | | | | | | 3D volume | 0.5893 | 0.5081 | -0.164 | -0.1632 | 0.0776 |
| | | | | | | 3D absol volume | 1.2893 | 1.6408 | 1.3956 | 0.7018 | 0.5156 |

FF LL

on V1 l on V2 on V5

| *Lesion Index #* | Subject # | Lesion # | Body Location | Diagnosis by Biopsy | Graphical data | Notes & Response to VitD | 3-D lesion data | Visit 1 | Visit 2 | Visit 3 | Visit 4 | Visit 5 | Response outcome | VDR gene |
|---|---|---|---|---|---|---|---|---|---|---|---|---|---|---|

**High-dose Vit D assignment: before V3**

Lengths are in (mm); Volumes in (mm^3)

**VDR alleles for patient 49**

| Ff | LS |
|---|---|

### *201* — 49 — 1 — Neck — BCC, Superficial

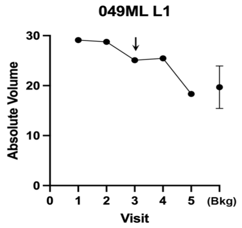

**VitD = No**

Slightly raised superficial BCC;
Visually gone at V3
Digitally gone at V5

| 049ML L1 | V1 | V2 | V3 | V4 | V5 |
|---|---|---|---|---|---|
| 3D diameter | 24.631 | 24.147 | 24.708 | 25.211 | 25.148 |
| 3D perp diameter | 14.008 | 14.133 | 13.897 | 14.435 | 14.757 |
| 3D av height | 0.14 | 0.126 | 0.051 | 0.068 | 0.033 |
| 3D Av Ht/Area * 10^4 | 1.194 | 1.095 | 0.435 | 0.551 | 0.264 |
| 3D volume | 26.366 | 22.959 | 9.895 | 13.672 | 6.359 |
| 3D absol volume | 29.123 | 28.755 | 25.092 | 25.471 | 18.349 |

Response outcome: **V5**

| Background region at visit | | | **Average bkg controls:** | |
|---|---|---|---|---|
| V1 | V2 | V5 | MEAN | SD |
| 16.344 | 18.22 | 24.482 | **19.681** | 4.262 |

### *202* — 49 — 2 — Arm — BCC, nodular and superficial

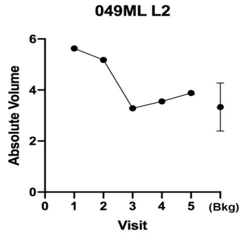

Atrophic; visually gone at V3
leaving a fibrotic-looking scar

| 049ML L2 | V1 | V2 | V3 | V4 | V5 |
|---|---|---|---|---|---|
| 3D diameter | 10.037 | 10.263 | 10.185 | 9.888 | 10.425 |
| 3D perp diameter | 8.834 | 8.749 | 8.97 | 8.893 | 9.043 |
| 3D av height | 0.013 | 0.01 | 0.002 | 0.008 | 0.003 |
| 3D Av Ht/Area * 10^4 | 0.465 | 0.352 | 0.069 | 0.289 | 0.101 |
| 3D volume | -4.097 | -4.049 | -2.995 | -2.643 | -3.567 |
| 3D absol volume | 5.629 | 5.176 | 3.281 | 3.551 | 3.881 |

Response outcome: **V3**

| Background region at visit | | | **Average bkg controls:** | |
|---|---|---|---|---|
| V1 | V2 | V5 | MEAN | SD |
| 2.274 | 3.652 | 4.075 | **3.334** | 0.942 |

### *203* — 49 — 3 — Back — N/A

V4 looks like a different lesion
Clinical note says gone at V5.
DO NOT USE

| 049ML L3 | V1 | V2 | V3 | V4 | V5 |
|---|---|---|---|---|---|
| 3D diameter | x | x | x | x | x |
| 3D perp diameter | | | | | |
| 3D volume | 1.8722 | 2.2131 | 0.3729 | photo | 0.7469 |
| 3D absol volume | 1.9755 | 2.2131 | 0.4578 | missing | 0.7485 |

Ff LS

| on V1 | l on V2 | on V5 | **Average bkd controls** | |
|---|---|---|---|---|
| | | | mean | SD |
| 0.0903 | 0.09 | 0.14401 | **0.108** | 0.031 |

### *204* — 49 — 4 — Back — N/A

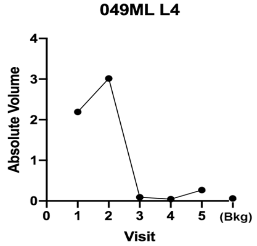

Small raised lesion, gone at V3

| 049ML L4 | V1 | V2 | V3 | V4 | V5 |
|---|---|---|---|---|---|
| 3D diameter | 4.81 | 6.45 | No lesioı | No lesion | No lesion |
| 3D perp diameter | 4.72 | 5.42 | visible | visible | visible |
| 3D av height | 0.072 | 0.126 | -- | -- | -- |
| 3D Av Ht/Area * 10^4 | 10.150 | 11.380 | -- | -- | -- |
| 3D volume | 2.121 | 3.007 | 0.037 | -0.014 | -0.226 |
| 3D absol volume | 2.191 | 3.014 | 0.093 | 0.043 | 0.270 |

Response outcome: **V3**

| Background region at visit | | | **Average bkg controls:** | |
|---|---|---|---|---|
| V1 | V2 | V5 | MEAN | SD |
| 0.076 | 0.064 | 0.100 | **0.080** | 0.019 |

| Lesion Index # | Subject # | Lesion # | Body Location | Diagnosis by Biopsy | Graphical data | Notes & Response to VitD | 3-D lesion data | Visit 1 | Visit 2 | Visit 3 | Visit 4 | Visit 5 | Response outcome | VDR gene |
|---|---|---|---|---|---|---|---|---|---|---|---|---|---|---|

**High-dose Vit D assignment: before V2**

Lengths are in (mm); Volumes in (mm^3)

**VDR alleles for patient 50**: Ff | LS

| Lesion Index # | Subject # | Lesion # | Body Location | Diagnosis by Biopsy |
|---|---|---|---|---|
| *205* | 50 | 1 | Chest | BCC, nodular |

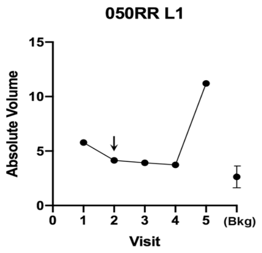

VitD = No

Relatively flat; shrank first, but then growing bigger at V5

| 050RR L1 | V1 | V2 | V3 | V4 | V5 |
|---|---|---|---|---|---|
| 3D diameter | 12.43 | 11.64 | 8.24 | 8.36 | 8.12 |
| 3D perp diameter | 8.96 | 9.21 | 7.23 | 7.51 | 7.43 |
| 3D av height | 0.059 | 0.055 | 0.080 | 0.074 | 0.146 |
| 3D Av Ht/Area * 10^4 | 1.651 | 1.611 | 4.256 | 3.741 | 7.688 |
| 3D volume | 5.266 | 3.952 | 3.914 | 3.569 | 11.150 |
| 3D absol volume | 5.782 | 4.138 | 3.923 | 3.735 | 11.206 |

Response outcome: **NC**

Background region at visit **Average bkg controls:**

| V1 | V2 | V5 | MEAN | SD |
|---|---|---|---|---|
| 2.466 | 2.353 | 1.8 | **2.206** | 0.356 |

| Lesion Index # | Subject # | Lesion # | Body Location | Diagnosis by Biopsy |
|---|---|---|---|---|
| *206* | 50 | 2 | Clavicle | BCC, superficial |

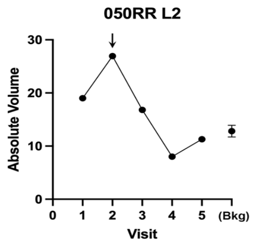

VitD=yes

Large very thin, flat, superficial lesion. Gone at V4

| 050RR L2 | V1 | V2 | V3 | V4 | V5 |
|---|---|---|---|---|---|
| 3D diameter | 30.905 | 29.472 | 27.16 | No lesion | No lesion |
| 3D perp diameter | 16.265 | 15.629 | 9.811 | visible | visible |
| 3D av height | 0.016 | 0.002 | 0.014 | -- | -- |
| 3D Av Ht/Area * 10^4 | 0.092 | 0.013 | 0.130 | -- | -- |
| 3D volume | 5.425 | 0.595 | 2.418 | 5.197 | 1.239 |
| 3D absol volume | 19.025 | 26.933 | 16.817 | 8.001 | 11.292 |

Response outcome: **V4**

Background region at visit **Average bkg controls:**

| V1 | V2 | V5 | MEAN | SD |
|---|---|---|---|---|
| 12.085 | 12.26 | 14.087 | **12.811** | 1.108 |

| Lesion Index # | Subject # | Lesion # | Body Location | Diagnosis by Biopsy |
|---|---|---|---|---|
| *207* | 50 | 3 | Shoulder | BCC, superficial |

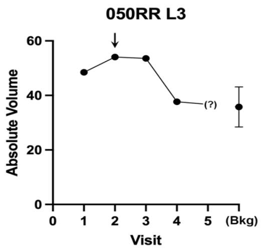

VitD=no

Large thin, flat, superficial lesion, Gone at V4

| 050RR L3 | V1 | V2 | V3 | V4 | V5 |
|---|---|---|---|---|---|
| 3D diameter | 18.845 | 18.905 | 20.084 | 19.238 | n/d |
| 3D perp diameter | 15.418 | 15.603 | 16.021 | 15.697 | n/d |
| 3D av height | 0.2547 | 0.279 | 0.254 | 0.19 | n/d |
| 3D Av Ht/Area * 10^4 | 2.760 | 2.980 | 2.480 | 1.980 | n/d |
| 3D volume | 48.419 | 54.078 | 53.537 | 37.671 | bad 3D |
| 3D absol volume | 48.452 | 54.085 | 53.56 | 37.685 | reconstruc |

Response outcome: **V4**

Background region at visit **Average bkg controls:**

| V1 | V2 | V5 | MEAN | SD |
|---|---|---|---|---|
| 31.745 | 44.3 | 31.346 | **35.796** | 7.365 |

| Lesion Index # | Subject # | Lesion # | Body Location | Diagnosis by Biopsy |
|---|---|---|---|---|
| *208* | 50 | 4 | Back | N/A |

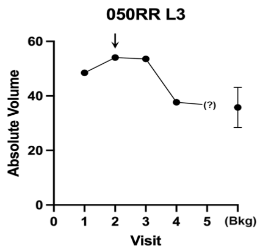

VitD=no

Flat lesion, digitally gone by V4
Clinic exam note says gone at V5

| 050RR L4 | V1 | V2 | V3 | V4 | V5 |
|---|---|---|---|---|---|
| 3D diameter | 12.211 | 10.868 | 11.206 | 11.496 | nd |
| 3D perp diameter | 7.957 | 7.105 | 7.957 | 8.151 | nd |
| 3D av height | 0.11 | 0.092 | 0.077 | 0.042 | nd |
| 3D Av Ht/Area * 10^4 | 3.440 | 3.630 | 2.670 | 1.390 | -- |
| 3D volume | 6.714 | 5.243 | 5.148 | 2.936 | nd |
| 3D absol volume | 6.737 | 5.448 | 5.481 | 3.405 | nd |

Response outcome: **V5**

Background region at visit **Average bkg controls:**

| V1 | V2 | V5 | MEAN | SD |
|---|---|---|---|---|
| 2.484 | 1.752 | 3.775 | **2.670** | 1.024 |

| Lesion Index # | Subject # | Lesion # | Body Location | Diagnosis by Biopsy |
|---|---|---|---|---|
| *209* | 50 | 5 | Back | N/A |

Photo for V3 is missing
Clinic exam note says gone at V5
DO NOT USE

| 050RR L5 | V1 | V2 | V3 | V4 | V5 |
|---|---|---|---|---|---|
| 3D diameter | x | x | x | x | x |
| 3D perp diameter | | | | | |
| 3D volume | 5.0178 | 2.034 | photo | 0.1224 | 0.0597 |
| 3D absol volume | 5.2384 | 3.0747 | missing | 0.98 | 0.5845 |

| on V1 | l on V2 | | **Average bkd controls** | |
|---|---|---|---|---|
| | | | mean | SD |
| 0.8677 | 0.724 | 0.63276 | **0.742** | 0.1184 |

| | | | | | | | | | | | |
|---|---|---|---|---|---|---|---|---|---|---|---|
| *210* | **50** | **6** | Back | N/A | | **050RR L6** | V1 | V2 | V3 | V4 | V5 |
| | | | | | | 3D diameter | x | x | x | x | x |
| | | | | | Can't use; only visible in V1, | 3D perp diameter | | | | | |
| | | | | | later images are labelled lesion 4 | 3D volume | 2.1583 | | | | |
| | | | | | DO NOT USE | 3D absol volume | 2.2386 | | | | |

| | | | | | | | | | | | | |
|---|---|---|---|---|---|---|---|---|---|---|---|---|
| *211* | **50** | **7** | Arm | N/A | | **050RR L7** | V1 | V2 | V3 | V4 | V5 | **V5** |
| | | | | | VitD = yes | 3D diameter | 15.457 | 15.451 | 15.345 | 15.207 | 15.15 | |
| | | | | | | 3D perp diameter | 12.778 | 12.342 | 12.122 | 12.242 | 12.561 | |
| | | | | | | 3D av height | 0.287 | 0.197 | 0.167 | 0.21 | 0.167 | |
| | | | | | | 3D Av Ht/Area * 10^4 | 4.580 | 3.247 | 2.818 | 3.549 | 2.769 | Background region at visit **Average bkg controls:** |
| | | | | | | 3D volume | 36.629 | 25.144 | 20.951 | 26.33 | 20.542 | V1 V2 V5 MEAN SD |
| | | | | | Superficial lesion, gone by V5 | 3D absol volume | 36.629 | 25.146 | 20.977 | 26.331 | 20.55 | 19.102 17.04 19.87 **18.670** 1.465 |

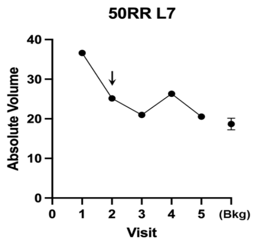
