## Supplementary figures and images for "A Clinical Trial to Determine the Impact of Tumor Size, Histological Subtype, and Vitamin D Status on the Therapeutic Response of Basal Cell Carcinoma to Photodynamic Therapy"

### Supplementary Figure S1

**SUPPLEMENTARY FIGURE S1. Flow chart of patient enrollment in the clinical trial**

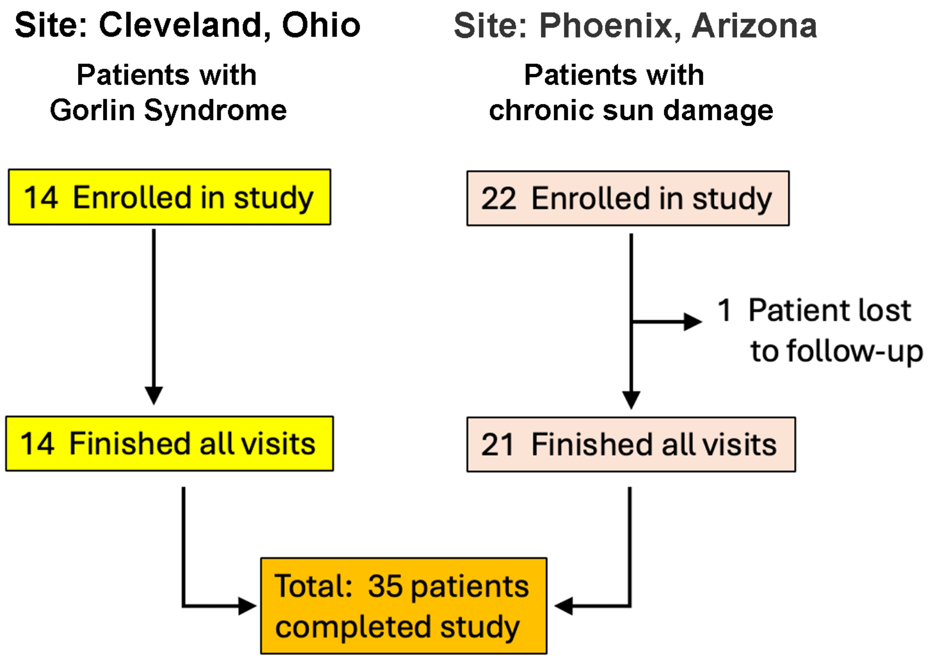

### Supplementary Figure S2

**SUPPLEMENTARY FIGURE S2. BCC lesions available for complete time course 3D analysis**

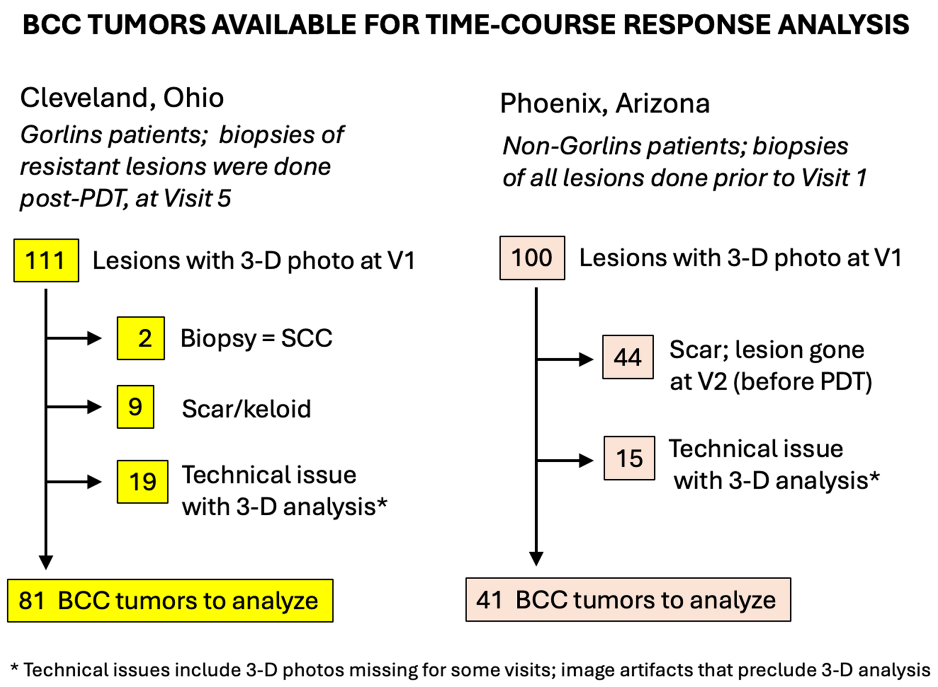
