## Supplementary Table S2 for "A Clinical Trial to Determine the Impact of Tumor Size, Histological Subtype, and Vitamin D Status on the Therapeutic Response of Basal Cell Carcinoma to Photodynamic Therapy"

### SUPPLEMENTARY TABLE S2. Comparison of BCC histological subtype, measured tumor depth (H&E), and the calculated 3D height (3DAvHt)

These are Depth-of-invasion (DOI) measurements of BCC tumors from H&E stained sections of PDT-resistant tumors, stained with H&E and digitized on a Leica scanner. The distance from the outer epidermis to the bottom of the tumor was measured using Q-Path software. Depths from 10 different locations per tumor were averaged.

| Index Number | Histological subtype category * | Original histological diagnosis by pathologist | Average DOI (microns) | 3DAvHeight (mm) |
|---|---|---|---|---|
| 1 | superficial | BCC, superficial | 129.433 | 0.025 |
| | | *mean DOI:* | **129** | **0.025** |
| 2 | nodular | BCC, nodular | 354.517 | 0.144 |
| 3 | nodular | BCC, nodular | 245.187 | 0.138 |
| 4 | nodular | BCC, nodular | 1672.2 | 0.282 |
| 5 | nodular | BCC, nodular | 564.852 | |
| 6 | nodular | BCC, superficial and nodular | 417.687 | |
| 7 | nodular | BCC, nodular | 1351.32 | |
| | | *mean DOI:* | **767.6** | **0.188** |
| | | SD | 594.3 | 0.081 |
| | | SEM | **242.6** | **0.047** |
| 8 | micronodular | BCC, nodular and microndodular | 832.511 | 0.166 |
| 9 | micronodular | BCC, nodular and microndodular | 2186.77 | 0.167 |
| 10 | micronodular | BCC, micronodular | 832.149 | 0.085 |
| 11 | micronod | BCC, nod, micronod, adenoid | 612.499 | 0.140 |
| 12 | micronodular | BCC, nodular and microndodular | 1548.5 | 0.102 |
| 13 | micronodular | BCC, nodular and microndodular | 2039.892 | 0.168 |
| 14 | micronodular | BCC, micronodular | 3080.48 | 0.473 |
| 15 | micronodular | BCC, superficial, nod, micronod | 1173.533 | 0.608 |
| 16 | micronodular | BCC, nodular and micronodular | 715.136 | |
| 17 | micronodular | BCC, nodular and micronodular | 795.158 | |
| 18 | micronodular | BCC, micronodular | 307.276 | |
| 19 | micronodular | BCC, superficial, nod, micronod | 966.945 | 0.134 |
| | | *mean DOI:* | **1258** | **0.227** |
| | | SD | 807.0 | 0.183 |
| | | SEM | **233.0** | **0.061** |
| 20 | infiltrative | BCC, infiltrative | 3090.58 | 0.318 |
| 21 | infiltrative | BCC, nodular and infiltrative | 1268.101 | 0.315 |
| 22 | infiltrative | BCC, micronodular & infiltrative | 308.343 | 0.343 |
| | | *mean DOI:* | **1556** | **0.325** |
| | | SD | 1413.2 | 0.015 |
| | | SEM | **815.9** | **0.009** |
| | "Other" | | | |
| 23 | adenoid | BCC, nodular and adenoid | 1432.455 | 0.067 |
| 24 | trichoepithelial | BCC, superf, nod, trichoepithelial | 1122.227 | 0.284 |
| 25 | trichoepithelial | BCC, superf, nod, trichoepithelial | 1183.094 | 0.223 |
| 26 | trichoepithelial | BCC, superf, nod, trichoepithelial | 767.352 | 0.243 |
| | | *mean DOI:* | **1126** | **0.204** |
| | | SD | 274.4 | 0.095 |
| | | SEM | **137.2** | **0.047** |

(Data for Graph A)

**Average Tumor Depth vs. Histologic subtype**

Depth (microns)

| Histo. Subtype* | mean | SEM | n |
|---|---|---|---|
| sBCC | 129 | 0 | 1 |
| nBCC | 767 | 242 | 6 |
| mnBCC | 1258 | 233 | 12 |
| infBCC | 1556 | 815 | 3 |
| otherBCC | 1126 | 137 | 4 |

**GRAPH A**

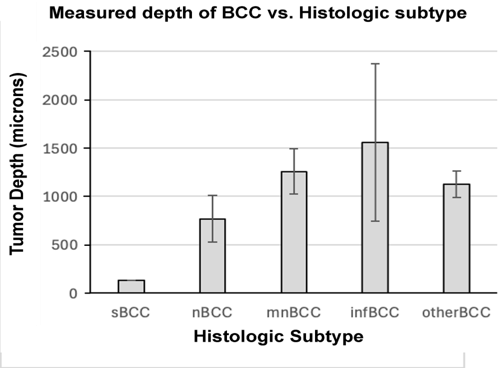


(Data for Graph B)

**Calculated 3D Height vs. Histologic Subtype**

3DAvHt (mm)

| Subtype | mean | SEM | n |
|---|---|---|---|
| sBCC | 0.025 | 0 | 1 |
| nBCC | 0.188 | 0.047 | 3 |
| mnBCC | 0.227 | 0.061 | 8 |
| infBCC | 0.325 | 0.009 | 3 |
| otherBCC | 0.204 | 0.047 | 4 |

**GRAPH B**

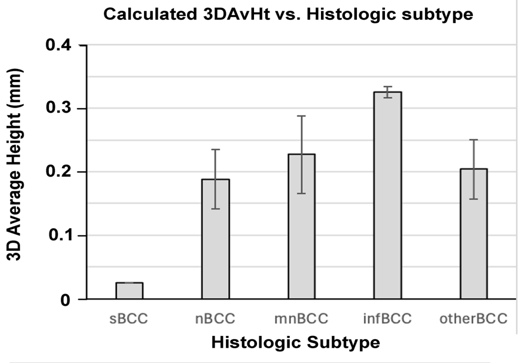


**Footnotes:**

* The most biologically aggressive subtype mentioned by the pathologists was chosen as the histologic category, using the following hierarchy: infiltrative, "other" > micronodular > nodular > superficial
