## Supplementary Table S3 for "A Clinical Trial to Determine the Impact of Tumor Size, Histological Subtype, and Vitamin D Status on the Therapeutic Response of Basal Cell Carcinoma to Photodynamic Therapy"

**SUPPLEMENTARY TABLE S3. Comparison of initial size of aggressive versus banal histologic subtypes of the subset of BCC that failed to respond to PDT**

***QUESTION:*** **For PDT-nonresponsive BCC, does histological subtype correlate with initial depth of lesion?**
***APPROACH:*** **Compare Groups 1 and 2 to see whether their tumor sizes are different.**

| Patient ID # | Lesion ID # | Location | Histological subtype of BCC * | Biopsy * prior | Biopsy * V5 | 3D AbsVol Visit 1 | 3D AbsVol Visit 2 | 3D AbsVol Mean | 3D AvHt Visit 1 | 3D AvHt Visit 2 | 3D AvHt Mean |
|---|---|---|---|---|---|---|---|---|---|---|---|
| **GROUP 1: Nodular BCC** | | | | | | | | | | | |
| **5** | 4 | back | nBCC | | • | 82.831 | 66.033 | **74.432** | 0.060 | 0.071 | **0.065** |
| **8** | 6 | back | nBCC | | • | | 14.468 | **14.468** | | 0.028 | **0.028** |
| **9** | 1 | scalp | nBCC | | • | 106.636 | 72.914 | **89.775** | 0.955 | 0.615 | **0.785** |
| **39** | 1 | Forehead | nBCC and sBCC | • | | 25.628 | | **25.628** | 0.727 | | **0.727** |
| **41** | 1 | Back | nBCC | • | | | 10.104 | **10.104** | | 0.168 | **0.168** |
| **41** | 3 | Neck | nBCC | • | | | 57.309 | **57.309** | | 0.349 | **0.349** |
| **43** | 4A | Behind ear | nBCC | • | | 14.897 | 14.7064 | **14.802** | 0.414 | 0.388 | **0.401** |
| **45** | 3 | Shoulder | nBCC | • | | 19.223 | 15.652 | **17.438** | 0.227 | 0.19 | **0.209** |
| **45** | 7 | Back | nBCC | • | | 21.967 | 37.7456 | **29.856** | 0.526 | 0.7632 | **0.645** |
| **50** | 1 | Chest | nBCC | | | 5.782 | 4.138 | **4.960** | 0.059 | 0.055 | **0.057** |
| | | | | | | | Average: | **33.877** | | Average: | **0.343** |
| | | | | | | | Std Dev | 29.4671 | | Std Dev | 0.2876 |
| | | | | | | | SEM: | **8.88468** | | SEM: | **0.08671** |
| | | | | | | | n = | 10 | | n = | 11 |
| **GROUP 2: Less common histologic subtypes of BCC **** | | | | | | | | | | | |
| **1** | 3 | forehead | nBCC and mnBCC | | • | 7.905 | 8.940 | **8.423** | 0.081 | 0.100 | **0.091** |
| **1** | 5 | low back | nBCC and mnBCC | | • | 3.029 | 3.127 | **3.078** | 0.058 | 0.060 | **0.059** |
| **5** | 6 | scapula | nBCC and infiltrBCC | | • | 4.038 | 4.421 | **4.229** | 0.124 | 0.142 | **0.133** |
| **5** | 7 | scapula | nBCC and adenBCC | | • | 7.069 | 10.128 | **8.599** | 0.060 | 0.166 | **0.113** |
| **5** | 8 | neck | mnBCC | | • | 1.561 | 1.106 | **1.334** | 0.045 | 0.026 | **0.035** |
| **5** | 9 | arm | mnB and infiltrBCC | | • | 27.065 | 17.004 | **22.035** | 0.164 | 0.101 | **0.132** |
| **6** | 1 | back | nBCC,mnBCC,adenBCC | | • | 2.996 | 2.526 | **2.761** | 0.067 | 0.102 | **0.085** |
| **9** | 2 | scalp | nBCC and mnBCC | | • | 6.599 | 3.897 | **5.248** | 0.105 | 0.064 | **0.085** |
| **9** | 4 | neck | nBCC and mnBCC | | • | 5.398 | 6.287 | **5.842** | 0.240 | 0.314 | **0.277** |
| **9** | 6 | back | mnBCC | | • | 51.243 | 57.343 | **54.293** | 0.549 | 0.611 | **0.580** |
| **14** | 6 | axilla | sBCC, nBCC, mnBCC | | • | 12.152 | | **12.152** | 0.254 | | **0.254** |
| **14** | 9 | chest | sBCC, nBCC, trichoBCC | | • | | 22.369 | **22.369** | | 0.216 | **0.216** |
| **15** | 3 | temple | sBCC, nBCC, trichoBCC | | • | 10.032 | 5.69 | **7.861** | 0.178 | 0.093 | **0.136** |
| **15** | 9 | shoulder | sBCC, nBCC, mnBCC | | • | 9.731 | 11.636 | **10.684** | 0.2 | 0.248 | **0.224** |
| **15** | 10 | back | nBCC and trichoBCC | | • | 12.194 | 13.361 | **12.778** | 0.283 | 0.35 | **0.317** |
| | | | | | | | Average: | **12.112** | | Average: | **0.182** |
| | | | | | | | Std Dev | 13.2643 | | Std Dev | 0.13871 |
| | | | | | | | SEM: | **3.42483** | | SEM: | **0.03582** |
| | | | | | 2-sided Student t-test, | | n = | 15 | | n = | 15 |
| | | | | | Group 2 vs. Group 1, | | P value = | 0.01905 | | P value = | 0.07314 |

**Footnotes:**

* Time of biopsy: Done prior to Visit 1, or at Visit 5

** Key to histological subtypes:

*sBCC* , superficial BCC
*nBCC* , nondular BCC
*mnBCC* , micronodular BCC
*trichoBCC* , trichoepitheliomatous
*infiltrBCC* , infiltrative BCC
*adenBCC* , adenoid BCC
