## Supplementary Table S4 for "A Clinical Trial to Determine the Impact of Tumor Size, Histological Subtype, and Vitamin D Status on the Therapeutic Response of Basal Cell Carcinoma to Photodynamic Therapy"

### SUPPLEMENTARY TABLE S4. Effect of tumor thickness on ability to detect responsiveness to VD + PDT

The data shown in this table are complementary to **Fig. 5** in the manuscript that describes how time course kinetics are used to determine whether or not the tumor shows enhanced clearance rate with VD (VD response = "Yes" or "No")

**HYPOTHESIS:** VitD response will be detectable primarily in thin BCC lesions, as thin lesions respond better to PDT overall.

**COROLLARY:** VitD responses will not be detectable in thick BCC lesions, which are generally nonresponsive to PDT.

**APPROACH:** Tumors were divided into two groups on the basis of 3D AvHt, using a predetermined threshold of 0.132 mm (which is the mean height of the V5 tumors in **Fig. 3D**).

**Group 1** tumors (thin): 3D AvHt < threshold. **Group 2** tumors (thick): 3D AvHt > threshold.

#### GROUP 1 LESIONS

**Thin lesions for which VitD response is "Yes"**

| Patient ID | Lesion ID | Response outcome | 3D AvHt at V1 | 3D AvHt at V2 |
|---|---|---|---|---|
| 1 | 1 | V5 | n/a | 0.044 |
| 1 | 2 | V4 | 0.058 | 0.198 |
| 1 | 5 | NC | 0.058 | 0.06 |
| 2 | 2 | V4 | 0.047 | 0.016 |
| 3 | 1 | V5 | 0.004 | 0.003 |
| 3 | 2 | V4 | 0.134 | 0.014 |
| 3 | 3 | V5 | 0.04 | 0.063 |
| 3 | 4 | V5 | 0.005 | 0.006 |
| 3 | 5 | V5 | 0.053 | 0.061 |
| 5 | 1 | V4 | 0.067 | 0.072 |
| 5 | 5 | NC | 0.079 | 0.279 |
| 5 | 6 | NC | 0.124 | 0.142 |
| 6 | 1 | NC | 0.067 | 0.102 |
| 6 | 5 | V4 | 0.009 | 0.003 |
| 7 | 3 | V4 | 0.126 | 0.079 |
| 9 | 2 | NC | 0.105 | 0.064 |
| 9 | 9 | NC | 0.061 | 0.031 |
| 12 | 6 | V4 | 0.015 | 0.018 |
| 14 | 2 | V4 | 0.036 | 0.044 |
| 14 | 7 | V5 | 0.095 | 0.144 |
| | | | **Total # of "Yes" lesions:** | 20 |
| | | | | **65%** |

**Thin lesions for which VitD response is "No"**

| Patient ID | Lesion ID | Response outcome | 3D AvHt at V1 | 3D AvHt at V2 |
|---|---|---|---|---|
| 1 | 3 | NC | 0.081 | 0.1 |
| 5 | 4 | NC | 0.06 | 0.071 |
| 5 | 8 | NC | 0.045 | 0.026 |
| 7 | 1 | V4 | 0.063 | 0.066 |
| 8 | 6 | NC | 0.028 | 0.132 |
| 9 | 8 | V5 | 0.025 | 0.021 |
| 12 | 1 | V5 | 0.045 | 0.03 |
| 12 | 2 | V4 | 0.038 | 0.04 |
| 12 | 4 | V4 | 0.047 | 0.011 |
| 12 | 3 | V4 | 0.006 | 0.008 |
| 14 | 5 | V5 | 0.116 | 0.071 |
| | | | **Total # of "No" lesions:** | 11 |
| | | | | **35%** |

#### GROUP 2 LESIONS

**Thick lesions for which VitD response is "Yes"**

| Patient ID | Lesion ID | Response outcome | 3D AvHt at V1 | 3D AvHt at V2 |
|---|---|---|---|---|
| 9 | 4 | NC | 0.24 | 0.314 |
| 9 | 6 | NC | 0.570 | 0.608 |
| 14 | 9 | NC | 0.216 | 0.191 |
| 14 | 10 | NC | 0.297 | 0.378 |
| 15 | 4 | NC | 0.203 | 0.242 |
| | | | **Total # of "Yes" lesions:** | 5 |
| | | | | **28%** |

**Thick lesions for which VitD response is " No"**

| Patient ID | Lesion ID | Response outcome | 3D AvHt at V1 | 3D AvHt at V2 |
|---|---|---|---|---|
| 5 | 9 | NC | 0.164 | 0.142 |
| 9 | 1 | NC | 0.955 | 0.615 |
| 14 | 1 | V5 | 0.198 | 0.124 |
| 14 | 4 | V5 | 0.214 | 0.256 |
| 14 | 6 | NC | 0.254 | 0.448 |
| 15 | 1 | NC | 0.928 | 0.848 |
| 15 | 2 | NC | 0.212 | 0.194 |
| 15 | 3 | NC | 0.178 | 0.093 |
| 15 | 5 | NC | 0.426 | 0.408 |
| 15 | 6 | NC | 0.661 | 0.568 |
| 15 | 7 | NC | 0.694 | 0.85 |
| 15 | 9 | NC | 0.2 | 0.248 |
| 15 | 10 | NC | 0.283 | 0.35 |
| | | | **Total # of "No" lesions:** | 13 |
| | | | | **72%** |

**Footnotes:**

Lesions that disappeared at V3 were not eligible for this analysis.

Only Cleveland patients were used here because none of their photos were missing; 49 total BCC were available for this analysis.
