## Supplementary Table S5 for "A Clinical Trial to Determine the Impact of Tumor Size, Histological Subtype, and Vitamin D Status on the Therapeutic Response of Basal Cell Carcinoma to Photodynamic Therapy"

### SUPPLEMENTARY TABLE S5. Effect of prior biopsy upon BCC lesion clearance

**Premise:** Patients at the two clinical trial sites differed as to the local immune environment of their tumors at the first study visit (Visit 1).
Patients at the Cleveland site did not require a pre-enrollment biopsy.
Patients at the Arizona site had their lesions biopsied in the weeks/months prior to Visit 1.

**Question:** Does inflammation after a biopsy affect lesion clearance rate only at Visit 1, or does it also affect PDT-mediated clearance at later visits?

**Approach:** Compare the distribution of BCC lesion clearance over time (at each visit) in the two cohorts.

#### CLEVELAND (no pre-biopsy)

| | No of lesions | (%) |
|---|---|---|
| BCC lesions at Visit 1 * | 81 | 100.0% |
| Lesions gone at V2: | 0 | 0.0% |
| Lesions gone at V3: | 28 | 34.6% |
| Lesions gone at V4: | 15 | 18.5% |
| Lesions gone at V5: | 11 | 13.6% |
| Lesions persisting at V5: | 27 | 33.3% |

**Note: These lesions were diagnosed clinically; only persistent lesions were biopsied.*

#### ARIZONA (lesions pre-biopsied) **

| | No of lesions | (%) |
|---|---|---|
| BCC lesions at Visit 1 ** | 70 | 100.0% |
| Lesions gone at V2: | 37 | 52.9% |
| Lesions gone at V3: | 12 | 17.1% |
| Lesions gone at V4: | 4 | 5.7% |
| Lesions gone at V5: | 9 | 12.9% |
| Lesions persisting at V5: | 8 | 11.4% |

***% clearance of lesions still visible at Visit 2:***

| | No of lesions | (%) |
|---|---|---|
| BCC remaining at V2: | 33 | 100.0% |
| Lesions gone at V3: | 12 | 36.4% |
| Lesions gone at V4: | 4 | 12.1% |
| Lesions gone at V5: | 9 | 27.3% |
| Lesions persisting at V5: | 8 | 24.2% |

*** Note: These lesions were all biopsied prior to Visit 1. PDT was administered at Visits 2, 3, and 4.*
