## Supplementary Table S6 for "A Clinical Trial to Determine the Impact of Tumor Size, Histological Subtype, and Vitamin D Status on the Therapeutic Response of Basal Cell Carcinoma to Photodynamic Therapy"

**SUPPLEMENTARY TABLE S6. VDR gene alleles *Fok1* and *poly-A,* versus BCC clearance after PDT.**

**Left:** Results of genotyping at the two alleles of the VDR gene, for the indicated patients.

| *Patient ID* | Sex | Age range *(Years)* | VDR alleles Fok1 * | VDR alleles Poly-A ** |
|---|---|---|---|---|
| **Cleveland study patients** | | | | |
| **1** | F | 66-70 | Ff | LS |
| **2** | F | 71-75 | FF | LS |
| **3** | F | 56-60 | ff | **SS** |
| **4** | M | 51-55 | ff | LS |
| **5** | F | 61-65 | Ff | LS |
| **6** | M | 61-65 | Ff | LS |
| **7** | M | 66-70 | Ff | **SS** |
| **8** | M | 41-45 | Ff | LS |
| **9** | M | 56-60 | Ff | LS |
| **10** | F | 21-25 | FF | **LL** |
| **12** | M | 66-70 | Ff | **SS** |
| **13** | F | 26-30 | Ff | **LL** |
| **14** | M | 61-65 | --- | --- |
| **15** | M | 66-70 | --- | --- |
| **Arizona study patients** | | | | |
| **26** | M | 61-65 | --- | --- |
| **27** | M | 71-75 | --- | --- |
| **28** | M | 71-75 | --- | --- |
| **29** | M | 56-60 | --- | --- |
| **31** | M | 71-75 | --- | --- |
| **33** | M | 61-65 | --- | --- |
| **34** | M | 46-50 | Ff | **LL** |
| **35** | F | 56-60 | ff | **LL** |
| **36** | M | 71-75 | Ff | LS |
| **38** | M | 76-80 | ff | **SS** |
| **39** | M | 46-50 | FF | LS |
| **41** | M | 66-70 | FF | **LL** |
| **42** | F | 45-50 | FF | LS |
| **43** | M | 31-35 | Ff | LS |
| **44** | F | 51-55 | FF | LS |
| **45** | M | 46-50 | FF | **LL** |
| **46** | M | 51-55 | FF | LS |
| **47** | M | 61-65 | Ff | LS |
| **48** | M | 51-55 | FF | **LL** |
| **49** | M | 66-70 | Ff | LS |
| **50** | F | 61-65 | Ff | LS |

***Notes:*** Fok1 alleles: f or F
Poly-A alleles: L (long) or S (short)

**Right:** A pattern is observed in the dual VDR genotypes: Presence of homozygous Poly-A (SS), or Fok1 (FF) is associated with a high degree of tumor clearance. Amongst patients with 3 or more BCC lesions to evaluate, genotypes that contain homozygous alleles (either SS, or FF) show more instances of 100% tumor clearance.

| Patient ID | BCC cleared/ total BCC | VDR Genotype | % BCC clear at V5 |
|---|---|---|---|
| # 03 | 6 / 6 | ff / **SS** | 100% |
| # 38 | 3 / 3 | ff/ **SS** | 100% |
| # 12 | 10 / 10 | Ff / **SS** | 100% |
| # 07 | 3 / 3 | Ff / **SS** | 100% |
| # 36 | 3 / 3 | Ff / LS | 100% |
| # 01 | 4 / 6 | Ff / LS | 67% |
| # 05 | 4 /10 | Ff / LS | 40% |
| # 06 | 4 / 5 | Ff / LS | 80% |
| # 09 | 5 / 10 | Ff / LS | 50% |
| # 08 | 3 / 4 | Ff / LS | 75% |
| # 50 | 4 / 5 | Ff / LS | 80% |
| # 02 | 3 / 3 | **FF** / LS | 100% |
| # 42 | 3 / 3 | **FF** / LS | 100% |
| # 46 | 3 / 3 | **FF** / LS | 100% |
| #49 | 3 / 3 | **FF** / LS | 100% |
| # 10 | 4 / 4 | **FF** / LL | 100% |
| # 41 | 2 / 5 | **FF** / LL | 40% |
| # 45 | 4 / 6 | **FF** / LL | 67% |

**Allelic frequencies:**

| **Fok1** | | **Poly-A** | |
|---|---|---|---|
| f = | 0.407 | S = | 0.444 |
| F = | 0.593 | L = | 0.556 |

**Genotypic frequencies:**

| **Fok1** | (%) of patients | **Poly-A** | (%) of patients |
|---|---|---|---|
| ff | 14.81% | LL | 25.93% |
| Ff | 51.85% | LS | 59.26% |
| FF | 33.33% | SS | 14.81% |
